# Prognostic performance across Alzheimer’s biomarkers, multi-modal physiological measures, and clinical history in asymptomatic individuals

**DOI:** 10.1101/2025.11.19.25340533

**Authors:** Randall J. Ellis, Audrey Airaud, Varuna H. Jasodanand, Sahana S. Kowshik, Matteo Bellitti, Vijaya B. Kolachalama, Hossein Estiri, M. Maria Glymour, Carole Dufouil, Reisa A. Sperling, David A. Bennett, Chirag J. Patel, the Alzheimer’s Disease Neuroimaging Initiative, the Australian Imaging Biomarkers and Lifestyle flagship study of ageing

## Abstract

**Importance:** Evaluating prognostic performance of Alzheimer’s biomarkers, multi-modal physiological measures, and clinical history in asymptomatic individuals versus established risk factors in asymptomatic individuals is can inform effcient screening strategies.

**Objective:** To determine and compare the prognostic performance of amyloid biomarkers, multi-modal physiological measures, and clinical/modifiable risk factors ^1^, we conducted a modality-wide assessment of predictors of AD (MODAL-AD) in cognitively asymptomatic patients.

**Design:** We used clinical trials (A4/LEARN), longitudinal cohorts (ADNI, AIBL, HABS, NACC, OASIS), and the UK Biobank spanning 2004-2025 (median follow-up time range: 1.8-13.72 years) in time-varying survival and binary classification analyses.

**Setting:** Settings included a United States clinical trial, longitudinal cohort studies spread across medical centers in the United States and Australia, and the volunteer-based UK Biobank.

**Participants:** Patients were cognitively asymptomatic and age 65+ at baseline, and potentially progressed to either clinical impairment, clinical AD diagnosis, or incurred AD ICD-codes. Patients were volunteer or convenience samples.

**Exposures:** PTau-217, amyloid-PET, CSF markers (AB1-42, pTau-181, total-Tau), plasma proteomics, multimodal brain-imaging, and cognitive tests were evaluated as predictors, along with demographics (age, sex, education), APOE genotype, and modifiable risk factors in the 2024 Lancet report ^1^.

**Main Outcome(s) and Measure(s):** PTau-217 and amyloid-PET from A4/LEARN were used to predict clinical impairment (CDR score of 0.5+ on two consecutive visits). PTau-217, amyloid-PET imaging across five cohorts, and CSF markers were used to predict clinical AD diagnosis. Plasma proteomics, multimodal neuroimaging, and cognitive assessments from the UK Biobank were used to predict AD ICD-codes.

**Results:** Sample-sizes ranged from 356-28,533 (31-519 cases; female percentages: 48.45-67.39). Models of demographics, APOE genotype, and risk-factors as predictors did not show statistically significant differences in time-dependent area under the receiver operating characteristic curve (AUROC) compared to separate models using amyloid biomarkers. Predicting cognitive impairment in A4/LEARN, pTau-217 improved AUROC by 0.045–0.084 (best: 0.616 (CI: 0.51-0.723) vs. 0.7 (CI: 0.609-0.793)). Amyloid-PET improved AD prediction (maximum AUROC increase 0.074; 0.561 (CI: 0.468-0.653) vs. 0.635 (CI: 0.537-0.733)), and CSF biomarkers showed slightly larger gains (maximum AUROC increase 0.127; 0.627 (CI: 0.438-0.816) vs. 0.754 (CI: 0.577-0.931)). In UK Biobank analyses, mean AUROC improvements were minor across proteomics (0.044), neuroimaging (0.143, with 99.8%/0.2% class-balance), and cognitive tests (0.064).

**Conclusions and Relevance:** In cognitively asymptomatic populations, biomarkers offer limited advantage over demographics, APOE genotype, and modifiable risk factors, supporting their importance in early AD screening strategies.

**Key Points:** 

**Question:** How does the prognostic performance of amyloid biomarkers (i.e., pTau-217, amyloid-PET, cerebrospinal fluid markers) and discovery-driven modalities (i.e., plasma protoemics, multimodal brain imaging, cognitive tests) compare to demographics and modifiable risk factors for predicting clinical impairment, clinical AD diagnosis, and AD ICD code outcomes in asymptomatic patients?

**Findings:** In this prognostic study of *>*300,000 patients, across cohorts, physiological modalities, and outcomes, predictive performance of demographics and modifiable risk factors did not statistically significantly differ from amyloid biomarkers, plasma proteomics, and other modalities.

**Meaning:** Alzheimer’s screening in asymptomatic patients can benefit from incorporating modifiable risk factors as additional predictors to amyloid biomarkers.

## 1 Introduction

Dementia is a leading cause of death, with 55 million current and 10 million new cases annually ^2^, with Alzheimer’s disease (AD) accounting for 60-70%. The dominant theory of AD is the amyloid cascade hypothesis, expanded to the Amyloid-Tau-Neurodegeneration (ATN) framework ^3^.

Diagnostics (e.g., pTau-217) are adept at predicting the presence of brain amyloid ^4–8^. However, a recent systematic review identified a high risk of bias in 90% of diagnostic studies of pTau-217 for not using predefined or externally-derived thresholds ^9^, and community-based autopsy samples have shown that over 30% of people with biomarker-based AD were dementia-free until death ^10^.

As 15-47% of cognitively asymptomatic patients are amyloid-positive across age-ranges ^11^, quantifying the prognostic performance of these biomarkers ^12–20^ is an important goal and could differentiate between patients at imminent risk versus exhibiting pathology without near-term consequence, informing prognosis, trial-enrollment, and utility of screening. A recent piece ^21^ highlighted potential issues in screening: pressure to seek anti-amyloid treatment with potential side effects; false positive diagnoses causing issues with driver licensing, insurance, and stoking depression; misinterpretation of symptoms; influence on advance care plans (e.g., do-not-resuscitate orders).

Previous discussions have highlighted the potential of these markers as screening tools in cognitively normal patients ^22–24^. How well do AD biomarkers and other modalities predict cognitive impairment or AD diagnosis compared to demographic and modifiable risk factors?

Acceptable performance benchmarks for the use of amyloid biomarkers as confirmatory or triaging tests in primary or secondary care have been given ^25^. If amyloid biomarkers are to be used for screening, then the low prevalence of amyloid positivity would limit their utility, among other challenges ^26^. Given the recent Food and Drug Administration approval of the first amyloid biomarker using the pTau-217/amyloid-beta 1-42 ratio in plasma ^27^ in “adult patients, aged 55 years and older, exhibiting signs and symptoms of the disease,” it is critical to understand whether these markers can predict future cognitive impairment or AD diagnosis in cognitively asymptomatic patients relative to modifiable risk factors.

Parallel to the investigation of amyloid biomarkers is the development of discovery-driven physiological modalities such as plasma proteomics, multi-modal brain imaging, and cognitive testing. These avenues provide novel, high-dimensional wellsprings of potential biomarkers that can also galvanize examination of complementary mechanisms of AD risk.

Here, we evaluated the performance of amyloid-predictive modalities (pTau-217, amyloid-PET, CSF markers), along with novel, multi-modal physiological modalities that are not established AD biomarkers–high-throughput plasma proteomics, multi-modal brain imaging, and cognitive/affective tests–for predicting cognitive impairment, AD diagnosis, as well as all-cause dementia diagnosis in cognitively asymptomatic patients. We introduce the MODALity-wide assessment for AD (MODAL-AD) to systematically evaluate predictive prognostics and emerging biomarkers. MODAL-AD is the first comprehensive, fair, and transparent view of the predictive landscape for AD. We compare all biomarkers to known risk factors including (a) demographics (i.e., age, sex, education), (b) APOE genotype, and (c) modifiable risk factors that have accumulated compelling evidence highlighted by the recent Lancet working group on dementia ^1^. We used both 1) time-to-event and 2) binary classification with automated machine learning approaches, and reported comprehensive metrics to evaluate our modeling approaches. We applying stratified cross-validation (by outcome) and strict separation of training and test data to ensure honest, out-of-sample evaluation to estimate real-world performance.

We evaluated three particular outcomes 1. First, we evaluated clinical impairment, defined as a Clinical Dementia Rating score of 0.5 or greater on two consecutive visits in the A4/LEARN study, as previously described ^28^. Second, we predicted clinically adjudicated AD diagnosis in the Alzheimer’s Disease Neuroimaging Initiative (ADNI), Australia Imaging, Biomarkers, and Lifesetyle (AIBL), Harvard Aging and Brain Study (HABS), National Alzheimer’s Coordinating Center (NACC), and Open Access Series of Imaging Studies (OASIS) cohorts. Finally, we predicted AD International Classification of Diseases-10 codes in the UK Biobank.

Across multiple aging and biobank cohorts and modalities, we did not find robust predictive performance beyond baselines of demographics and modifiable risk factors. While these results have implications for the clinical utility of the examined modali-ties ^29,30^, they also indicate the importance of broadly testing the Lancet risk factors for AD risk assessment and invite investigations of how to use the Lancet factors in clinical practice on an individual case level.

To our knowledge, this is the first study to evaluate six predictive modalities across seven cohorts using multivariate time-to-event and machine learning approaches to comprehensively evaluate prediction of clinical impairment, clinical AD diagnosis, and AD ICD codes.

## 2 Results

We deployed MODAL-AD to evaluate established and emerging modalities as potential predictors of clinical impairment, AD diagnosis, or AD ICD codes in cognitively asymptomatic patients (Figure 1). We discuss findings in each cohort and modality in turn.

**Fig. 1:**
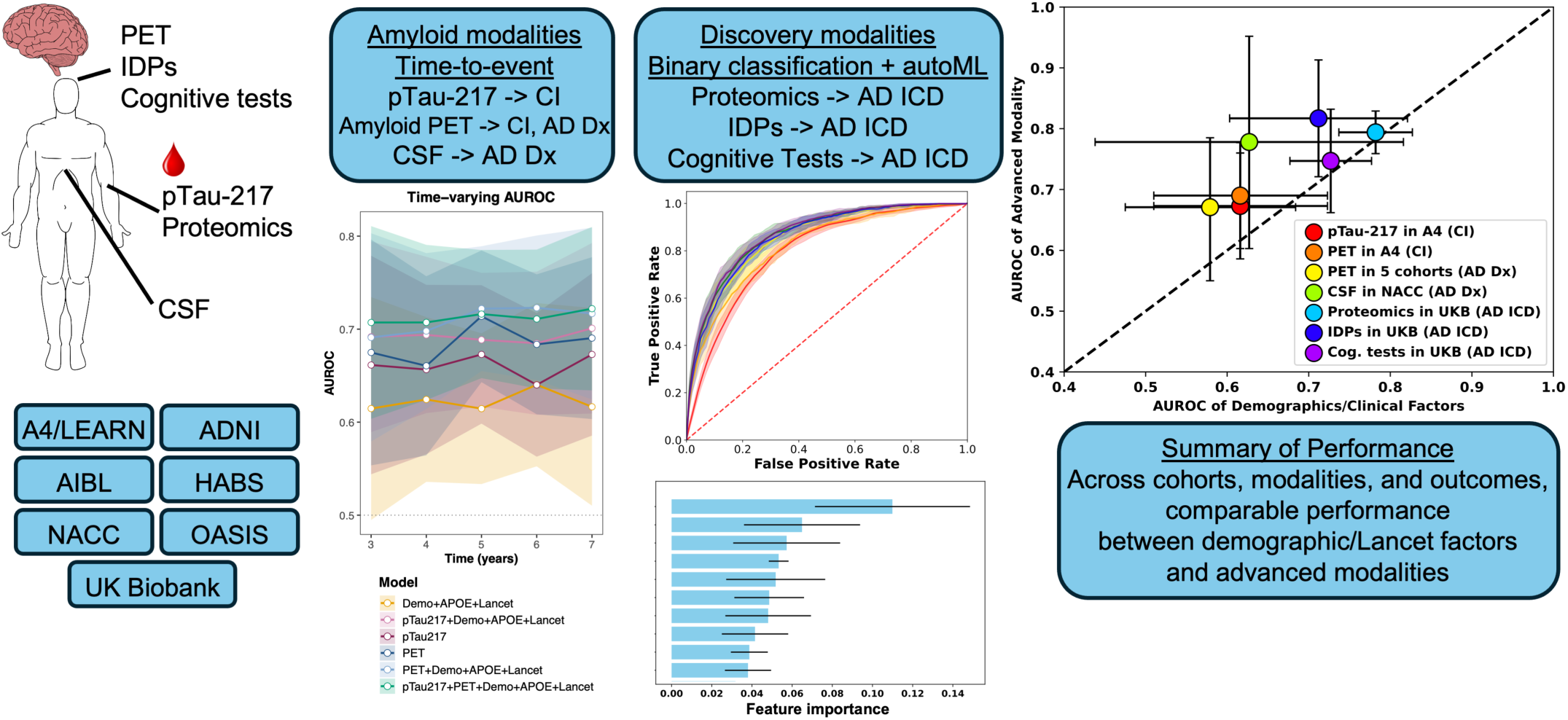
Summary of MODAL-AD and results. IDPs: Imaging-derived phenotypes. Inclusion criteria across cohorts were no history of clinical impairment or any form of dementia. In A4/LEARN, participants needed at least one pTau-217 measurement and one amyloid-PET scan, both before reaching criterion for clinical impairment. For the five PET cohorts (i.e., ADNI, AIBL, HABS, NACC, OASIS), participants needed at least one PET scan prior to any clinical impairment or dementia diagnosis. For prediction from CSF markers in NACC, participants needed at least one set of CSF marker measurements before any diagnosis of clinical impairment or dementia. For the UK Biobank, patients needed either proteomics (*→*2900 proteins), IDPs (*→*3900) or cognitive tests (*→*40 tests) measured before any ICD codes for dementia. A4: Anti-Amyloid Treatment in Asymptomatic Alzheimer’s. LEARN: Longitudinal Evaluation of Amyloid Risk and Neurodegeneration. ADNI: Alzheimer’s Disease Neuroimaging Initiative. AIBL: Australian Imaging, Biomarker & Lifestyle. HABS: Harvard Aging Brain Study. NACC: National Alzheimer’s Coordinating Center. OASIS: Open Access Series of Imaging Studies. CSF: cerebrospinal fluid. IDPs: Imaging-derived phenotypes. ICD: International Classification of Diseases.

### 2.1 pTau-217 and amyloid-PET in the A4/LEARN cohort to predict Clinical Dementia Rating score-based clinical impairment

First, we examined the prediction of clinical impairment (CDR-score of 0.5+ on two consecutive visits ^28^; N=1031 controls, 409 cases) in the A4/LEARN study using a five-fold stratified CV procedure, using pTau-217, amyloid-PET, demographics, APOE genotype, and Lancet modifiable risk factors as predictors in time-varying Cox proportional hazards models (Figure 2). We assessed performance at multiple time-points (Years 3-7) across CV folds to ensure both temporal resolution and robustness to sampling variability ^31^. This yielded 25 predictive scenarios, i.e., five yearly time points and five CV-folds, yielding 25 total p-values (corrected for multiple hypothesis-testing ^31^). PTau-217 was associated with clinical impairment (mean log hazard ratio: 1.62 (SD: 0.13), p-value range: 3.39e-33 - 8.34e-9 across scenarios; Table S2, Table S1) but did not significantly increase predictive performance beyond demographics, APOE, and Lancet factors. The range of increases in mean AUROC when adding pTau-217 was 0.045-0.084 compared to Demographics+APOE+Lancet (0.616 (CI: 0.51-0.723) vs. 0.701 (CI: 0.609-0.793); Figure 2B). 6/25 p-values were statistically significant (Figure S1), occurring at Years 4 (2/5 folds), 5(1), and 7(3). Years 3 and 6 had no significant p-values, and neither did Folds 1 and 5, pointing to a lack of robustness (Table S3). 3/6 significant p-values were all at 7 years, which had less stable estimates due to lower sample sizes (i.e., by 7 years, more patients had been censored or experienced the event). All AUROC statistics are in Table S6. We observed similarly small differences in mean sensitivity (range: 0.042-0.072), specificity (0.002-0.027), PPV (0.0002-0.0067), NPV (0.0003-0.0026), and Brier score (0.0022-0.0069) across time-points (Figure 2C-D).

**Fig. 2:**
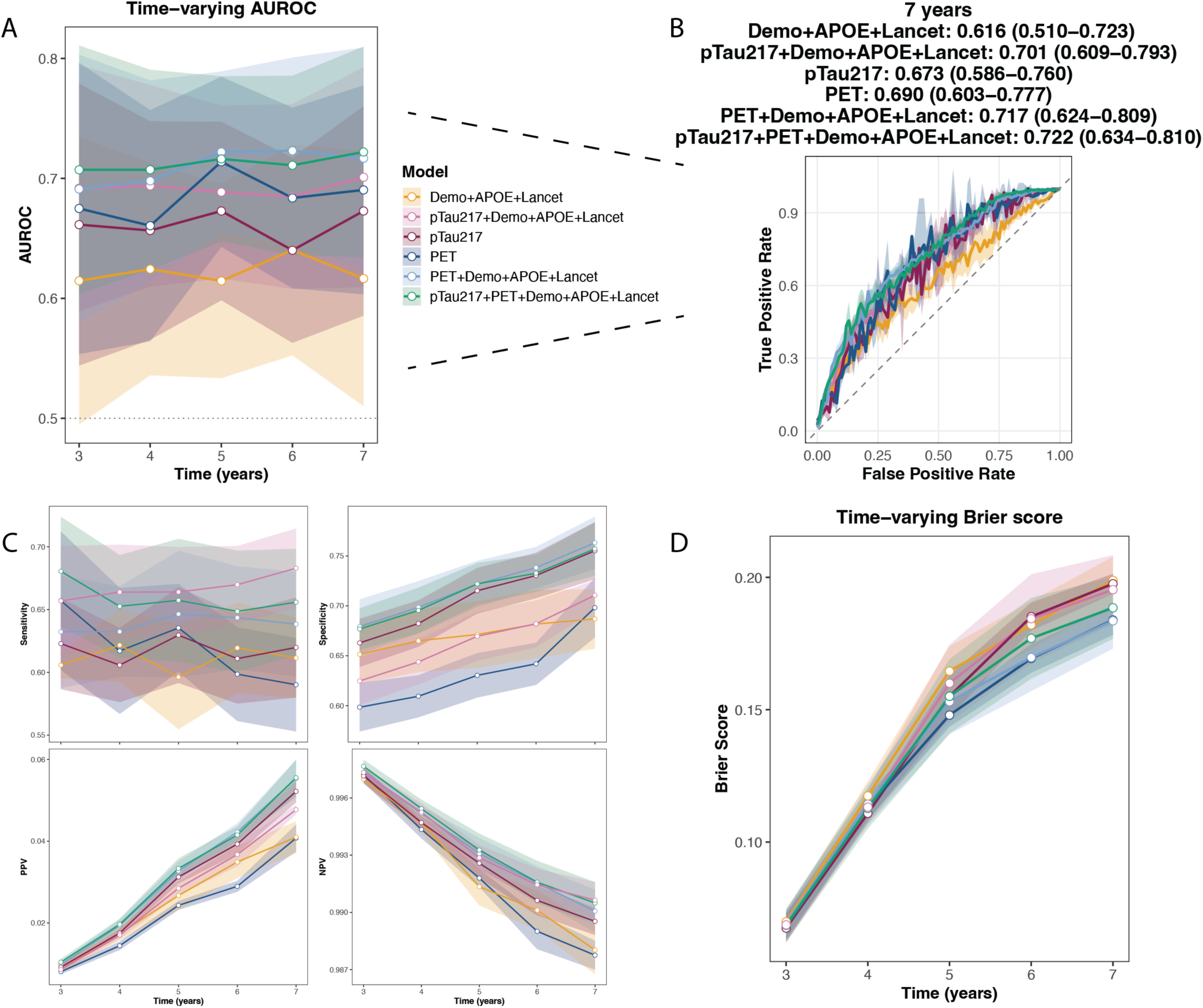
Predicting CDR-based clinical impairment using six models: 1) Demo+APOE+Lancet (demographics, APOE geno-type, and Lancet modifiable risk factors), 2) pTau217+Demographics+APOE+Lancet, 3) pTau-217, 4) Amyloid PET, 5) PET+ Demographics+APOE+Lancet, 6) pTau217+PET+Demographics+APOE+Lancet. A) Time-varying AUROC for each year, with 95% confidence intervals. B) ROC curve for Year 7, which showed the largest difference in mean AUC between Demo+Lancet and pTau217+Demo+Lancet. C) Sensitivity, specificity, positive predictive value, and negative predictive value for all six models using optimal cutpoints by Youden’s J statistic on the training set. D) Time-varying Brier score for each year, with standard deviations.

Next, we combined pTau-217 and amyloid-PET in A4/LEARN. Amyloid-PET was significantly associated with clinical impairment (mean log hazard ratio: 1.01 (SD: 0.0035), p-value range: 3.16e-32 - 0.0017; Table S2). The largest difference in mean AUROC between PET+Demographics+Lancet and Demographics+Lancet was 0.108 at 5 years compared to Demographics+APOE+Lancet (0.615 (CI: 0.533-0.696) vs. 0.722 (CI: 0.655-0.789)), indicating increased predictive performance. 15/25 p-values were statistically significant (Figure S2), occurring at Years 3 (1/5 folds), 4 (2), 5 (5), 6 (3), 7 (4) (Table S4).

Comparing PET to pTau-217, the mean difference in AUROC between PET+Demographics+APOE+Lancet and pTau-217+Demographics+APOE+Lancet ranged from 0.0007 to 0.038 across time-points.

Geriatric Depression Scale score, a Lancet factor, showed consistent statistical significance in these analyses. Across all models including Lancet factors, GDS score was significantly associated with clinical impairment (mean hazard ratio: 1.28 (SD: 0.035), p-value range: 6.52e-13 - 3.04e-5).

These results were consistent when only including patients who were amyloid-positive at enrollment time (Figure S4).

Using the PACC score as a continuous outcome, Demographics+APOE+Lancet had a similar *R*^2^ (mean: 0.125, SD: 0.026) to pTau-217 alone (mean: 0.137, SD: 0.033), but inferior to pTau-217+Demo+Lancet (mean: 0.199, SD: 0.029) (FigureS5).

### 2.2 pTau-217 in ADNI to predict AD

Next, we predicted clinical AD diagnosis in the ADNI cohort using pTau-217, demographics, APOE genotype, and Lancet modifiable risk factors using the same survival analysis approach. We assessed predictive performance at six time-points (Years 2-7) across five cross-validation folds for 30 total predictive scenarios.

Again, pTau-217 was statistically associated with AD (mean hazard ratio: 2.37 (SD: 0.4), p-value range: 4.12e-11 - 1.13e-3, Figure S6A; Table S8) but did not increase predictive performance when added to a model consisting of demographics, APOE genotype, and Lancet factors.

The range of differences in mean AUROC was 0.021-0.04, with 0.04 at 7 years compared to Demographics+APOE+Lancet (0.927 (CI: 0.885-0.97) vs. 0.967 (CI: 0.943-991); Figure S6B). 9/30 p-values were statistically significant, occurring at Years 2 (3/5 folds), 3 (1), 4 (2), 5 (1), 6 (1), and 7(1) (Table S9).

These results remained largely unchanged when using all-cause dementia as the outcome (Figure S7).

### 2.3 Amyloid-PET in five cohorts to predict AD

Next, we predicted AD in ADNI, AIBL, HABS, NACC, and OASIS cohorts using amyloid-PET, demographics, and APOE genotype, as these cohorts did not all measure Lancet factors.

Amyloid-PET was associated with AD diagnosis (mean log hazard ratio: 1.013 (SD: 0.0015), p-value range: 1.05e-36 - 4.79e-14 (Table S10, S12), but its predictive performance in combination with demographics and APOE genotype was comparable to demographics and APOE alone across nine yearly time points (Figure 3). The range of differences in mean AUROC was 0.055-0.074, with 0.074 at 5 years compared to Demographics (0.561 (CI: 0.468-0.653) vs. 0.635 (CI: 0.537-0.733); Figure 3B; Table S14). 15/45 p-values were statistically significant (Figure S8) at Years 4 (1), 5 (3), 6 (2), 7 (1), 8 (2), 9 (3), 10 (3) (Years 2 and 3 had none; Table S3), pointing to a lack of robust differences.

**Fig. 3:**
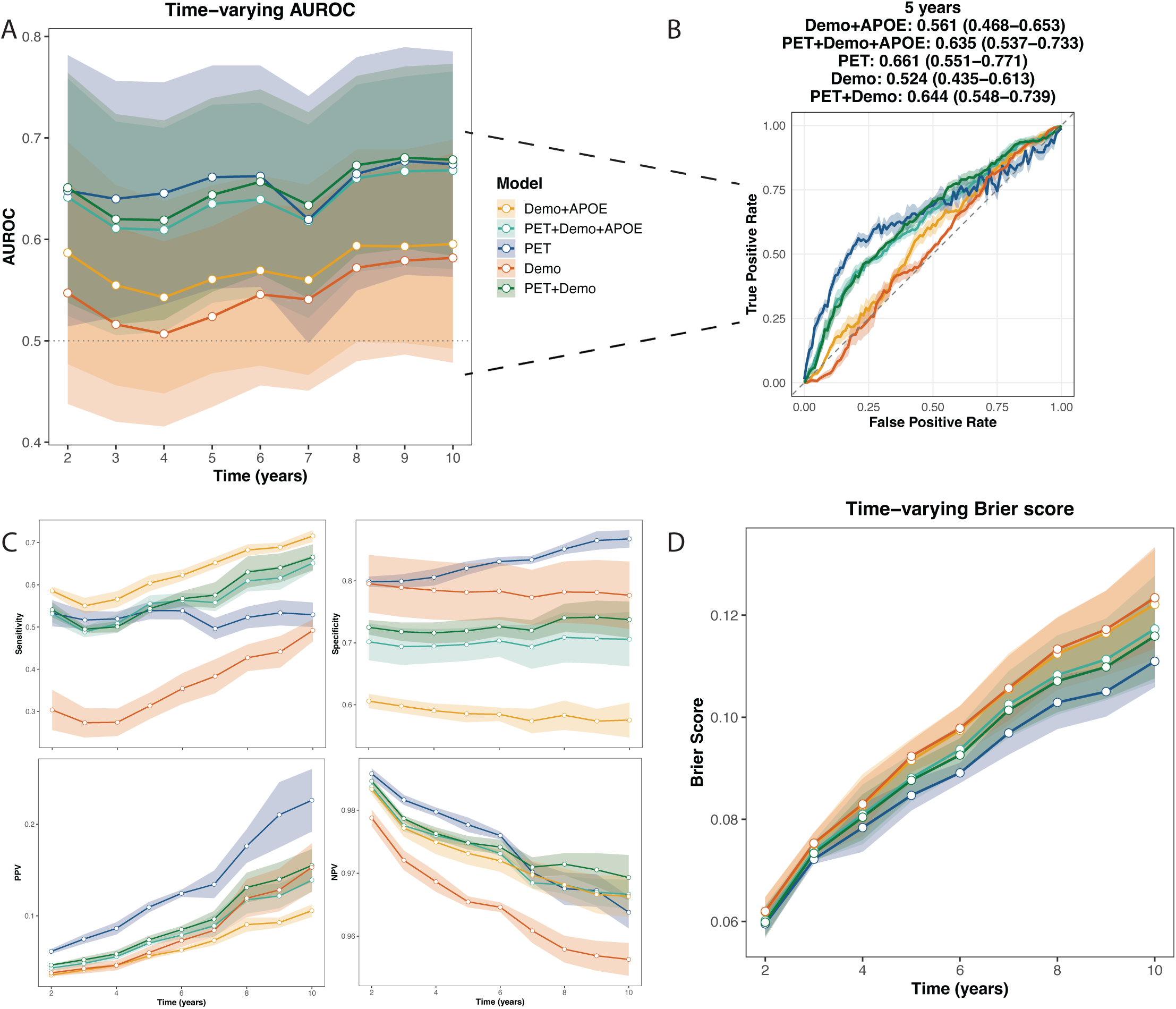
Predicting Alzheimer’s diagnosis across five PET cohorts using five models: 1) Demographics (age, sex, educa-tion)+APOE genotype, 2) Amyloid PET+demographics+APOE, 3) PET, 4) Demographics, 5) PET+Demographics. A) Time-varying AUROC for each year, with 95% confidence intervals. B) ROC curve for Year 5, which showed the largest dif-ference in mean AUC between Demo+APOE and PET+Demo+APOE. C) Sensitivity, specificity, positive predictive value, and negative predictive value for all five models using optimal cutpoints by Youden’s J statistic on the training set. D) Time-varying Brier score for each year, with standard deviations.

Demographics+APOE showed increased mean sensitivity (range: 0.048-0.094), but decreased specificity (0.096-0.126), PPV (0.007-0.027), NPV (0.0013-0.0016), and increased Brier score (0.0013-0.0052) across time-points (Figure 3C-D).

These results remained largely unchanged when using all-cause dementia as the outcome (Figure S9).

### 2.4 Cerebrospinal fluid amyloid-beta42, pTau-181, total tau, and pTau-181/Abeta42 ratio in NACC to predict clinical AD

Next, we predicted AD in NACC using CSF Abeta42, pTau-181, total tau, and pTau-181/Abeta42 ratio, along with demographics, APOE, and Lancet factors (Table S15). Abeta42 was significantly associated with AD diagnosis (mean HR: 0.62 (SD: 0.013), p-value range: 1.79e-6 - 2.5e-5 across models including it), along with pTau-181/Abeta42 ratio (mean HR: 1.39 (SD: 0.2), p-value range: 2.13e-9 - 4.67e-7) and pTau-181 (mean HR: 1.21 (SD: 0.16), p-value range: 0.0015 - 0.98), while total tau was not significant (mean HR: 1.09 (SD: 0.07), p-value range: 0.126 - 0.715). CSF markers showed predictive performance comparable to the Demographics+APOE+Lancet model across nine yearly time points; Figure 4). The range of mean AUROC differences between Demo+APOE+Lancet and CSF+Demo+APOE+Lancet was 0.072-0.127, with 0.127 at 2 years (0.627 (CI: 0.438-0.816) vs. 0.754 (CI: 0.577-0.931); Figure 4B; Table S17). 20/45 p-values were statistically significant (Figure S10) at Years 2 (2/5 folds), 3 (2), 4 (2), 5 (2), 6 (3), 7 (3), 8 (2), 9 (2), 10 (2) (Table S16), showing a lack of robustness.

**Fig. 4:**
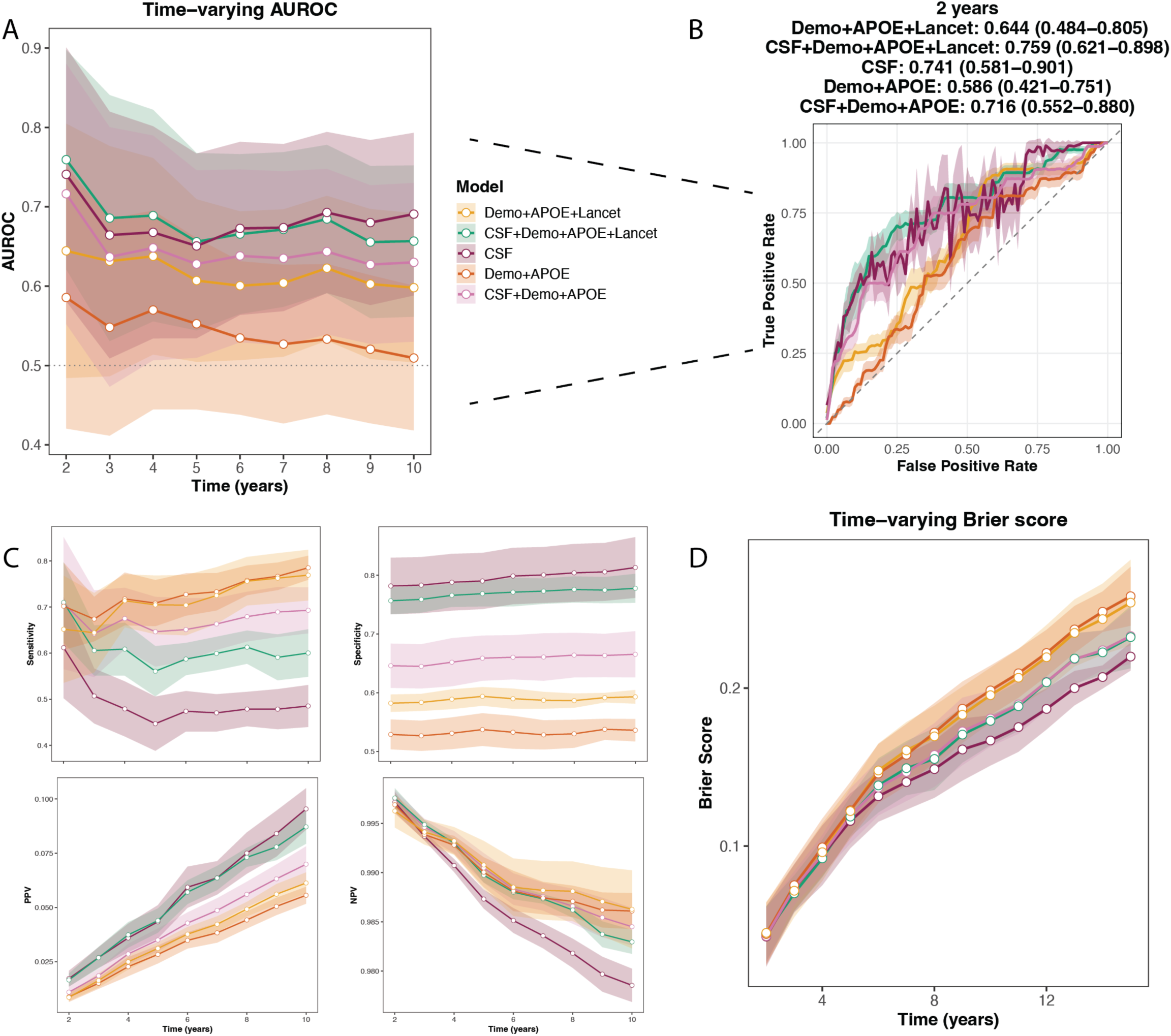
Predicting Alzheimer’s diagnosis in NACC using CSF Abeta42, pTau-181, and total tau, showing the following models: 1) Demographics+Lancet, 2) CSF+Demo+Lancet, 3) CSF, 4) Demographics, 5) CSF+Demographics. A) Time-varying AUROC for each year, with 95% confidence intervals. B) ROC curve for Year 2, which showed the largest difference in mean AUC between Demo+Lancet and CSF+Demo+Lancet. C) Sensitivity, specificity, positive predictive value, and negative predictive value for all five models using optimal cutpoints by Youden’s J statistic on the training set. D) Time-varying Brier score for each year, with standard deviations.

Demo+APOE+Lancet showed increased mean sensitivity (range: 0.011-0.072), but reduced specificity (0.082-0.098), PPV (0.009-0.024), and comparable NPV (0.0001-0.001) and Brier score (0.0027-0.0275) across time-points (Figure 4C-D).

We replicated the statistically significant effects of GDS score predicting AD in NACC as seen when predicting CDR-based clinical impairment in A4/LEARN (mean hazard ratio: 1.38 (SD: 0.057), p-value range: 2.19e-14 - 5.62e-5).

These results remained largely unchanged when using all-cause dementia as the outcome (Figure S11).

### 2.5 Plasma proteomics in the UK Biobank to predict AD ICD codes

We developed machine learning models to interrogate plasma proteomics, neuroimaging, and cognitive tests exhaustively to predict ICD-10 codes for AD. These modalities are complex and high-dimensional, providing a large possible array of potential predictors.

Using auto-machine learning, we trained LightGBM classifiers models on the UK Biobank plasma proteomics sample (N=10669; 496 cases) (Figure S12), ^32^ using combinations of proteomics (Note: this dataset does not include pTau-217 or any other form of pTau), demographics, APOE genotype, and Lancet factors.

Demographics+APOE+Lancet had a mean AUROC of 0.782 (SD: 0.045), proteomics had a mean AUROC of 0.794 (SD: 0.035), with a mean difference of 0.012 (Figure 5A). The best-performing model was the maximal Demographics+Lancet+Proteins model with feature selection with a mean AUROC of 0.835 (SD: 0.037). Examining the top 20 features for the maximal model showed a blend of plausible proteins (e.g., GFAP, BCAN, SYT1, APOE), demographics (age, APOE genotype) and Lancet factors (depression, head injury, diabetes) (Figure 5D). True positive rates were 69.71% (SD: 7.20%) for Feature-selected (FS) Demo+APOE+Proteins+Lancet and 60.56% (SD: 11.85%) for Demo+APOE+Lancet (9.15% difference), and true negative rates were 81.30% (SD: 3.92%) and 81.56% (SD: 5.38%) (0.26% difference). These results were largely similar when either removing the age filter (Figure S12), when the outcome was all-cause dementia (Figures S15, S16), and when using logistic regression models instead of LightGBM (Figures S13, S14, S17, S18).

**Fig. 5:**
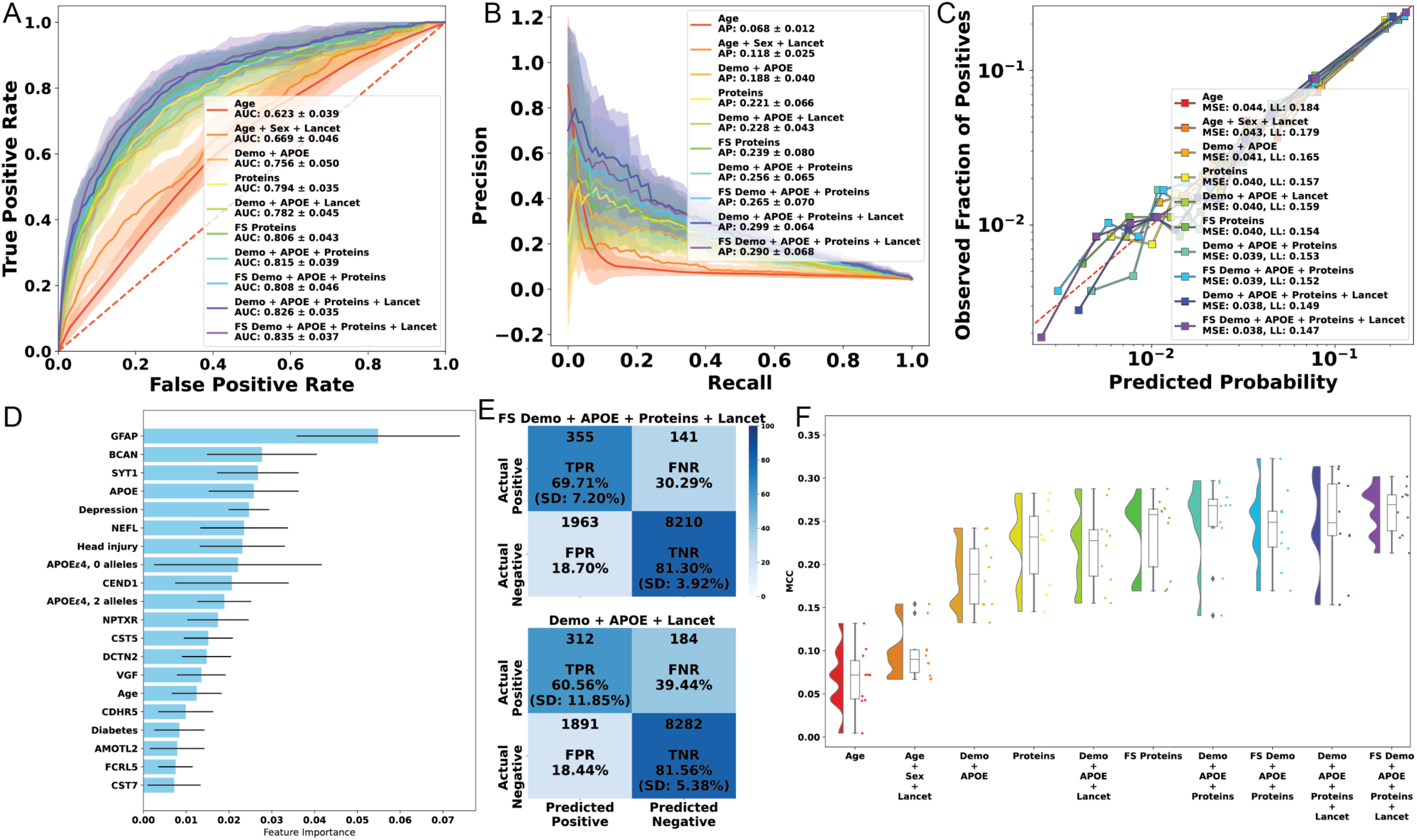
Predicting Alzheimer’s dementia with plasma proteomics, only including participants aged 65 and older. A) Mean ROC curves. B) Mean precision-recall curves. C) Calibration curves (all folds combined). D) Mean feature importance plots. E) Confusion matrices for Demo+Lancet and Feature Selected Demo+Lancet+Proteins experiments. F) Raincloud plots of Matthews correlation coefficient.

### 2.6 Brain imaging-derived phenotypes in the UK Biobank to predict AD ICD codes

The next modality included imaging-derived phenotypes (IDPs) in the UK Biobank ^33^ such as anatomical region area, thickness, and volume, structural and functional connectivity measures, and white matter integrity as predictors for AD (N=21892; 51 cases).

The Demo+APOE+Lancet model had a mean AUROC of 0.712 (SD: 0.109), and IDPs alone had a mean AUROC of 0.817 (SD: 0.096), with a mean difference of 0.105 (Figure 6A). The best-performing model was the Demo+APOE+IDPs+Lancet model with a mean AUROC of 0.855 (SD: 0.075). The top 20 features for the Demo+APOE+IDPs+Lancet model showed mostly IDPs, with the top five being volume of the subiculum-body, APOEe4 heterozygosity, mean isotropic volume fraction in the corticospinal tract, a rfMRI correlation amplitude, volume of left presubiculum-head, and another rfMRI correlation magnitude (Figure S19D). True positive rates were 49% (SD: 28.44%) for FS Demo+APOE+IDPs+Lancet and 33% (SD: 32.88%) for Demo+APOE+Lancet (16% difference), and true negative rates were 91.2% (SD: 3.51%) and 86.89% (SD: 8.38%) (4.31% difference). These results were largely similar when removing the age filter (Figure S19), when the outcome was all-cause dementia (Figures S22, S23), and when using logistic regression models instead of LightGBM (Figures S20, S21, S24, S25).

**Fig. 6:**
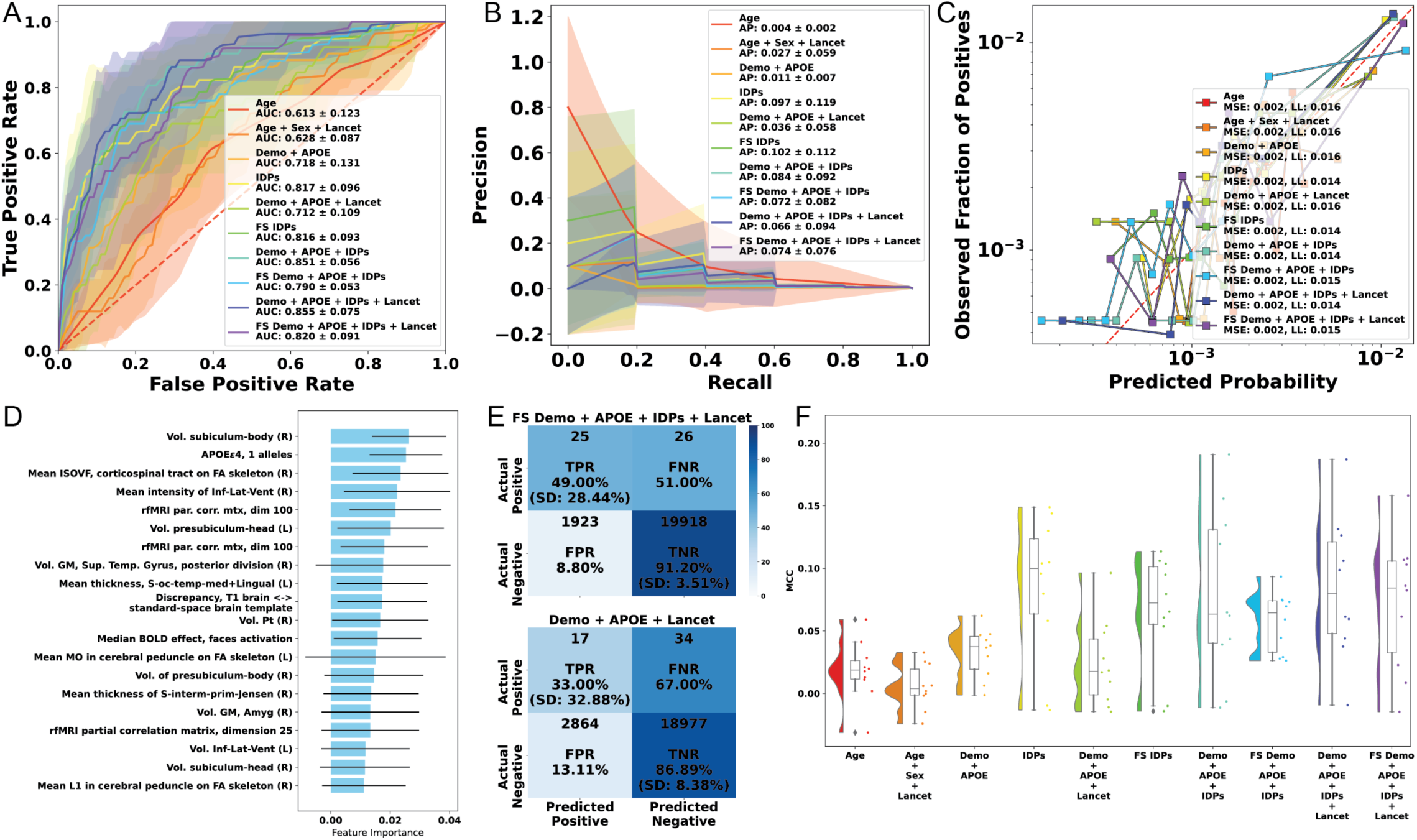
Predicting Alzheimer’s dementia with brain imaging-derived phenotypes, only including participants aged 65 and older. A) Mean ROC curves. B) Mean precision-recall curves. C) Calibration curves (all folds combined). D) Mean feature importance plots. E) Confusion matrices for Demo+Lancet and Feature Selected Demo+Lancet+IDPs experiments. F) Raincloud plots of Matthews correlation coefficient.

### 2.7 Cognitive tests in the UK Biobank to predict AD ICD codes

We interrogated measures of cognitive function in the UK Biobank ^33^, including performance on a fluid intelligence test written for the UK Biobank, matrix pattern completion, mood, numeric memory, pairs matching, symbol digit substitution, and trail making as predictors for AD/dementia (N=212448; 439 cases).

The Demo+APOE+Lancet model had a mean AUROC of 0.727 (SD: 0.05), and cognitive tests alone had a mean AUROC of 0.747 (SD: 0.085), with a mean differ-ence of 0.02 (Figure 7A). The best-performing model was Demo+APOE+Cognitive Tests with a mean AUROC of 0.799 (SD: 0.066). Examining the top 20 features for the maximal Demo+APOE+Cognitive Tests+Lancet model showed a blend of cognitive tests (symbol-digit matches, fluid intelligence score, nervous mood over the last week), demographics (Age, APOE genotype), and Lancet factors (head injury, distance to nearest major road, depression, air pollution, smoking, LDL) (Figure S26D). True positive rates were 59.17% (SD: 20.9%) for FS Demo+Cognitive Tests+Lancet and 60.57% (SD: 11.53%) for Demo+APOE+Lancet (1.4% difference), and true negative rates were 80.33% (SD: 6.43%) and 74.05% (SD: 5.19%) (6.28% difference). These results were largely similar when removing the age filter (Figure S26), when the outcome was all-cause dementia (Figures S29, S30), and when using logistic regression models instead of LightGBM (Figures S27, S28, S31, S32).

**Fig. 7:**
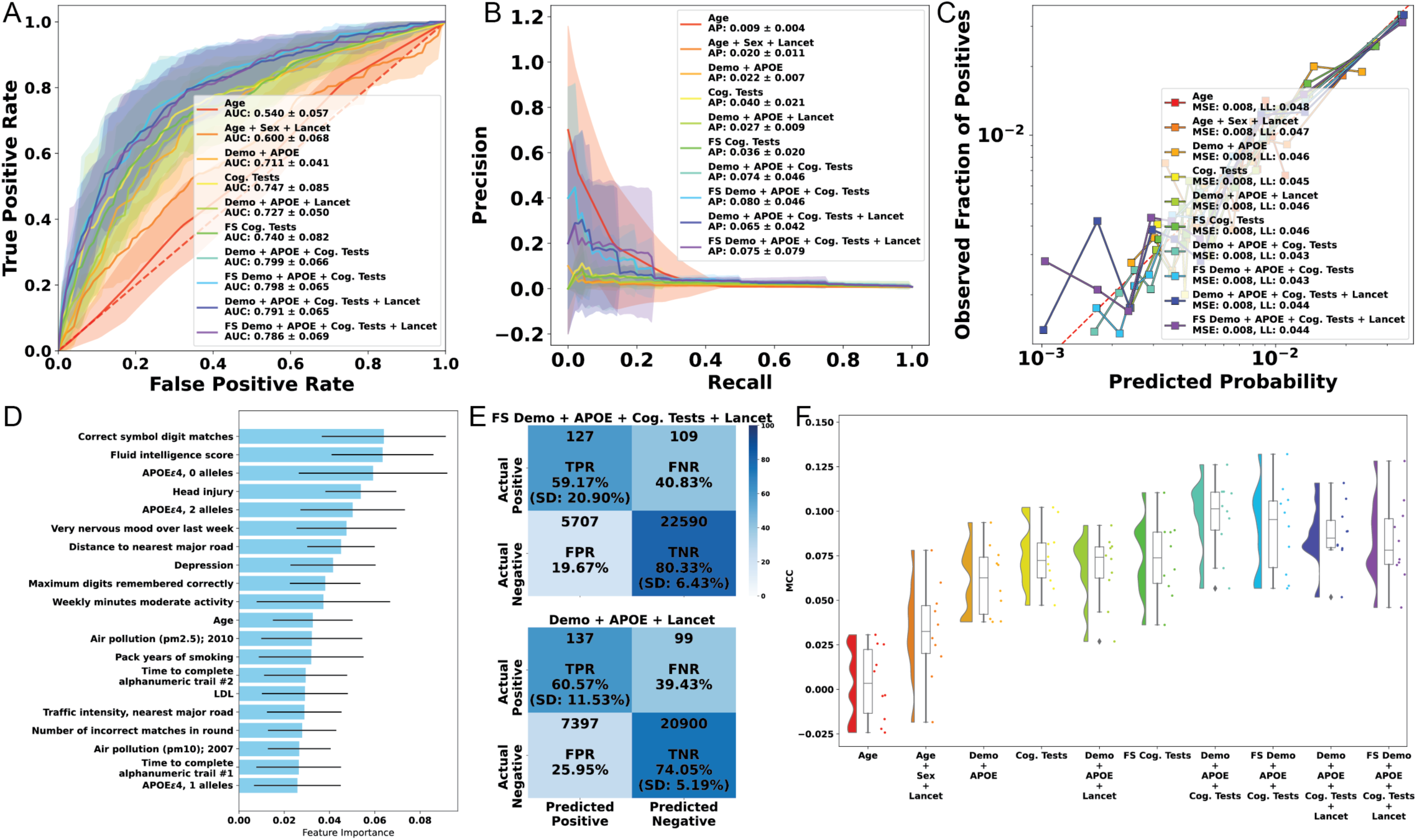
Predicting Alzheimer’s dementia with cognitive tests, only including participants aged 65 and older. A) Mean ROC curves. B) Mean precision-recall curves. C) Calibration curves (all folds combined). D) Mean feature importance plots. E) Confusion matrices for Demo+Lancet and Feature Selected Demo+Lancet+Cognitive Tests experiments. F) Raincloud plots of Matthews correlation coefficient.

### 2.8 MODAL-AD summarizes the clinical applicability of various modalities for predicting Alzheimer’s

We finally used MODAL-AD to assess our predictors of clinical impairment, clinical AD, and AD ICD codes AD predictors against clinical recommendations of acceptable biomarker performance for amyloid positivity ^25^. Briefly, Schindler and colleagues proposed minimum acceptable performance benchmarks based on amyloid prevalence and clinical context. To this end, we plotted precision-prevalence curves for each of our examined modalities and their corresponding baselines consisting of demographics, APOE genotype, and Lancet factors (Figure 8). These curves hold sensitivity and specificity constant at their empirically derived values, allowing prevalence to vary for calculating precision. This enables the comparison of these models against recommended acceptable benchmarks defined by the sensitivity and specificity of the test and prevalence of amyloid-positivity (Table 2 in ^25^: confirmatory test (90% sensitivity and specificity), high-specificity triaging test (90% sensitivity, 85% specificity), and low-specificity triaging test (90% sensitivity, 75% specificity), all examined specifically at 20% (e.g., primary care), 50 (e.g., secondary care), and 80% (e.g., dementia clinic) prevalence of amyloid-positivity).

**Fig. 8:**
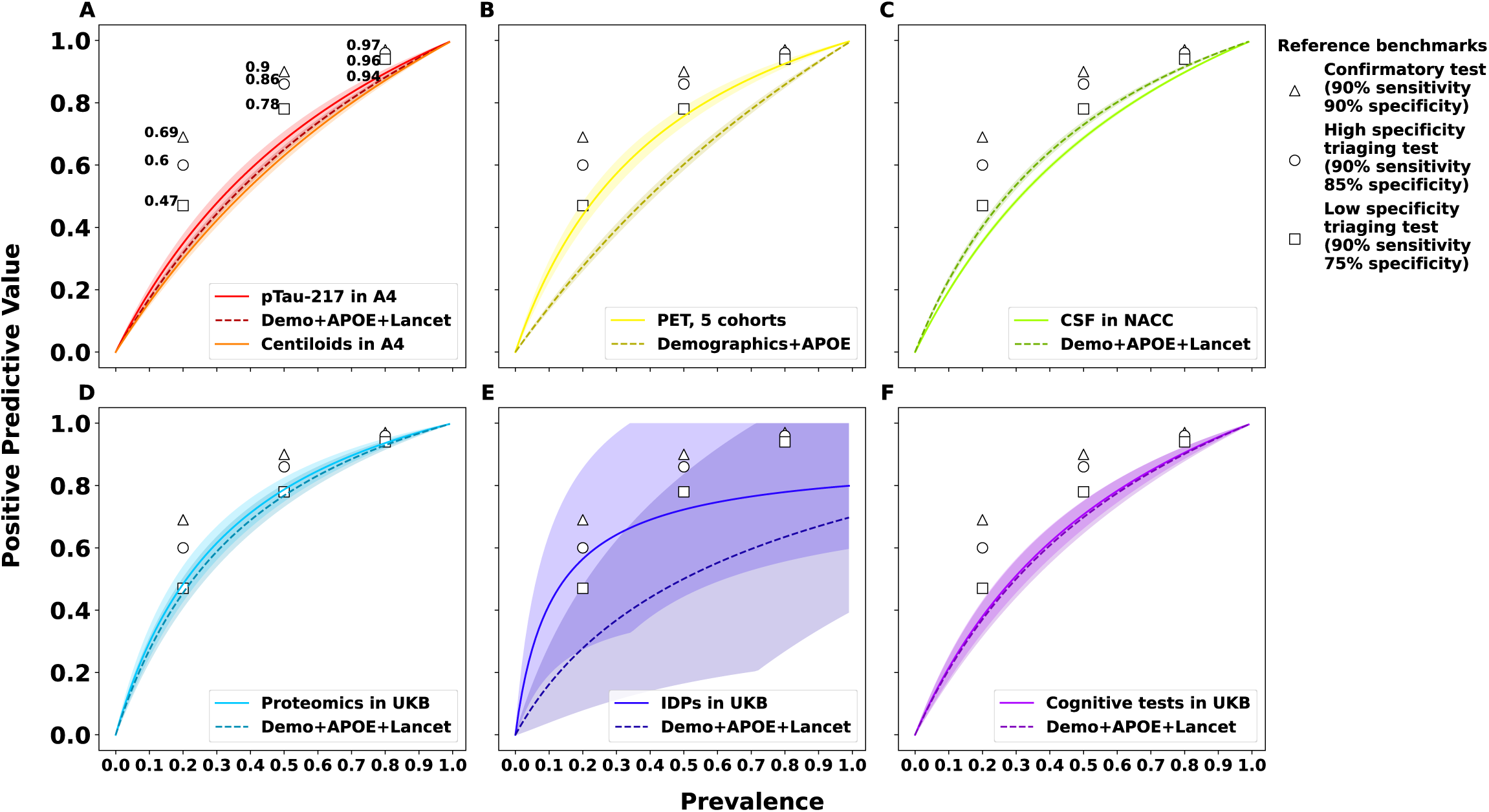
Precision-prevalence curves for all six experiments, with reference benchmarks from Table 2 in ^25^ for confirmatory, high-specificity triaging, and low-specificity triaging tests at prevalences of 20%, 50%, and 80%. Sensitivities and specificities were fixed, with prevalence allowed to vary from 0 to 1 in step sizes of 0.01 to calculate positive predictive value. A) pTau217 predicting clinical impairment in A4/LEARN, B) PET predicting AD Dx in five cohorts, C) CSF predicting AD Dx in NACC, D) Plasma proteomics predicting AD ICD codes in the UK Biobank, E) Brain imaging-derived phenotypes predicting AD ICD codes in the UK Biobank, F) Cognitive tests predicting AD ICD codes in the UK Biobank.

While the acceptable benchmarks from Schindler and colleagues are for diagnostic rather than prognostic purposes, and were developed for the outcome of amyloid positivity rather than our three outcomes, we consider this to be a valuable benchmark for the field to strive for as novel biomarkers for prognosis are developed for different clinical contexts.

## 3 Discussion

With MODAL-AD, we showcase a systematic evaluation of diverse modalities across cohorts, diagnoses, and clinical scenarios. We showed that the predictive performance of pTau-217, amyloid-PET, CSF markers, plasma proteomics, neuroimaging, and cognitive tests are marginally better than demographics and Lancet modifiable risk factors, across outcomes of clinical impairment (CDR score), and AD diagnosis and ICD codes. Thus, modifiable risk factors are a critical tool for early screening for AD that are complementary to these novel biomarkers.

It is important for future studies to 1) Compare modalities of interest to rigorous baselines of demographic and modifiable risk factors, and 2) Report comprehensive threshold-free (e.g., AUROC, AUPRC), and threshold-based (e.g., sensitivity, specificity, PPV, NPV) metrics and procedures for choosing thresholds (e.g., maximizing Youden’s J). A third issue we did not explore is data leakage ^34^, where data pre-processing or variable selection can use information from validation/test sets and inflate performance.

While many studies use demographics and APOE genotype as baseline comparators, we advocate the additional use of Lancet modifiable risk factors based on their predictive performance in the current study ^1^. Despite robust statistical associations with cognitive impairment and AD, biomarkers such as pTau-217, amyloid-PET, and CSF markers provided only marginal improvements in predictive performance when compared to simpler models incorporating these variables. Our findings also reveal a critical gap between statistical significance and practical clinical utility for these outcomes.

Our findings re-affirm the importance of evaluation of modifiable factors, such as the Lancet Modifiable Factors, as risk stratification tools in future investigations. We emphasize we did not embark on this question here. Public health data are pointing to potential population level correlates of age-dependent modification of these risk factors on gross dementia prevalence (e.g., improved education and decrease in cardiovascular risk factors such as obesity, cholesterol, diabetes, hypertension)^35^, there is growing evidence of the impact of intensive lifestyle modification (e.g., monitoring of cardiovascular risk) on slowing cognitive decline as presented in the POINTER trial ^36^. Whether these modifiable factors predict AD should be evaluated in the near future.

### 3.1 Limitations

Our study has the following limitations. First, median follow-up in our different cohorts ranged from 1.8-13.72 years, and some patients not labeled as AD may incur it after our the end of follow-up. Second, additional cohorts and increased sample sizes may yield improved and more precise estimates of predictive performance. Third, the outcome in our PET analyses was clinical AD diagnosis, but these cohorts adjudicated this in slightly different ways based on a combination of biomarkers and cognitive measures (e.g., MMSE, CDR) with uncertain accuracy for neuropathological changes ^37^ or contrasting to other forms of dementia. Fourth, the outcome in our UK Biobank analyses was AD ICD codes, which may not be as accurate as clinical adjudication, although their precision has been shown to be fairly strong ^32^. Fifth, the translational value of these datasets (A4/LEARN being clinical trials, ADNI, AIBL, HABS, NACC, and OASIS being cohort studies, and the UK Biobank being a volunteer-based biobank) to primary care is an open question in terms of prevalence impacting PPV and NPV, along with follow-up and detail of clinical history.

Looking forward, we advocate for a standardized and transparent approach in dementia prediction research. Future studies should consistently report multiple predictive metrics, and employ rigorous baseline comparators to fully evaluate the evidence in a unbiased manner. Moreover, integrating modifiable risk factors into electronic health records as screening tools, alongside advanced biomarkers, could offer a more holistic, scalable, low-cost predictive approach that aligns closely with patient management strategies in primary care.

## 4 Methods

We developed MODAL-AD Table 1 to systematically compare the performance of AD biomarkers with demographics and modifiable risk factors in cognitively asymptomatic patients 65 and older.

**Table 1:** Summary of cohorts, outcomes, and data sources in the different analysis. In the amyloid-predictive modalities (i.e., pTau-217, amyloid-PET, CSF), time-to-event analysis with time-varying covariates was used. In the discovery modalities (proteomics, multimodal neuroimaging, and cognitive tests in the UK Biobank), we used binary classification with auto-machine learning. Lancet factors were not used in the Amyloid-PET experiments and marked with ”N/A” because across the five included cohorts, these variables were not uniformly measured. All patients were cognitively unimpaired at baseline. The three different outcomes were: 1) Clinical impairment (A4/LEARN), defined as a Clinical Dementia Rating score of 0.5 or greater on two consecutive visits, 2) AD Diagnosis adjudicated by clinicians in the designated cohorts, or 3) AD ICD codes from electronic health records in the UK Biobank. In each cohort, ”controls” refer to patients who do not meet the outcome at any time point, and ”cases” refer to patients who progress to the outcome during follow-up. Follow-up times are presented as medians and interquartile ranges calculated using reverse Kaplan-Meier analysis. A4: Anti-Amyloid Treatment in Asymptomatic Alzheimer’s. LEARN: Longitudinal Evaluation of Amyloid Risk and Neurodegeneration. ADNI: Alzheimer’s Disease Neuroimaging Initiative. AIBL: Australian Imaging, Biomarker & Lifestyle. NACC: National Alzheimer’s Coordinating Center. OASIS: Open Access Series of Imaging Studies. CSF: cerebrospinal fluid. IDPs: Imaging-derived phenotypes. ICD: International Classification of Diseases. GDTotal: Geriatric Depression Scale total score. STAITotal: State-Trait Anxiety Inventory for Adults total score.

|  | A4/LEARN | ADNI | AIBL | HABS | NACC | OASIS | UK Biobank |
| --- | --- | --- | --- | --- | --- | --- | --- |
| **Data** | pTau-217, PET (1031 Controls, 409 Cases) | pTau-217 (325 Controls, 31 Cases); Amyloid-PET (561 Controls, 171 Cases) | Amyloid-PET (393 Controls, 26 Cases) | Amyloid-PET (279 Controls, 0 Cases) | Amyloid-PET (342 Controls, 29 Cases); CSF markers (AB42, pTau-181, Total tau) (759 Controls, 519 Cases) | Amyloid-PET (509 Controls, 44 Cases) | Plasma proteomics (10,173 Controls, 496 Cases); Multimodal neuroimaging (21,841 Controls, 51 Cases); Cognitive tests (28,297 Controls, 236 Cases) |
| **Outcome** | CDR-Global Score 0.5+ on two consecutive visits | AD Diagnosis | AD Diagnosis | AD Diagnosis | AD Diagnosis | AD Diagnosis | AD ICD codes |
| **Lancet Factors** | Smoking, alcohol, substance use, aerobic activity, walking, GDTotol, STAITotol, diastolic + systolic BP | Smoking, alcohol, head- /eyes/ears/nose/throat issues, visual/auditory impairment, LDL, hypertension, BMI, GDTotol; N/A | N/A | N/A | N/A; TBI, smoking, alcohol abuse, hypertension, BMI, diabetes, hearing, depression, GDTotol | N/A | Hearing loss, vision loss, LDL cholesterol, depression, TBI, physical inactivity, diabetes, smoking, hypertension, obesity, alcohol, social isolation, air pollution |
| **Age** | 65–86 | 65–99; 65–98 | 65–96 | 65–92 | 65–95; 67–100 | 65–92 | 65–70; 65–82; 65–72 |
| **Sex** |  |  |  |  |  |  |  |
| Female | 856 | 197; 398 | 221 | 165 | 250; 725 | 320 | 5,346; 10,606; 14,019 |
| Male | 584 | 159; 334 | 198 | 114 | 121; 553 | 233 | 5,323; 11,286; 14,514 |
| **Follow-up (years)** | 6.78 (5.55-7.53) | 4.71 (2.28-6.44); 4.25 (2.19-8.08) | 1.8 (0.26-5.33) | 5.15 (4.43-5.4) | 3.98 (1.24-8.53); 9.05 (5-13.96) | 7.11 (3.31-12.11) | 13.72 (13.09-14.42); 4.29 (3.46-5.62); 13.54 (12.92-14.23) |

An overview of MODAL-AD is described in Table 1. In brief, MODAL-AD iterates through possible predictor modalities and compares the predictions with a a priori chosen suite of risk factors. A *modality* is defined as a class or combination of potential predictors, such as pTau, proteins, congitive tests, or genetic factors (to name a few). We used all available participants from the included datasets who met the eligibility and exclusion criteria. Study size was determined by data availability rather than a priori sample size calculation. This approach is standard in secondary analyses and model development studies, where maximizing the number of eligible participants helps ensure stable model estimation, improve generalizability, and reduce overfitting.

A study protocol was not prepared, and the study was not registered. Code is available at www.github.com/randalljellis/rfb

For both the model development and evaluation phases, all eligible samples were included and partitioned into training and testing subsets (details provided in the data splitting section). Formal power or sample size calculations were not performed, as these are not directly applicable to complex AI-based models trained on observational data; instead, adequacy of sample size was evaluated empirically through model performance stability, cross-validation, and uncertainty quantification.

These modalities are evaluated in plausible clinical scenarios. We describe the deployment of each clinical scenario in turn, below:

### 4.1 pTau-217 and predicting clinical impairment in A4/LEARN

The Anti-Amyloid Treatment in unimpaired Alzheimer’s (A4) Study (2014-2022) was a multisite, longitudinal clinical trial S7 that tested whether amyloid reduction in asymptomatic older adults with elevated brain amyloid levels would slow cognitive decline ^38^. Participants were cognitively unimpaired at baseline but had elevated amyloid levels as assessed by ^18^F-florbetapir PET.

The Longitudinal Evaluation of Amyloid Risk and Neurodegeneration (LEARN) study (2015-2023) enrolled amyloid-negative individuals, without intervention. A4 and LEARN participants underwent longitudinal clinical, cognitive, and neuroimaging evaluations.

We used the A4/LEARN cohorts to assess pTau-217 and amyloid-PET for prediction of clinical impairment, defined by two consecutive visits with a Clinical Dementia Rating (CDR) score of 0.5 or greater as used in prior work on A4/LEARN ^28,38^.

Lancet factors in A4 included smoking, alcohol, substance use, aerobic activity, walking, depression, anxiety, and blood pressure.

We conducted a subgroup analysis of patients who were amyloid-positive at baseline, and examined a different outcome (Preclinical Alzheimer Cognitive Composite (PACC)) to evaluate the robustness of our results.

For each fold in a 5-fold cross-validation (CV) procedure, stratified by quantile distribution of times to censoring or experiencing an event, we fit 20 time-varying Cox models: Demographics (i.e., age, sex, education), Demographics+APOE genotype, Demographics+Lancet factors, Demographics+APOE+Lancet, Lancet factors alone, pTau-217 alone, pTau-217+Demographics, pTau-217+Demographics+APOE, pTau-217+Demographics+Lancet factors+APOE, pTau-217+Demographics+Lancet factors, PET alone, PET+Demographics+APOE, PET+Demographics, PET+Demographics+Lancet+APOE, PET+Demographics+Lancet, pTau-217+PET, pTau-217+PET+Demographics+APOE, pTau-217+PET+Demographics, pTau-217+PET+Demographics+Lancet+APOE, pTau-217+PET+Demographics+Lancet. We predicted at Years 3-7 for a total of 25 analytical ”scenarios” (5 CV folds x 5 time points; multiple testing correction was used when comparing models ^31^). The timeROC package accounts for multiple testing when comparing time-dependent AUCs at several time points by modeling the joint distribution of the test statistics as multivariate normal and adjusting p-values accordingly. Specifically, it estimates the covariance matrix of the standardized AUC differences and computes asymptotically exact adjusted p-values using the joint multivariate normal distribution of these statistics ^31^.

PTau-217 data were transformed using the box-cox transformation to make the data normally distributed, with the lambda parameter chosen using the training folds, and applied to the validation folds to prevent data leakage.

Demographics included age, age-squared, sex, years of education, APOE genotype, and interaction terms between APOE genotype and the two age variables. Lancet factors in A4 included smoking (average cigarette packs per day), alcohol (average drinks per day), substance use (binary of whether participant used other substances), aerobic activity (average number of hours of aerobic exercise per week), walking (average number of minutes walking per day), GDTotal (Geriatric Depression Scale total score), STAITotal (State-Trait Anxiety Inventory for Adults total score), diastolic and systolic blood pressure.

### 4.2 Cross-cohort amyloid-PET and AD prediction with MODAL-AD

We used amyloid-PET data from the Australian Imaging Biomarkers & Lifestyle (AIBL), Alzheimer’s Disease Neuroimaging Initiative (ADNI), Harvard Aging Brain Study (HABS), National Alzheimer’s Coordinating Center (NACC), and Open Access Series of Imaging Studies (OASIS) to predict AD diagnosis at Years 2-10. The criteria and methodology for AD diagnosis in each cohort is described in the Supplemental Methods. As Lancet factors were not uniformly measured across cohorts, we only modeled demographics and APOE genotype, and then assessed the added value of Amyloid-PET as the only biomarker.

#### 4.2.1 AIBL

AIBL is a two-site longitudinal cohort designed to identify biomarkers, cognitive and lifestyle factors associated with AD ^39^ that enrolled in 2006-2008 and additional patients from 2009-2019, and following up to present every 18 months. Amyloid-PET imaging was conducted primarily using ^11^C-PiB or ^18^F-florbetaben. For this study, we included participants with at least one amyloid-PET scan.

AD diagnosis was determined according to the National Institute of Neurological and Communicative Disorders and Stroke and the Alzheimer’s Disease and Related Disorders Association (NINCDS-ADRDA) Criteria, as well as Diagnostic and Statistical Manual (DSM)-IV and International Classification of Diseases (ICD)-10 diagnostic criteria ^39^.

Data used in the preparation of this article was obtained from the Australian Imaging Biomarkers and Lifestyle flagship study of ageing (AIBL). See www.aibl.csiro.au for further details.

Data was collected by the AIBL study group. AIBL study methodology has been reported previously ^39^.

#### 4.2.2 ADNI

ADNI is a multicenter, longitudinal study launched in 2004 and still enrolling patients to develop clinical, imaging, genetic, and biochemical biomarkers for the early detection and tracking of AD ^40^ from about 60 sites. We used data from ADNI-1, GO, 2, and 3 phases. Participants underwent amyloid-PET imaging with either ^11^C-PiB, ^18^F-florbetapir, or ^18^F-florbetaben.

Lancet factors in ADNI used in the pTau-217 analyses included smoking (average cigarette packs per day, years of smoking, time since quitting smoking, and current smoking status), alcohol (average drinks per day, years of abuse, time since quitting alcohol abuse, current alcohol status), head/eyes/ears/nose/throat issues (binary), cardiovascular history (binary), low density lipoprotein, history of hypertension (binary), GDTotal (Geriatric Depression Scale total score), visual impairment (binary), auditory impairment (binary).

AD diagnosis was determined by ^41^ a sequence beginning with memory complaints, followed by a Mini-Mental State Examination score 20-26 (inclusive), a Clinical Dementia Rating score of 0.5 or 1, and delayed recall of 1 paragraph from the Logical Memory II subscale of the Wechsler Memory Scale-Revised with cutoffs based on education: *↑* 8 for 16 years of education, *↑* 4 for 8-15 years of education, and *↑* 2 for 0-7 years of education. Additionally, AD subjects had to meet the NINCDS-ADRDA criteria for probable AD.

Data used in the preparation of this article were obtained from the Alzheimer’s Disease Neuroimaging Initiative (ADNI) database (adni.loni.usc.edu). The ADNI was launched in 2003 as a public-private partnership, led by Principal Investigator Michael W. Weiner, MD. The original goal of ADNI was to test whether serial magnetic resonance imaging (MRI), positron emission tomography (PET), other biological markers, and clinical and neuropsychological assessment can be combined to measure the progression of mild cognitive impairment (MCI) and early Alzheimer’s disease (AD). The current goals include validating biomarkers for clinical trials, improving the generalizability of ADNI data by increasing diversity in the participant cohort, and to provide data concerning the diagnosis and progression of Alzheimer’s disease to the scientific community. For up-to-date information, see adni.loni.usc.edu.

#### 4.2.3 HABS

The Harvard Aging Brain Study (HABS; 2010-2017) is a one-site prospective observational cohort focused on identifying early biomarkers of AD among cognitively asymptomatic older adults ^42^. All participants were clinically healthy at enrollment and underwent baseline amyloid-PET imaging using ^11^C-PiB.

Diagnoses of cognitive impairment or dementia ^42^ were assessed during follow-up based on a consensus review of neuropsychological trajectories and clinical assessments. For this analysis, we included all participants with a baseline PET scan and at least one follow-up cognitive or diagnostic evaluation.

#### 4.2.4 NACC

The National Alzheimer’s Coordinating Center (NACC; 2004-present) maintains a large, harmonized dataset compiled from 36 Alzheimer’s Disease Research Centers (ADRCs) across the United States ^43^. While not all NACC participants undergo PET imaging, we included the subset for whom amyloid-PET data were available, typically uploaded by individual centers.

Lancet factors used when predicting with cerebrospinal fluid markers were: history of TBI (binary), TBI (absent, recent/active, remote/inactive), TBI with brief loss of consciousness (none, single, repeated), TBI without loss of consciousness (none, single, repeated), smoking (smoked in last 30 days, more than 100 cigarettes in life, age at quitting), alcohol (consumption frequency in last 3 months (less than once per month, once per month, once per week, a few times a week, daily or almost daily), ”clinically significant impairment occurring over a 12-month period manifested in one of the following areas: work, driving, legal, or social” (absent, recent/active, remote/inactive), current abuse (binary)), hypertension (currently present (binary), history (absent, recent/active, remote/inactive), history or present (binary)), current use of an antihypertensive or blood pressure medication (binary), current use of an antihypertensive combination therapy (binary), diabetes (currently present (binary), history (absent, recent/active, remote/inactive)), current use of a diabetes medication (binary), functional hearing without a hearing aid (binary), functional hearing with a hearing aid (binary), using a hearing aid (binary), depression or dysphoria in the last month (binary), depression or dysphoria severity (mild, moderate, severe), current use of an antidepressant (binary), depression treated or untreated (binary), active depression in last two years (binary), depression episodes more than two years ago (binary).

Diagnosis of dementia and its etiology (e.g., AD, vascular dementia) were determined according to the NACC Uniform Data Set Coding Guidebook ^44^ Form D1b (Etiological Diagnosis and Biomarker Support) completed by clinicians. Section 1 (Biomarkers and imaging) of this form has choices of ”No, inconsistent,” ”Yes, consistent,” ”Indeterminate,” and ”Not assessed” for fluid (blood, CSF) and imaging (PET, single-photon emission computed tomography (SPECT), magnetic resonance imaging (MRI), computed tomography (CT)) biomarkers. Section 2 (Etiological diagnoses), pertaining to AD, incorporates the 2011 NIA-AA criteria as well as specific criteria for probable versus possible AD. Probable includes 1) meeting criteria for dementia, and exhibiting 2) ”Insidious onset,” 3) ”Clear-cut history of worsening of cognition,” and 4) ”initial and most prominent cognitive deficits” belonging to the categories of amnestic disorder or non-amnestic disorder (i.e., language, visuospatial, executive and behavioral disorder). Exclusions for probable AD include ”substantial concomitant cerebrovascular disease” and exhibiting features of dementia with Lewy bodies, frontotemporal dementia, semantic variant primary progressive aphasia or non-fluent/agrammatic variant primary progressive aphasia or ”another concurrent, active neurological disease, or a non-neurological medical co-morbidity or medication use that could have a substantial impact on cognition.” Possible AD describes patients meeting one of two criterion: 1) Atypical course meeting criteria (1) or (4) for probable AD, but with ”sudden onset of cognitive impairment or demonstrates insufficient historical detail or objective cognitive documentation of progressive decline,” or 2) ”Etiologically mixed presentation” meeting (1) *through* (4) above for probable AD but with evidence of concomitant cerebrovascular disease, features of dementia with Lewy bodies other than the dementia itself, or ”evidence for another neurological disease or a non-neurological medical co-morbidity or medication use that could have a substantial impact on cognition.”

#### 4.2.5 OASIS

The Open Access Series of Imaging Studies (OASIS) is a publicly available cross-sectional and longitudinal neuroimaging dataset from older adults ^45^ collected from 2004-2019 at one site. We used OASIS-3, which includes ^11^C-PiB amyloid-PET scans and longitudinal clinical evaluations. Dementia status and etiology were determined based on Clinical Dementia Rating (CDR) scores and clinical consensus.

AD/dementia diagnosis was determined ^45^ using the same NACC Uniform Data Set, as well as CDR score, where 0.5 indicated ”very mild impairment,” 1 was ”mild impairment,” and 2 was ”moderate dementia.” During these assessments, ”clinicians completed a diagnostic impression intake and interview culminating with a coded dementia diagnosis that is recorded in the OASIS datatype ’ADRC Clinical Data’ dx1-dx5. Diagnoses for this variable include ’cognitively normal’, ’AD dementia’, ’vascular dementia’ and contributing factors such as vitamin deficiency, alcoholism, and mood disorders. The diagnostic determination for variables dx1-dx5 is separate from UDS assessments, however they may overlap in diagnostic conclusions.”

### 4.3 Cerebrospinal fluid predicting AD in NACC

A subset of participants in the NACC cohort contributed cerebrospinal fluid (CSF) samples (including amyloid-beta_42_, total tau (t-tau), and phosphorylated tau-181 (pTau-181), collected according to respective ADRC protocols ^43^. We modeled combinations of demographics, APOE genotype, Lancet factors (TBI, smoking, alcohol consumption, hypertension, BMI, diabetes, hearing, depression), and CSF markers.

A subset of participants in the NACC cohort contributed cerebrospinal fluid (CSF) samples, collected and assayed according to protocols established by their affiliated Alzheimer’s Disease Research Centers (ADRCs). The availability and assay methods for CSF biomarkers (including amyloid-beta_42_, total tau (t-tau), and phosphorylated tau-181 (pTau-181) varied across centers and over time ^43^. We fit 10 models: Demographics, Demographics w/o APOE genotype, Demo+Lancet w/o APOE genotype, Demo+Lancet, Lancet alone, CSF+Demo w/o APOE genotype, CSF+Demo, CSF+Demo+Lancet w/o APOE genotype, CSF+Demo+Lancet, CSF alone.

### 4.4 Time-varying longitudinal modeling with MODAL-AD

For amyloid-predictive modalities (i.e., pTau-217, amyloid-PET, CSF), we used the survival package in R ^46^ to conduct time-to-event analyses with time-varying covariates. Here, predictors may change over time, and performance is assessed over follow-up time. In a cross-validation (CV) procedure stratified by control/case status with an 80/20 split, we predicted AD diagnosis at annual timepoints from the start of the study, for a total number of analytical ”scenarios” (i.e., 5 CV-folds multiplied by the number of timepoints).

We compared time-varying AUROC values between models, and computed confidence intervals and p-values using timeROC in R ^31^. For each model, decision thresholds maximizing Youden’s J statistic ^47^ were identified from the training set and used to calculate sensitivity, specificity, positive predictive value, and negative predictive value on the validation set.

Missing data were handled in survival models by using the last value carried forward and next observation carried backward, but only within the follow-up window.

No data were ever imputed post-censoring time. In classification models no missing data handling was needed because LightGBM natively handles missing data. Patients with no values of age, sex, education, or APOE genotype were excluded.

### 4.5 MODAL-AD implementation across high-dimensional modalities in the UK Biobank

The UK Biobank is a volunteer-based biobank based in the United Kingdom with approximately 500,000 participants. We used the proteomics (collected 2006-2010), neuroimaging (2014-2022), and cognitive tests (2006-2010) from 22 assessment centers in the the UK Biobank ^33^ (Table 1). The proteomics dataset consisted of 10,669 participants (496 AD cases), the neuroimaging dataset consisted of 21,892 participants (51 AD cases), and the cognitive tests dataset consisted of 28,533 participants (236 AD cases).

Aside from these advanced modalities, we also used demographics (age, sex, education, APOE genotype) for prediction, as well as Lancet-identified modifiable risk factors when available. The 14 Lancet modifiable risk factors are: education (age at education completed; already included in demographics), hearing loss (current (binary), length suffering (*<* 2 weeks, 2-4 weeks, 4-12 weeks, *>*12 weeks), extent affected (binary), LDL cholesterol, depression (episode (binary), recurrent (binary)), traumatic brain injury (ICD codes S00-S09), physical inactivity (metabolic equivalent of task (MET) minutes per week of moderate activity), diabetes (insulin-dependent, non-insulin-dependent, malnutrition-related, other, unspecified, during pregnancy; all binary), smoking (pack years), hypertension (primary, secondary, gestational with or without proteinuria, pre-existing hypertension complicating pregnancy, childbirth and the puerperium; all binary)), obesity (binary), excessive alcohol consumption, social isolation (frequency of friends/family visit (no friends/family outside household, never or almost never, once every few months, about once a month, about once a week, 2-4 times a week, almost dailty), loneliness/isolation (binary)), air pollution (inverse distance to nearest major road, nitrogen dioxide air pollution, particulate matter air pollution (pm10 and pm2.5), sum of read length of major roads within 100m, traffic intensity on nearest major road; all continuous), and vision loss (age cataract diagnosed, age when diabetes-related eye disease diagnosed).

We used the autoML framework FLAML ^48^ for hyperparameter optimization of a LightGBM classifier ^49^. We used the LightGBM classifier because of its speed when fitting to high-dimensional datasets.

For each of the three modalities, we ran ten models: 1) Age alone, 2) Age, sex, Lancet factors, 3) Demographics (Age, sex, ethnicity, APOE*ω*4 alleles, last completed education, age when full-time education was completed), 4) Modality (proteins, neuroimaging, or cognitive tests), 5) Demographics and Lancet factors, 6) Feature selection of modality, 7) Demographics and modality, 8) Feature selection of demographics and modality, 9) Demographics, modality, and Lancet factors, and 10) Feature selection of demographics, modality, and Lancet factors. For experiments using feature selection, we calculated feature importance on the trained models from experiments (4), (7), and (9), and iteratively retrained and predicted on the top *k* features, for values of *k* 1-100.

For each experiment, we used a 10-fold geographical cross-validation approach to estimate the uncertainty of our performance metrics. Grouping the UK Biobank assessment centers into 10 geographical regions (in the same fashion as ^50^), we performed autoML on each fold consisting of a training set of 9/10 geographical regions, and predicting on the one held out geographical region, with this procedure performed separately for all 10 folds. For the neuroimaging dataset, only four geographical regions were represented so we instead used a 10-fold stratified cross-validation procedure.

On the test set in each fold, we calculated two threshold-free metrics: the area under the receiver operating characteristic curve, and the average precision of the precision-recall curve. Then, we chose the decision threshold that maximized Youden’s J statistic, and used this threshold to calculate confusion matrices, the true positive rate (TPR; also referred to as ”sensitivity” or ”recall”), false positive rate (FPR), true negative rate (TNR; also referred to as ”specificity”), false negative rate (FNR), positive predictive value (PPV; also referred to as ”precision”), and the Matthews correlation coefficient (MCC). We also calculated feature importance for all experiments/folds (with the exception of experiments with Age alone), along with calibration curves. All metrics and feature importance values were averaged across the 10 cross-validation folds and reported as means and standard deviations.

## Data Availability

Data are available from their respective sources upon access being granted after application.

## Acknowledgements

RE and CJP conceived of the study and wrote the initial draft. RE developed the analytics pipeline (MODAL-AD) and attained the A4/LEARN cohort study data. AA provided analytics support. VJ, MB, SK, VBK provided cohort data, VBK edited the initial draft. MMG, CD, and DAB edited the initial draft. The authors have no conflicts of interest to disclose.

The authors thank Aaron Kesselheim, Randy Buckner, Lee Rubin, Rachel Buckley, Zahra Shirzadi, Jasmeer Chhatwal, and Wai-Ying Wau for their comments.

This study was supported in part by NIEHS R01ES032470, NIA RF1AG074372, the Vranos Foundation,and NLM BIRT T32 Training Fellowship T15LM007092.

The A4 Study was a secondary prevention trial in preclinical Alzheimer’s disease, aiming to slow cognitive decline associated with brain amyloid accumulation in clinically normal older individuals. The A4 Study was funded by a public-private-philanthropic partnership, including funding from the National Institutes of Health-National Institute on Aging, Eli Lilly and Company, Alzheimer’s Association, Accelerating Medicines Partnership, GHR Foundation, an anonymous foundation, and additional private donors, with in-kind support from Avid Radiopharmaceuticals, Cogstate, Albert Einstein College of Medicine and the Foundation for Neurologic Diseases.The companion observational Longitudinal Evaluation of Amyloid Risk and Neurodegeneration (LEARN) Study was funded by the Alzheimer’s Association and GHR Foundation. The A4 and LEARN Studies were led by Dr. Reisa Sperling at Brigham and Women’s Hospital, Harvard Medical School, and Dr. Paul Aisen at the Alzheimer’s Therapeutic Research Institute (ATRI) at the University of Southern California. The A4 and LEARN Studies were coordinated by ATRI at the University of Southern California, and the data are made available under the auspices of Alzheimer’s Clinical Trial Consortium through the Global Research & Imaging Platform (GRIP).

The complete A4 Study Team list is available on: https://www.actcinfo.org/a4-study-team-lists/. We would like to acknowledge the dedication of the study participants and their study partners who made the A4 and LEARN Studies possible.

Data collection and sharing for the Alzheimer’s Disease Neuroimaging Initiative (ADNI) is funded by the National Institute on Aging (National Institutes of Health Grant U19AG024904). The grantee organization is the Northern California Institute for Research and Education. In the past, ADNI has also received funding from the National Institute of Biomedical Imaging and Bioengineering, the Canadian Institutes of Health Research, and private sector contributions through the Foundation for the National Institutes of Health (FNIH) including generous contributions from the following: AbbVie, Alzheimer’s Association; Alzheimer’s Drug Discovery Foundation; Araclon Biotech; BioClinica, Inc.; Biogen; Bristol-Myers Squibb Company; CereSpir, Inc.; Cogstate; Eisai Inc.; Elan Pharmaceuticals, Inc.; Eli Lilly and Company; EuroImmun; F. Hoffmann-La Roche Ltd and its affiliated company Genentech, Inc.; Fujirebio; GE Healthcare; IXICO Ltd.; Janssen Alzheimer Immunotherapy Research & Development, LLC.; Johnson & Johnson Pharmaceutical Research & Development LLC.; Lumosity; Lundbeck; Merck & Co., Inc.; Meso Scale Diagnostics, LLC.; NeuroRx Research; Neurotrack Technologies; Novartis Pharmaceuticals Corporation; Pfizer Inc.; Piramal Imaging; Servier; Takeda Pharmaceutical Company; and Transition Therapeutics.

Data used in the preparation of this article were obtained from the Harvard Aging Brain Study (Data Release 2.00; P01AG036694; https://habs.mgh.harvard.edu). The HABS study was launched in 2010, funded by the National Institute on Aging. and is led by principal investigators Reisa A. Sperling MD and Keith A. Johnson MD at Massachusetts General Hospital/Harvard Medical School in Boston, MA. We obtained the data in August 2022 from synapse.org.

The NACC database is funded by NIA/NIH Grant U24 AG072122. NACC data are contributed by the NIA -funded ADRCs: P30 AG062429 (PI James Brewer, MD, PhD), P30 AG066468 (PI Oscar Lopez, MD), P30 AG062421 (PI Bradley Hyman, MD, PhD), P30 AG066509 (PI Thomas Grabowsk i, MD), P30 AG066514 (PI Mary Sano, PhD), P30 AG066530 (PI Helena Chui, MD), P30 AG066507 (PI Marilyn Albert, PhD), P30 AG066444 (PI John Morris, MD), P30 AG066518 (PI Jeffrey Kaye, MD), P30 AG066512 (PI Thomas Wisniewski, MD), P30 AG066462 (PI Scott Small, MD), P30 AG072979 (PI David Wolk, MD), P30 AG072972 (PI Charles DeCarli, MD), P30 AG072976 (PI Andrew Saykin, PsyD), P30 AG072975 (PI David Bennett, MD), P30 AG072978 (PI Neil Kowall, MD), P30 AG072977 (PI Robert Vassar, PhD), P30 AG066519 (PI Frank LaFerla, PhD), P30 AG062677 (PI Ronald Petersen, MD, PhD), P30 AG079280 (PI Eric Reiman, MD), P30 AG062422 (PI Gil Rabinovici, MD), P30 AG066511 (PI Allan Levey, MD, PhD), P30 AG072946 (PI Linda Van Eldik, PhD), P30 AG062715 (PI Sanjay Asthana, MD, FRCP), P30 AG072973 (PI Russell Swerdlow, MD), P30 AG066506 (PI Todd Golde, MD, PhD), P30 AG066508 (PI Stephen Strittmatter, MD, PhD), P30 AG066515 (PI Victor Henderson, MD, MS), P30 AG072947 (PI Suzanne Craft, PhD), P30 AG072931 (PI Henry Paulson, MD, PhD), P30 AG066546 (PI Sudha Seshadri, MD), P20 AG068024 (PI Erik Roberson, MD, PhD), P20 AG068053 (PI Justin Miller, PhD), P20 AG068077 (PI Gary Rosenberg, MD), P20 AG068082 (PI Angela Jefferson, PhD), P30 AG072958 (PI Heather Whitson, MD), P30 AG072959 (PI James Leverenz, MD).

The NACC database is funded by NIA/NIH Grant U24 AG072122. SCAN is a multi-institutional project that was funded as a U24 grant (AG067418) by the National Institute on Aging in May 2020. Data collected by SCAN and shared by NACC are contributed by the NIA-funded ADRCs as follows: Arizona Alzheimer’s Center - P30 AG072980 (PI: Eric Reiman, MD); R01 AG069453 (PI: Eric Reiman (contact), MD); P30 AG019610 (PI: Eric Reiman, MD); and the State of Arizona which provided additional funding supporting our center; Boston University - P30 AG013846 (PI Neil Kowall MD); Cleveland ADRC - P30 AG062428 (James Leverenz, MD); Cleveland Clinic, Las Vegas - P20AG068053; Columbia - P50 AG008702 (PI Scott Small MD); Duke/UNC ADRC - P30 AG072958; Emory University - P30AG066511 (PI Levey Allan, MD, PhD); Indiana University - R01 AG19771 (PI Andrew Saykin, PsyD); P30 AG10133 (PI Andrew Saykin, PsyD); P30 AG072976 (PI Andrew Saykin, PsyD); R01 AG061788 (PI Shannon Risacher, PhD); R01 AG053993 (PI Yu-Chien Wu, MD, PhD); U01 AG057195 (PI Liana Apostolova, MD); U19 AG063911 (PI Bradley Boeve, MD); and the Indiana University Department of Radiology and Imaging Sciences; Johns Hopkins - P30 AG066507 (PI Marilyn Albert, Phd.); Mayo Clinic - P50 AG016574 (PI Ronald Petersen MD PhD); Mount Sinai - P30 AG066514 (PI Mary Sano, PhD); R01 AG054110 (PI Trey Hedden, PhD); R01 AG053509 (PI Trey Hedden, PhD); New York University - P30AG066512-01S2 (PI Thomas Wisniewski, MD); R01AG056031 (PI Ricardo Osorio, MD); R01AG056531 (PIs Ricardo Osorio, MD; Girardin Jean-Louis, PhD); North-western University - P30 AG013854 (PI Robert Vassar PhD); R01 AG045571 (PI Emily Rogalski, PhD); R56 AG045571, (PI Emily Rogalski, PhD); R01 AG067781, (PI Emily Rogalski, PhD); U19 AG073153, (PI Emily Rogalski, PhD); R01 DC008552, (M.-Marsel Mesulam, MD); R01 AG077444, (PIs M.-Marsel Mesulam, MD, Emily Rogalski, PhD); R01 NS075075 (PI Emily Rogalski, PhD); R01 AG056258 (PI Emily Rogalski, PhD); Oregon Health and Science University - P30 AG008017 (PI Jeffrey Kaye MD); R56 AG074321 (PI Jeffrey Kaye, MD); Rush University - P30 AG010161 (PI David Bennett MD); Stanford - P30AG066515; P50 AG047366 (PI Victor Henderson MD MS); University of Alabama, Birmingham - P20; University of California, Davis - P30 AG10129 (PI Charles DeCarli, MD); P30 AG072972 (PI Charles DeCarli, MD); University of California, Irvine - P50 AG016573 (PI Frank LaFerla PhD); University of California, San Diego - P30AG062429 (PI James Brewer, MD, PhD); University of California, San Francisco - P30 AG062422 (Rabinovici, Gil D., MD); University of Kansas - P30 AG035982 (Russell Swerdlow, MD); University of Kentucky - P30 AG028283-15S1 (PIs Linda Van Eldik, PhD and Brian Gold, PhD); University of Michigan ADRC - P30AG053760 (PI Henry Paulson, MD, PhD) P30AG072931 (PI Henry Paulson, MD, PhD) Cure Alzheimer’s Fund 200775 - (PI Henry Paulson, MD, PhD) U19 NS120384 (PI Charles DeCarli, MD, University of Michigan Site PI Henry Paulson, MD, PhD) R01 AG068338 (MPI Bruno Giordani, PhD, Carol Persad, PhD, Yi Murphey, PhD) S10OD026738-01 (PI Douglas Noll, PhD) R01 AG058724 (PI Benjamin Hampstead, PhD) R35 AG072262 (PI Benjamin Hampstead, PhD) W81XWH2110743 (PI Benjamin Hampstead, PhD) R01 AG073235 (PI Nancy Chiaravalloti, University of Michigan Site PI Benjamin Hampstead, PhD) 1I01RX001534 (PI Benjamin Hampstead, PhD) IRX001381 (PI Benjamin Hampstead, PhD); University of New Mexico - P20 AG068077 (Gary Rosenberg, MD); University of Pennsylvania - State of PA project 2019NF4100087335 (PI David Wolk, MD); Rooney Family Research Fund (PI David Wolk, MD); R01 AG055005 (PI David Wolk, MD); University of Pittsburgh - P50 AG005133 (PI Oscar Lopez MD); University of Southern California - P50 AG005142 (PI Helena Chui MD); University of Washington - P50 AG005136 (PI Thomas Grabowski MD); University of Wisconsin - P50 AG033514 (PI Sanjay Asthana MD FRCP); Vanderbilt University - P20 AG068082; Wake For-est - P30AG072947 (PI Suzanne Craft, PhD); Washington University, St. Louis - P01 AG03991 (PI John Morris MD); P01 AG026276 (PI John Morris MD); P20 MH071616 (PI Dan Marcus); P30 AG066444 (PI John Morris MD); P30 NS098577 (PI Dan Marcus); R01 AG021910 (PI Randy Buckner); R01 AG043434 (PI Catherine Roe); R01 EB009352 (PI Dan Marcus); UL1 TR000448 (PI Brad Evanoff); U24 RR021382 (PI Bruce Rosen); Avid Radiopharmaceuticals / Eli Lilly; Yale - P50 AG047270 (PI Stephen Strittmatter MD PhD); R01AG052560 (MPI: Christopher van Dyck, MD; Richard Carson, PhD); R01AG062276 (PI: Christopher van Dyck, MD); 1Florida - P30AG066506-03 (PI Glenn Smith, PhD); P50 AG047266 (PI Todd Golde MD PhD) Data were provided by OASIS-3: Longitudinal Multimodal Neuroimaging: Principal Investigators: T. Benzinger, D. Marcus, J. Morris; NIH P30 AG066444, P50 AG00561, P30 NS09857781, P01 AG026276, P01 AG003991, R01 AG043434, UL1 TR000448, R01 EB009352. AV-45 doses were provided by Avid Radiopharmaceuticals, a wholly owned subsidiary of Eli Lilly.

This research has been conducted using the UK Biobank Resource under Project ID 52887.

## Notes

### Competing Interest Statement

The authors have declared no competing interest.

### Author Declarations

Source data were openly available before the initiation of this study from the following sources: A4/LEARN - https://www.a4studydata.org/ ADNI - https://adni.loni.usc.edu/ AIBL - https://aibl.csiro.au/adni/index.html HABS - https://www.synapse.org/Synapse:syn53910452/wiki/631007 NACC - https://naccdata.org/requesting-data/data-request-process OASIS - https://sites.wustl.edu/oasisbrains/home/access/ UK Biobank - https://www.ukbiobank.ac.uk/use-our-data/apply-for-access/

## References

[1] Livingston G, Huntley J, Liu KY, Costafreda SG, Selbæk G, Alladi S, et al. Dementia prevention, intervention, and care: 2024 report of the Lancet standing Commission. The Lancet. 2024;404(10452):572–628.

2. Organization WH.: World Health Organization. Available from: https://www.who.int/news-room/fact-sheets/detail/dementia.

[3] Jack Jr CR, Andrews JS, Beach TG, Buracchio T, Dunn B, Graf A, et al. Revised criteria for diagnosis and staging of Alzheimer’s disease: Alzheimer’s Association Workgroup. Alzheimer’s & Dementia. 2024;20(8):5143–5169.

[4] Ashton NJ, Brum WS, Di Molfetta G, Benedet AL, Arslan B, Jonaitis E, et al. Diagnostic accuracy of a plasma phosphorylated tau 217 immunoassay for Alzheimer disease pathology. JAMA neurology. 2024;81(3):255–263.

[5] Brum WS, Cullen NC, Janelidze S, Ashton NJ, Zimmer ER, Therriault J, et al. A two-step workflow based on plasma p-tau217 to screen for amyloid *ε* posi-tivity with further confirmatory testing only in uncertain cases. Nature Aging. 2023;3(9):1079–1090.

[6] Warmenhoven N, Salvadó G, Janelidze S, Mattsson-Carlgren N, Bali D, Orduña Dolado A, et al. A comprehensive head-to-head comparison of key plasma phosphorylated tau 217 biomarker tests. Brain. 2025;148(2):416–431.

[7] Palmqvist S, Tideman P, Mattsson-Carlgren N, Schindler SE, Smith R, Ossenkoppele R, et al. Blood biomarkers to detect Alzheimer disease in primary care and secondary care. JAMA. 2024;332(15):1245–1257.

[8] Shaw LM, Arias J, Blennow K, Galasko D, Molinuevo JL, Salloway S, et al. Appropriate use criteria for lumbar puncture and cerebrospinal fluid testing in the diagnosis of Alzheimer’s disease. Alzheimer’s & Dementia. 2018;14(11):1505–1521.

[9] Therriault J, Brum WS, Trudel L, Macedo AC, Bitencourt FV, Martins-Pfeifer CC, et al. Blood phosphorylated tau for the diagnosis of Alzheimer’s disease: a systematic review and meta-analysis. The Lancet Neurology. 2025;24(9):740–752.

[10] Burke BT, Latimer C, Keene CD, Sonnen JA, McCormick W, Bowen JD, et al. Theoretical impact of the AT (N) framework on dementia using a community autopsy sample. Alzheimer’s & Dementia. 2021;17(12):1879–1891.

[11] Jansen WJ, Janssen O, Tijms BM, Vos SJ, Ossenkoppele R, Visser PJ, et al. Prevalence estimates of amyloid abnormality across the Alzheimer disease clinical spectrum. JAMA neurology. 2022;79(3):228–243.

[12] Hansson O, Edelmayer RM, Boxer AL, Carrillo MC, Mielke MM, Rabinovici GD, et al. The Alzheimer’s Association appropriate use recommendations for blood biomarkers in Alzheimer’s disease. Alzheimer’s & Dementia. 2022;18(12):2669–2686.

[13] Hansson O, Blennow K, Zetterberg H, Dage J. Blood biomarkers for Alzheimer’s disease in clinical practice and trials. Nature aging. 2023;3(5):506–519.

[14] Insel PS, Mattsson-Carlgren N, Langford O, Caruso VM, Leuzy A, Young CB, et al. Concurrent changes in plasma phosphorylated tau 217, tau PET, and cognition in preclinical Alzheimer disease. JAMA neurology. 2025;.

[15] Martino-Adami PV, Chatterjee M, Kleineidam L, Weyerer S, Bickel H, Wiese B, et al. Prognostic value of Alzheimer’s disease plasma biomarkers in the oldest-old: a prospective primary care-based study. The Lancet Regional Health–Europe. 2024;45.

[16] Cullen NC, Leuzy A, Janelidze S, Palmqvist S, Svenningsson AL, Stomrud E, et al. Plasma biomarkers of Alzheimer’s disease improve prediction of cognitive decline in cognitively unimpaired elderly populations. Nature communications. 2021;12(1):3555.

[17] McKenna MR, Gbadeyan O, Andridge R, Schroeder MW, Pugh EA, Scharre DW, et al. p-Tau/A*ε*42 ratio associates with cognitive decline in Alzheimer’s disease, mild cognitive impairment, and cognitively unimpaired older adults. Neuropsychology. 2025;39(2):137.

[18] Mattsson-Carlgren N, Salvadó G, Ashton NJ, Tideman P, Stomrud E, Zetterberg H, et al. Prediction of longitudinal cognitive decline in preclinical Alzheimer disease using plasma biomarkers. JAMA neurology. 2023;80(4):360–369.

[19] Ossenkoppele R, Salvadó G, Janelidze S, Pichet Binette A, Bali D, Karlsson L, et al. Plasma p-tau217 and tau-PET predict future cognitive decline among cognitively unimpaired individuals: implications for clinical trials. Nature Aging. 2025;p. 1–14.

[20] Stocker H, Beyer L, Perna L, Rujescu D, Holleczek B, Beyreuther K, et al. Association of plasma biomarkers, p-tau181, glial fibrillary acidic protein, and neurofilament light, with intermediate and long-term clinical Alzheimer’s disease risk: results from a prospective cohort followed over 17 years. Alzheimer’s & Dementia. 2023;19(1):25–35.

[21] Van Gool WA, Siebrand JF, Brayne C, Larson EB, Richard E. Evidence gap in blood biomarkers for Alzheimer’s disease. bmj. 2025;390.

[22] Hampel H, Hu Y, Cummings J, Mattke S, Iwatsubo T, Nakamura A, et al. Blood-based biomarkers for Alzheimer’s disease: Current state and future use in a transformed global healthcare landscape. Neuron. 2023;111(18):2781–2799.

[23] Assfaw AD, Schindler SE, Morris JC. Advances in blood biomarkers for Alzheimer disease (AD): A review. The Kaohsiung Journal of Medical Sciences. 2024;40(8):692–698.

[24] Zheng H, Wu Z, Mielke M, Murray A, Ryan J. Plasma biomarkers of Alzheimer’s disease and neurodegeneration according to sociodemographic characteristics and chronic health conditions. The Journal of Prevention of Alzheimer’s Disease. 2024;11(5):1189–1197.

[25] Schindler SE, Galasko D, Pereira AC, Rabinovici GD, Salloway S, Súarez-Calvet M, et al. Acceptable performance of blood biomarker tests of amyloid pathol-ogy—recommendations from the Global CEO Initiative on Alzheimer’s Disease. Nature Reviews Neurology. 2024;p. 1–14.

[26] Schöll M, Verberk IM, Del Campo M, Delaby C, Therriault J, Chong JR, et al. Challenges in the practical implementation of blood biomarkers for Alzheimer’s disease. The Lancet Healthy Longevity. 2024;5(10).

27. Administration FD.: FDA clears first blood test used in diagnosing alzheimer’s disease. FDA. Available from: https://www.fda.gov/news-events/press-announcements/fda-clears-first-blood-test-used-diagnosing-alzheimers-disease.

[28] Sperling RA, Donohue M, Rissman R, Johnson K, Rentz D, Grill J, et al. Amyloid and tau prediction of cognitive and functional decline in unimpaired older individuals: longitudinal data from the A4 and LEARN studies. The Journal of Prevention of Alzheimer’s Disease. 2024;11(4):802–813.

[29] Planche V, Schindler S, Knopman DS, Frisoni G, Galasko D, Grill JD, et al. The science does not yet support regulatory approval of amyloid-targeting therapies for Alzheimer’s disease based solely on biomarker evidence. Alzheimer’s & Dementia. 2025;21(4):e70068.

[30] Bouteloup V, Villain N, Vidal JS, Gonzalez-Ortiz F, Yuksekel I, Santos C, et al. Cognitive Phenotyping and Interpretation of Alzheimer Blood Biomarkers. JAMA neurology. 2025;.

[31] Blanche P, Dartigues JF, Jacqmin-Gadda H. Estimating and comparing time-dependent areas under receiver operating characteristic curves for censored event times with competing risks. Statistics in medicine. 2013;32(30):5381–5397.

[32] Wilkinson T, Schnier C, Bush K, Rannikmäe K, Henshall DE, Lerpiniere C, et al. Identifying dementia outcomes in UK Biobank: a validation study of primary care, hospital admissions and mortality data. European journal of epidemiology. 2019;34:557–565.

[33] Miller KL, Alfaro-Almagro F, Bangerter NK, Thomas DL, Yacoub E, Xu J, et al. Multimodal population brain imaging in the UK Biobank prospective epidemiological study. Nature neuroscience. 2016;19(11):1523–1536.

[34] Kapoor S, Narayanan A. Leakage and the reproducibility crisis in machine-learning-based science. Patterns. 2023;4(9).

[35] Stallard PE, Ukraintseva SV, Doraiswamy PM. Changing Story of the Dementia Epidemic. JAMA. 2025;333(18):1579–1580.

[36] Baker LD, Espeland MA, Whitmer RA, Snyder HM, Leng X, Lovato L, et al. Structured vs self-guided multidomain lifestyle interventions for global cognitive function: the US POINTER randomized clinical trial. JAMA. 2025;334(8):681–691.

[37] Beach TG, Monsell SE, Phillips LE, Kukull W. Accuracy of the clinical diagnosis of Alzheimer disease at National Institute on Aging Alzheimer Disease Centers, 2005–2010. Journal of neuropathology and experimental neurology. 2012;71(4):266–273.

[38] Sperling RA, Donohue MC, Raman R, Rafii MS, Johnson K, Masters CL, et al. Trial of solanezumab in preclinical Alzheimer’s disease. New England Journal of Medicine. 2023;389(12):1096–1107.

[39] Ellis KA, Bush AI, Darby D, De Fazio D, Foster J, Hudson P, et al. The Australian Imaging, Biomarkers and Lifestyle (AIBL) study of aging: methodology and baseline characteristics of 1112 individuals recruited for a longitudinal study of Alzheimer’s disease. International Psychogeriatrics. 2009;21(4):672–687. 10.1017/S1041610209009405.

[40] Jack CR, Bernstein MA, Fox NC, Thompson P, Alexander G, Harvey D, et al. The Alzheimer’s Disease Neuroimaging Initiative (ADNI): MRI methods. Journal of Magnetic Resonance Imaging. 2008;27(4):685–691. 10.1002/jmri.21049.

[41] Petersen RC, Aisen PS, Beckett LA, Donohue MC, Gamst AC, Harvey DJ, et al. Alzheimer’s disease Neuroimaging Initiative (ADNI) clinical characterization. Neurology. 2010;74(3):201–209.

[42] Dagley AS, LaPoint M, Huijbers W, Hedden T, McLaren DG, Papp KV, et al. Harvard Aging Brain Study: dataset and accessibility. NeuroImage. 2017;144:255–258. 10.1016/j.neuroimage.2015.03.049.

[43] Besser L, Kukull W, Knopman DS, Chui H, Galasko D, Weintraub S, et al. Version 3 of the national Alzheimer’s coordinating center’s uniform data set. Alzheimer Disease & Associated Disorders. 2018;32(4):351–358.

44. National Alzheimer’s Coordinating Center.: Uniform Data Set (UDS) Version 4: Coding Guidebook. Accessed: 2025-10-01. Available from: https://files.alz.washington.edu/documentation/UDSv4CodingGuidebook.pdf.

[45] LaMontagne PJ, Benzinger TL, Morris JC, Keefe S, Hornbeck R, Xiong C, et al. OASIS-3: longitudinal neuroimaging, clinical, and cognitive dataset for normal aging and Alzheimer disease. medrxiv. 2019; p. 2019–12.

46. Therneau TM.: A Package for Survival Analysis in R. R package version 3.8-3. Available from: https://CRAN.R-project.org/package=survival.

[47] Youden WJ. Index for rating diagnostic tests. Cancer. 1950;3(1):32–35.

[48] Wang C, Wu Q, Weimer M, Zhu E. Flaml: A fast and lightweight automl library. Proceedings of Machine Learning and Systems. 2021;3:434–447.

[49] Ke G, Meng Q, Finley T, Wang T, Chen W, Ma W, et al. Lightgbm: A highly efficient gradient boosting decision tree. Advances in neural information processing systems. 2017;30.

[50] Knuppel A, Papier K, Fensom GK, Appleby PN, Schmidt JA, Tong TY, et al. Meat intake and cancer risk: prospective analyses in UK Biobank. International Journal of Epidemiology. 2020;49(5):1540–1552.

