## Supplemental Methods and Figures for "Prognostic performance across Alzheimer’s biomarkers, multi-modal physiological measures, and clinical history in asymptomatic individuals"

### 6 Supplement

#### 6.1 Supplementary Methods

##### 6.1.1 Data Sharing Statement

All of the data used in this study can be accessed via requests to the respective curators of the datasets, including A4/LEARN, ADNI, AIBL, HABS, NACC, OASIS, and the UK Biobank. All datasets except for the UK Biobank do not require payment as part of the procedures for gaining access to the data.

#### 6.2 Supplementary Figures and Tables

##### 6.2.1 pTau-217 and amyloid-PET in A4/LEARN

| | Model | Coef | P-value |
|---|---|---|---|
| 1 | pTau217+PET+Demo+Lancet | 1.557 (0.15) | 5.84e-28 - 8.34e-09 |
| 2 | pTau217+PET+Demo+APOE | 1.56 (0.122) | 4.02e-27 - 4.17e-09 |
| 3 | pTau217+PET+Demo+APOE+Lancet | 1.577 (0.117) | 5.73e-29 - 4.24e-10 |
| 4 | pTau217+Demo+APOE | 1.603 (0.144) | 2.69e-31 - 5.92e-09 |
| 5 | pTau217+Demo+Lancet | 1.606 (0.144) | 4.71e-32 - 2.86e-09 |
| 6 | pTau217+Demo+APOE+Lancet | 1.608 (0.114) | 3.34e-32 - 2.63e-10 |
| 7 | pTau217+PET+Demo | 1.637 (0.127) | 2.01e-27 - 2.8e-09 |
| 8 | pTau217+PET | 1.656 (0.139) | 3.82e-29 - 1.04e-09 |
| 9 | pTau217+Demo+APOE (-APOE) | 1.683 (0.142) | 7.82e-33 - 6.76e-10 |
| 10 | pTau217 | 1.701 (0.136) | 3.39e-33 - 1.95e-10 |

**Table S1**: Coefficients and p-values for pTau-217 in Cox models including different sets of predictors in the A4/LEARN study predicting clinical impairment (Clinical Dementia Rating score of 0.5 or greater on two consecutive visits). Coefficients are shown as mean (SD) across cross-validation folds, and p-values are shown as ranges across folds for the association of pTau-217 with clinical impairment. Demo = "Demographics" (age, sex, education); Lancet = Lancet modifiable risk factors (smoking, alcohol, substance use, aerobic exercise, walking, Geriatric Depression score, State-Trait Anxiety Inventory score, systolic blood pressure, diastolic blood pressure, BMI); PET = Amyloid positron emission tomography performed at baseline; APOE = Apolipoprotein E genotype.

| | Term | P-value | Coef | exp(Coef) | se(Coef) | z |
|---|---|---|---|---|---|---|
| 1 | pTau-217 | 4.55e-24–1.16e-21 | 0.51–0.562 | 1.67–1.75 | 0.0515–0.0573 | 9.56–10.1 |
| 2 | GDScore | 3.43e-08–3.04e-05 | 0.2–0.25 | 1.22–1.28 | 0.0452–0.0492 | 4.17–5.52 |
| 3 | Sex-Female | 7.13e-06–2e-05 | 0.524–0.548 | 1.69–1.73 | 0.121–0.123 | 4.27–4.49 |
| 4 | APOEe2 carrier | 3.27e-05–0.0163 | 0.767–1.26 | 2.15–3.54 | 0.297–0.337 | 2.4–4.15 |
| 5 | APOEe3/e4 | 0.00151–0.0419 | 0.401–0.654 | 1.49–1.92 | 0.197–0.206 | 2.03–3.17 |
| 6 | Walking | 0.0595–0.173 | -0.136—0.0923 | 0.872–0.912 | 0.0678–0.0731 | -1.88—1.36 |
| 7 | Smoking | 0.0311–0.204 | -0.464–0.0408 | 0.629–1.04 | 0.0189–0.366 | -1.27–2.16 |
| 8 | APOEe2/e4 | 0.0272–0.305 | 0.442–0.869 | 1.56–2.38 | 0.393–0.431 | 1.03–2.21 |
| 9 | Age:APOEe2 carrier | 0.0655–0.376 | -0.186—0.11 | 0.83–0.896 | 0.0982–0.124 | -1.84—0.885 |
| 10 | Systolic BP | 0.0524–0.365 | 0.0588–0.125 | 1.06–1.13 | 0.0646–0.0652 | 0.905–1.94 |
| 11 | Aerobic exercise | 0.0326–0.425 | 0.0475–0.116 | 1.05–1.12 | 0.0545–0.0596 | 0.798–2.14 |
| 12 | $Age^2$ | 0.0688–0.436 | 0.00258–0.00575 | 1–1.01 | 0.00316–0.0035 | 0.779–1.82 |
| 13 | Education | 0.0311–0.715 | -0.13–0.0223 | 0.878–1.02 | 0.06–0.0621 | -2.16–0.365 |
| 14 | Substance use | 0.12–0.812 | -0.705—0.0863 | 0.494–0.917 | 0.362–0.453 | -1.55—0.238 |
| 15 | Age:APOEe4/e4 | 0.122–0.824 | -0.14–0.026 | 0.869–1.03 | 0.089–0.117 | -1.55–0.223 |
| 16 | $Age^2$:APOEe2/e4 | 0.142–0.755 | -0.0108–0.0139 | 0.989–1.01 | 0.00875–0.0196 | -0.551–1.47 |
| 17 | $Age^2$:APOEe4/e4 | 0.0815–0.991 | -0.000132–0.0158 | 1–1.02 | 0.00907–0.0111 | -0.0119–1.74 |
| 18 | $Age^2$:APOEe3/e4 | 0.226–0.924 | -0.00523—0.000421 | 0.995–1 | 0.00432–0.00469 | -1.21—0.0953 |
| 19 | Diastolic BP | 0.169–0.826 | -0.0928–0.022 | 0.911–1.02 | 0.0666–0.0674 | -1.38–0.327 |
| 20 | BMI | 0.153–0.89 | -0.0497–0.0875 | 0.952–1.09 | 0.0612–0.0646 | -0.77–1.43 |
| 21 | Age:APOEe2/e4 | 0.216–0.913 | -0.164–0.0941 | 0.849–1.1 | 0.12–0.167 | -1.24–0.564 |
| 22 | APOEe4/e4 | 0.226–0.931 | -0.13–0.372 | 0.878–1.45 | 0.307–0.342 | -0.381–1.21 |
| 23 | Age | 0.511–0.928 | 0.0043–0.0313 | 1–1.03 | 0.0471–0.0508 | 0.0903–0.657 |
| 24 | Age2:APOEe2 carrier | 0.43–0.974 | -0.0119–0.00277 | 0.988–1 | 0.00906–0.0151 | -0.789–0.291 |
| 25 | STAI | 0.296–0.982 | -0.00509–0.0647 | 0.995–1.07 | 0.0619–0.0626 | -0.0813–1.04 |
| 26 | Age:APOEe3/e4 | 0.619–0.987 | -0.0295–0.0248 | 0.971–1.03 | 0.0592–0.0622 | -0.498–0.418 |
| 27 | Alcohol | 0.535–0.969 | -0.0345–0.00522 | 0.966–1.01 | 0.0543–0.0578 | -0.62–0.0928 |

**Table S2**: Statistics for the pTau217+Demographics+Lancet Cox model in the A4/LEARN study predicting clinical impairment (Clinical Dementia Rating score of 0.5 or greater on two consecutive visits). Statistics are shown as ranges across cross-validation folds. Demographics = age, sex, education; Lancet = Lancet modifiable risk factors (smoking, alcohol, substance use, aerobic exercise, walking, Geriatric Depression score, State-Trait Anxiety Inventory score, systolic blood pressure, diastolic blood pressure, BMI); APOE = Apolipoprotein E genotype.

| Fold | 3y | 4y | 5y | 6y | 7y |
|---|---|---|---|---|---|
| 1 | 0.1993 | 0.8776 | 0.3044 | 0.9497 | 0.4036 |
| 2 | 0.1118 | **0.0097** | 0.8407 | 0.7316 | **0.0278** |
| 3 | 0.3977 | **0.0246** | 0.1532 | 0.199 | **0.0357** |
| 4 | 1 | 0.1789 | **8e-04** | 0.4161 | **0.0122** |
| 5 | 0.2984 | 0.5846 | 0.1305 | 0.5116 | 0.3089 |

**Table S3**: P-values comparing the AUROC of Demographics+APOE+Lancet vs. pTau-217+Demographics+APOE+Lancet models across five cross-validation folds and five time points (Years 3-7) in A4/LEARN predicting clinical impairment (Clinical Dementia Rating score of 0.5 or greater on two consecutive visits). Six out of 25 p-values were statistically significant after correcting for multiple testing.

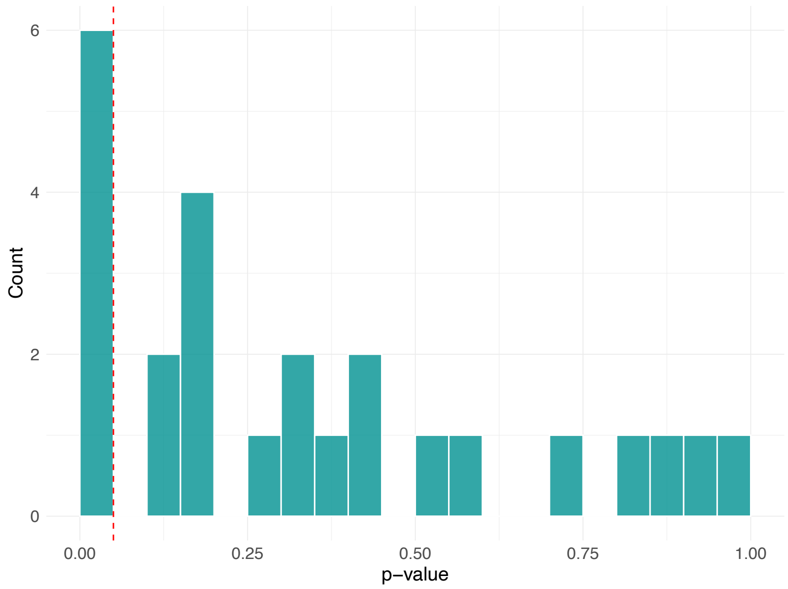


**Fig. S1**: Histogram of P-values comparing the AUROC of Demographics+APOE+Lancet vs. pTau-217+Demographics+APOE+Lancet models across five cross-validation folds and five time points (Years 3-7) in A4/LEARN predicting clinical impairment (Clinical Dementia Rating score of 0.5 or greater on two consecutive visits). Six out of 25 p-values were statistically significant after correcting for multiple testing.

| Fold | 3y | 4y | 5y | 6y | 7y |
|---|---|---|---|---|---|
| 1 | 0.4167 | 0.3584 | **0.0023** | 0.2207 | **5e-04** |
| 2 | **3e-04** | **4e-04** | **0.0038** | **0.0098** | **0.007** |
| 3 | 0.1199 | **0.0424** | **0.0133** | **4e-04** | **0.0151** |
| 4 | 1 | 0.3182 | **0** | **0.0167** | **0.004** |
| 5 | 0.3218 | 0.1046 | **4e-04** | 0.1532 | 0.0812 |

**Table S4**: P-values for time-varying AUROC with 95% confidence intervals for all folds and time points comparing PET+Demographics+APOE+Lancet vs. Demographics+APOE+Lancet in A4/LEARN predicting clinical impairment (Clinical Dementia Rating score of 0.5 or greater on two consecutive visits). 15 out of 25 p-values were statistically significant after correcting for multiple testing.

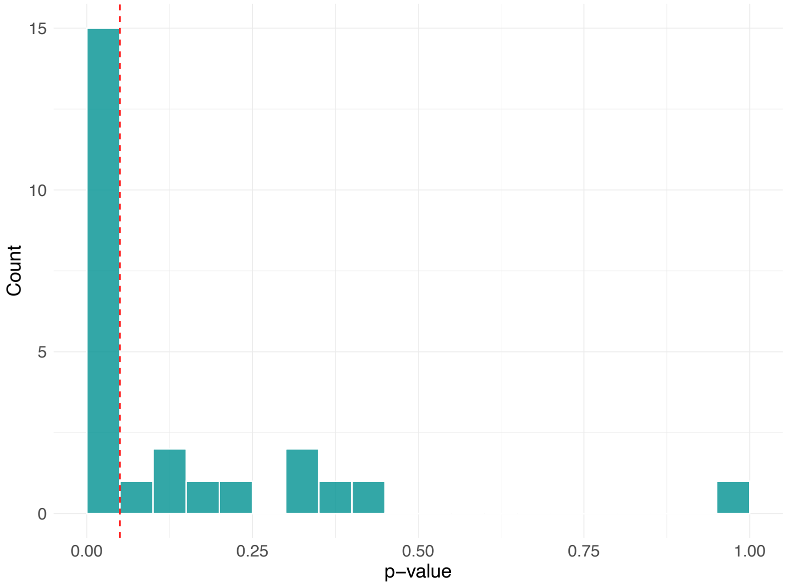


**Fig. S2**: Histogram of P-values comparing Demographics+APOE+Lancet vs. PET+Demographics+APOE+Lancet models across five cross-validation folds and five time points (Years 3-7) in A4/LEARN predicting clinical impairment (Clinical Dementia Rating score of 0.5 or greater on two consecutive visits). 15 out of 25 p-values were statistically significant after correcting for multiple testing.

| Model | 3y | 4y | 5y | 6y | 7y |
|---|---|---|---|---|---|
| Demo | 0.55 (0.43-0.68) | 0.55 (0.46-0.65) | 0.54 (0.45-0.62) | 0.56 (0.46-0.65) | 0.55 (0.45-0.65) |
| Demo+APOE+Lancet (-APOE) | 0.59 (0.47-0.71) | 0.60 (0.51-0.69) | 0.59 (0.50-0.67) | 0.62 (0.53-0.70) | 0.61 (0.51-0.72) |
| Demo+APOE | 0.59 (0.47-0.72) | 0.60 (0.50-0.69) | 0.58 (0.50-0.67) | 0.60 (0.51-0.69) | 0.57 (0.47-0.68) |
| Lancet | 0.59 (0.48-0.70) | 0.59 (0.50-0.69) | 0.61 (0.52-0.69) | 0.63 (0.54-0.71) | 0.61 (0.51-0.71) |
| Demo+APOE+Lancet | 0.61 (0.49-0.73) | 0.62 (0.54-0.71) | 0.61 (0.53-0.70) | 0.64 (0.55-0.73) | 0.62 (0.51-0.72) |
| pTau217 | 0.66 (0.54-0.78) | 0.66 (0.57-0.75) | 0.67 (0.60-0.75) | 0.64 (0.56-0.72) | 0.67 (0.59-0.76) |
| PET+Demo | 0.67 (0.55-0.79) | 0.66 (0.57-0.76) | 0.70 (0.63-0.77) | 0.69 (0.61-0.77) | 0.70 (0.61-0.78) |
| PET | 0.67 (0.55-0.80) | 0.66 (0.56-0.76) | 0.71 (0.64-0.78) | 0.68 (0.61-0.76) | 0.69 (0.60-0.78) |
| pTau217+Demo | 0.68 (0.56-0.79) | 0.68 (0.59-0.76) | 0.68 (0.60-0.75) | 0.66 (0.58-0.73) | 0.69 (0.61-0.78) |
| pTau217+PET | 0.68 (0.56-0.80) | 0.67 (0.58-0.76) | 0.70 (0.63-0.78) | 0.67 (0.59-0.74) | 0.69 (0.61-0.78) |
| PET+Demo+APOE | 0.68 (0.56-0.80) | 0.68 (0.59-0.77) | 0.71 (0.64-0.78) | 0.70 (0.62-0.78) | 0.69 (0.60-0.78) |
| pTau217+Demo+APOE | 0.68 (0.57-0.79) | 0.69 (0.61-0.77) | 0.68 (0.61-0.76) | 0.67 (0.59-0.74) | 0.69 (0.60-0.78) |
| pTau217+Demo+Lancet | 0.69 (0.58-0.79) | 0.68 (0.60-0.77) | 0.68 (0.61-0.75) | 0.68 (0.60-0.75) | 0.71 (0.62-0.79) |
| PET+Demo+Lancet | 0.69 (0.58-0.80) | 0.69 (0.61-0.77) | 0.72 (0.66-0.79) | 0.72 (0.65-0.79) | 0.72 (0.64-0.81) |
| PET+Demo+APOE+Lancet | 0.69 (0.58-0.80) | 0.70 (0.61-0.78) | 0.72 (0.65-0.79) | 0.72 (0.65-0.80) | 0.72 (0.62-0.81) |
| pTau217+PET+Demo | 0.69 (0.58-0.81) | 0.69 (0.60-0.78) | 0.70 (0.63-0.77) | 0.68 (0.61-0.76) | 0.71 (0.63-0.79) |
| pTau217+Demo+APOE+Lancet | 0.69 (0.59-0.79) | 0.69 (0.61-0.78) | 0.69 (0.62-0.76) | 0.68 (0.61-0.76) | 0.70 (0.61-0.79) |
| pTau217+PET+Demo+APOE | 0.70 (0.59-0.81) | 0.70 (0.62-0.79) | 0.71 (0.64-0.78) | 0.70 (0.62-0.77) | 0.71 (0.62-0.80) |
| pTau217+PET+Demo+Lancet | 0.70 (0.60-0.81) | 0.70 (0.61-0.78) | 0.71 (0.64-0.78) | 0.71 (0.63-0.78) | 0.73 (0.64-0.81) |
| pTau217+PET+Demo+APOE+Lancet | 0.71 (0.60-0.81) | 0.71 (0.62-0.79) | 0.72 (0.65-0.79) | 0.71 (0.64-0.79) | 0.72 (0.63-0.81) |

**Table S5**: Time-varying AUROC with 95% confidence intervals for all models and time points in A4/LEARN predicting clinical impairment (Clinical Dementia Rating score of 0.5 or greater on two consecutive visits).

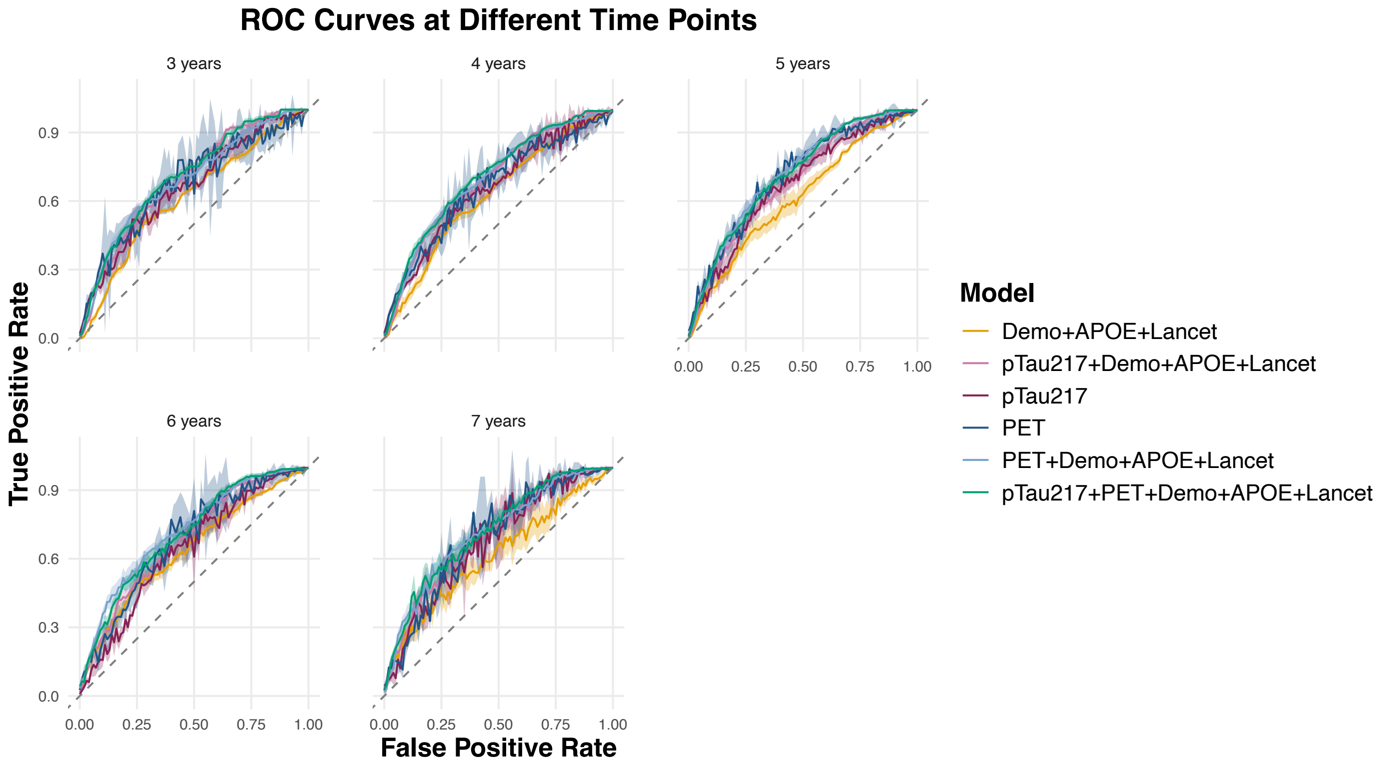


**Fig. S3**: ROC curves for Years 3-7 in the A4 study for the prediction of CDR-based clinical impairment in A4/LEARN. Demographics include age, sex, and years of education. Lancet factors include smoking, alcohol, substance use, aerobic exercise, walking, Geriatric Depression score, State-Trait Anxiety Inventory score, systolic blood pressure, diastolic blood pressure, BMI.

| Model | Mean pAUROC | SD |
|---|---|---|
| Demo+APOE+Lancet | 0.2409 | 0.0308 |
| pTau217 | 0.2626 | 0.0329 |
| PET | 0.2890 | 0.0265 |
| pTau217+Demo+APOE+Lancet | 0.3066 | 0.0163 |
| pTau217+PET+Demo+APOE+Lancet | 0.3348 | 0.0176 |
| PET+Demo+APOE+Lancet | 0.3356 | 0.0180 |

**Table S6**: Mean partial AUROC (FPR range: 0-25%), averaged across five cross-validation folds and five time points (Years 3-7) in A4/LEARN predicting clinical impairment (Clinical Dementia Rating score of 0.5 or greater on two consecutive visits).

| **N=1440** | **Cases (409)** | **Controls (1031)** |
|---|---|---|
| Age (Mean ± SD) | 72.88 ± 5.00 | 71.23 ± 4.58 |
| Sex | | |
| Female | 195 (47.68%) | 661 (64.11%) |
| Male | 214 (52.32%) | 370 (35.89%) |
| APOE Genotype | | |
| E3/E3 | 137 (33.5%) | 464 (45%) |
| E2/E4 | 15 (3.67%) | 26 (2.52%) |
| E4/E4 | 28 (6.85%) | 63 (6.11%) |
| E3/E4 | 203 (49.63%) | 407 (39.48%) |
| E2 carrier | 25 (6.11%) | 71 (6.89%) |

**Table S7**: A4/LEARN - Summary statistics for age, sex, and APOE genotype by group. "Cases" refers to patients who progress to the criterion of clinical impairment (Clinical Dementia Rating score of 0.5 or greater on two consecutive visits) and "Controls" refers to patients who do not.

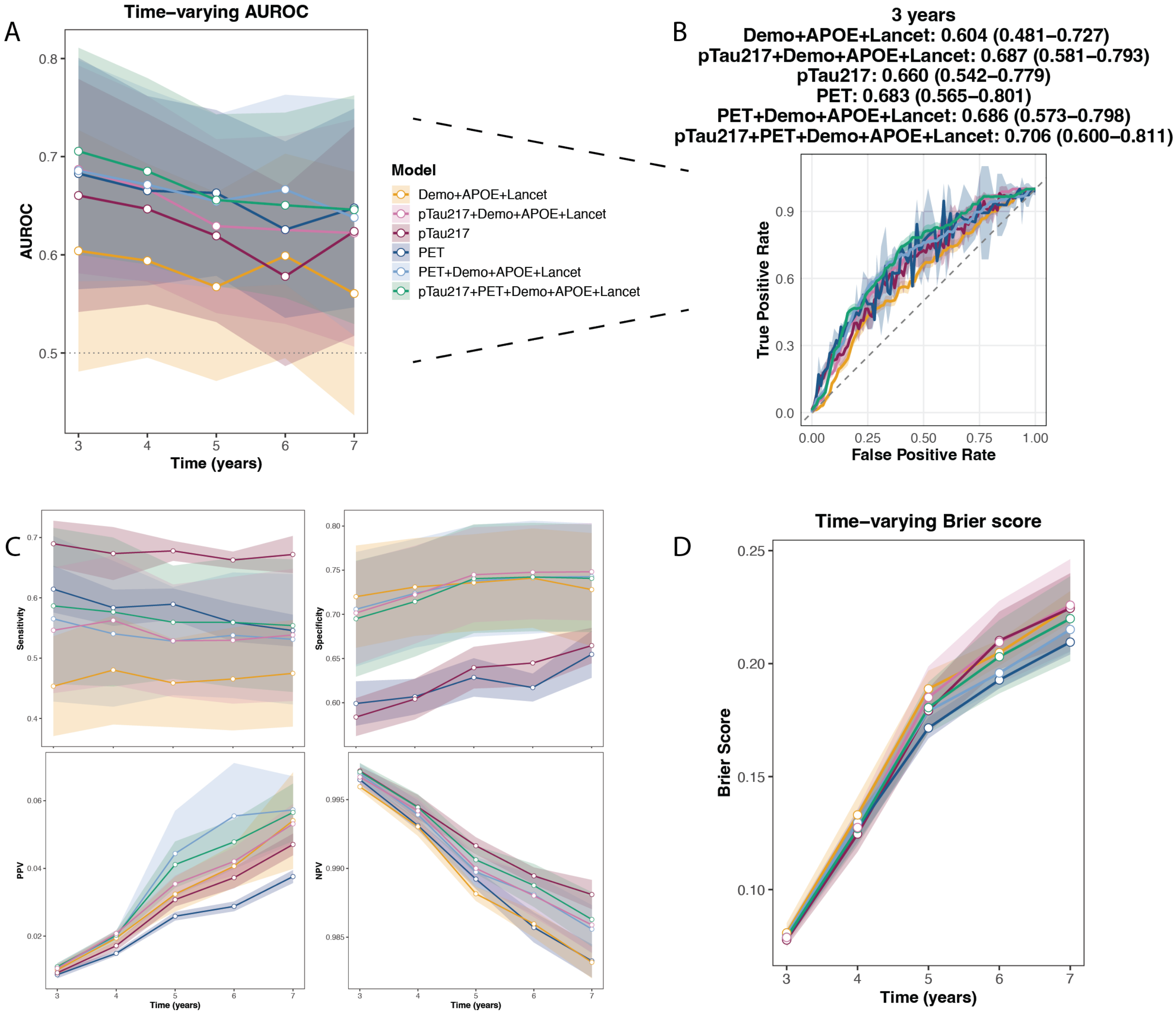


**Fig. S4**: Predicting CDR-based clinical impairment in only amyloid-positive participants using six models in A4/LEARN: 1) Demo + Lancet (demographics, APOE and Lancet modifiable risk factors), 2) pTau-217, 3) pTau217 + Demographics + Lancet, 4) Amyloid PET, 5) PET + Demographics + Lancet, 6) pTau217 + PET + Demographics + Lancet. A) Time-varying AUROC for each year, with 95% confidence intervals. B) ROC curve for Year 3, which showed the largest difference in mean AUC between Models (1) and (3). C) Sensitivity, specificity, positive predictive value, and negative predictive value for all six models using optimal cutpoints by Youden's J statistic. D) Time-varying Brier score for each year, with standard deviations.

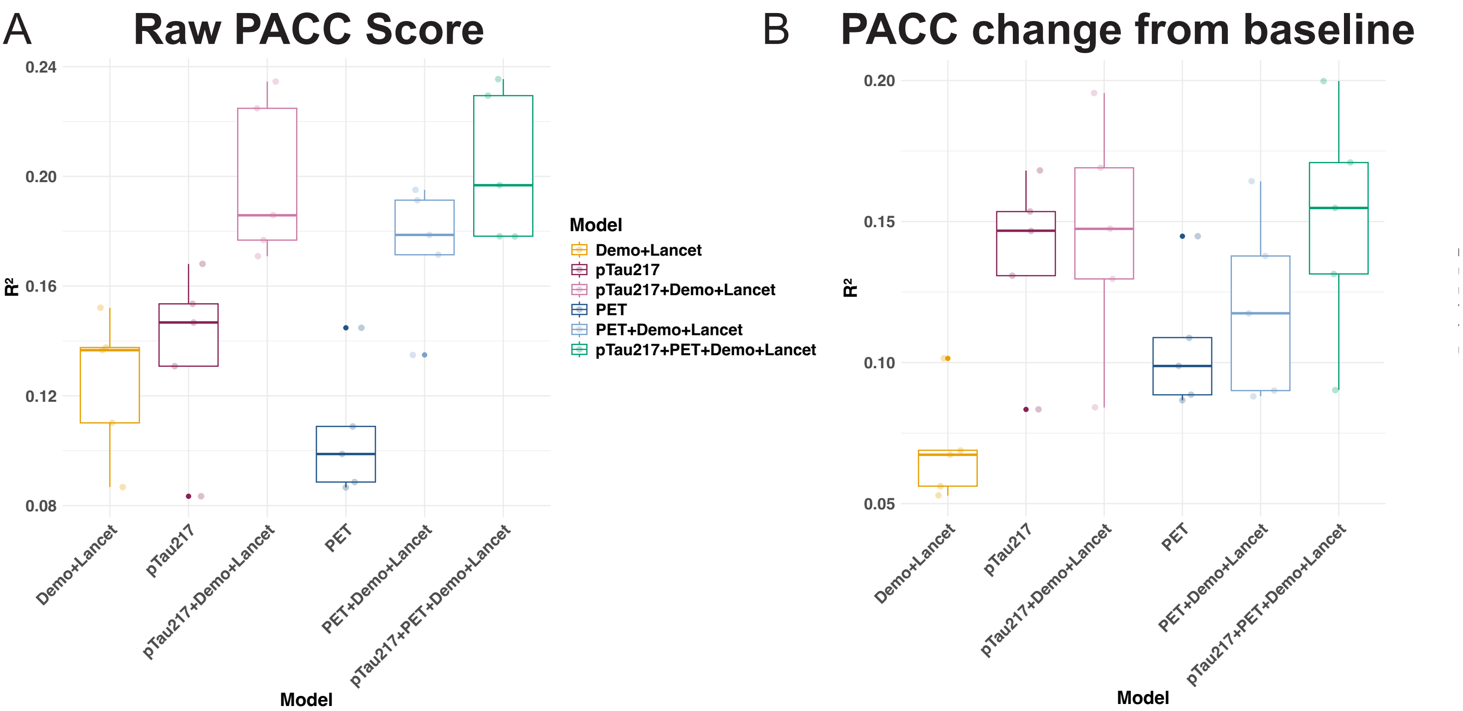


**Fig. S5**: Predicting PACC score using six models in A4/LEARN: 1) Demo + Lancet (demographics, APOE, and Lancet modifiable risk factors), 2) pTau-217, 3) pTau217 + Demographics + Lancet, 4) Amyloid PET, 5) PET + Demographics + Lancet, 6) pTau217 + PET + Demographics + Lancet. A) Boxplots of raw PACC score across five folds. B) Boxplots of PACC change from baseline across five folds.

##### 6.2.2 pTau-217 in ADNI

| | Model | Coef | P-value |
|---|---|---|---|
| 1 | pTau217+Demo+Lancet (-APOE) | 1.988 (0.223) | 9.17e-10 - 4.95e-04 |
| 2 | pTau217 | 2.169 (0.157) | 7.58e-10 - 1.13e-03 |
| 3 | pTau217+Demo | 2.204 (0.323) | 5.44e-10 - 7.38e-04 |
| 4 | pTau217+Demo+Lancet | 2.73 (0.362) | 2.00e-10 - 2.04e-04 |
| 5 | pTau217+Demo (-APOE) | 2.738 (0.169) | 4.23e-11 - 8.31e-05 |

**Table S8**: Coefficients and p-values for pTau-217 in Cox models in the ADNI cohort predicting clinical AD diagnosis. Coefficients are shown as mean (SD) across cross-validation folds, and p-values are shown as ranges across folds.

| | Fold | 2y | 3y | 4y | 5y | 6y | 7y |
|---|---|---|---|---|---|---|---|
| 1 | 1 | **0** | 0.0617 | **0.012** | 0.1557 | 0.197 | 0.2485 |
| 2 | 2 | 0.1993 | 0.4616 | 0.3617 | 0.4639 | 0.4632 | 1 |
| 3 | 3 | 0.0984 | 0.1835 | 0.9216 | 0.9034 | 1 | 1 |
| 4 | 4 | **0.0242** | **0.0176** | **0.004** | **0.0041** | **0.0091** | **0.0121** |
| 5 | 5 | **0.0084** | 0.4862 | 0.5406 | 0.3476 | 0.2436 | 0.5369 |

**Table S9**: P-values for time-varying AUROC with 95% confidence intervals for all folds and time points comparing pTau+Demographics+APOE+Lancet vs. Demographics+APOE+Lancet in ADNI predicting clinical AD diagnosis.

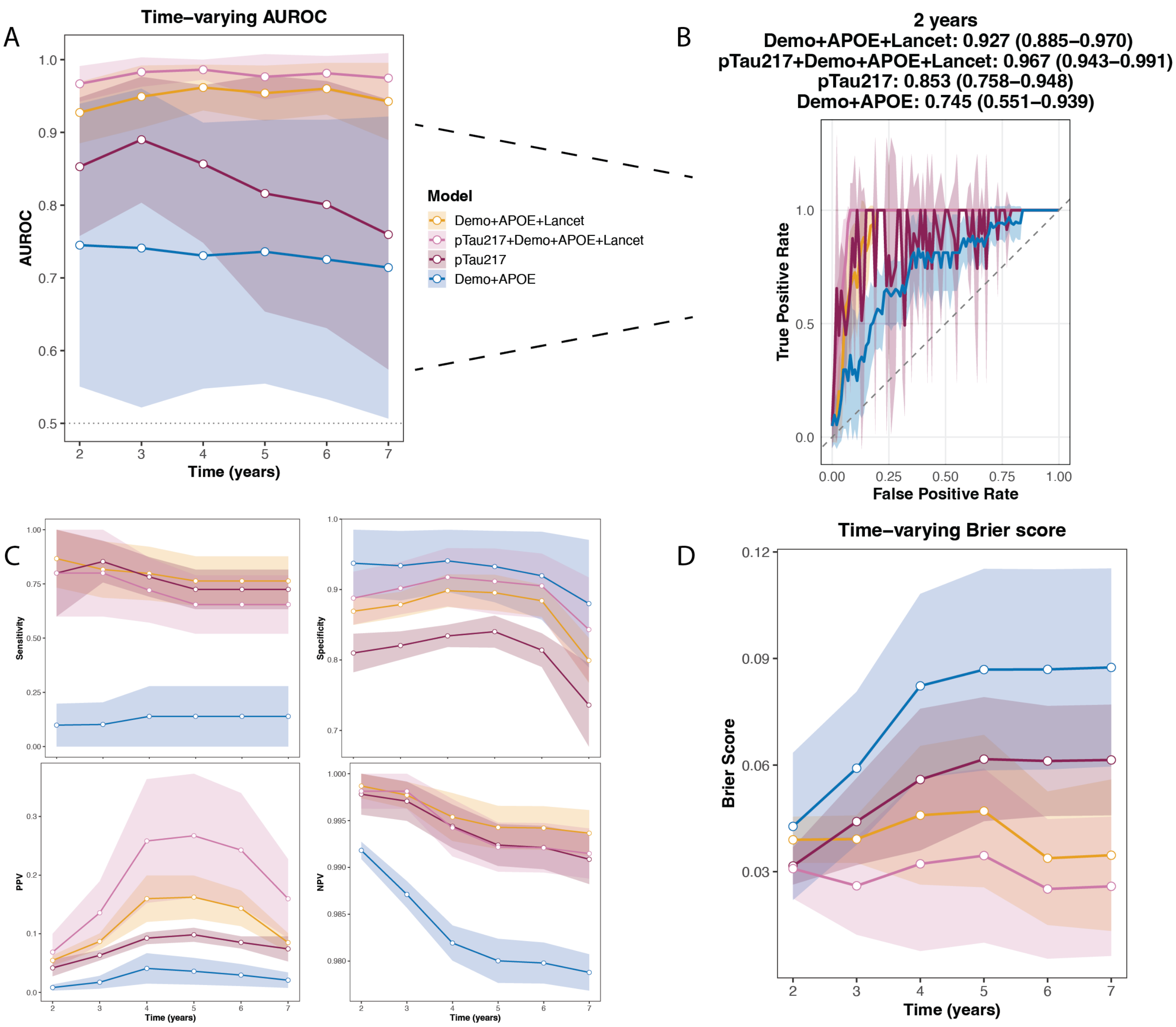


**Fig. S6**: Predicting clinical AD diagnosis in ADNI: 1) Demo + APOE + Lancet, 2) pTau-217, 3) pTau217 + Demographics + APOE + Lancet, 4) Demo + APOE. A) Time-varying AUROC for each year, with 95% confidence intervals. B) ROC curve for Year 2, which showed the largest difference in mean AUC between Models (1) and (3). C) Sensitivity, specificity, positive predictive value, and negative predictive value for all six models using optimal cutpoints by Youden's J statistic. D) Time-varying Brier score for each year, with standard deviations.

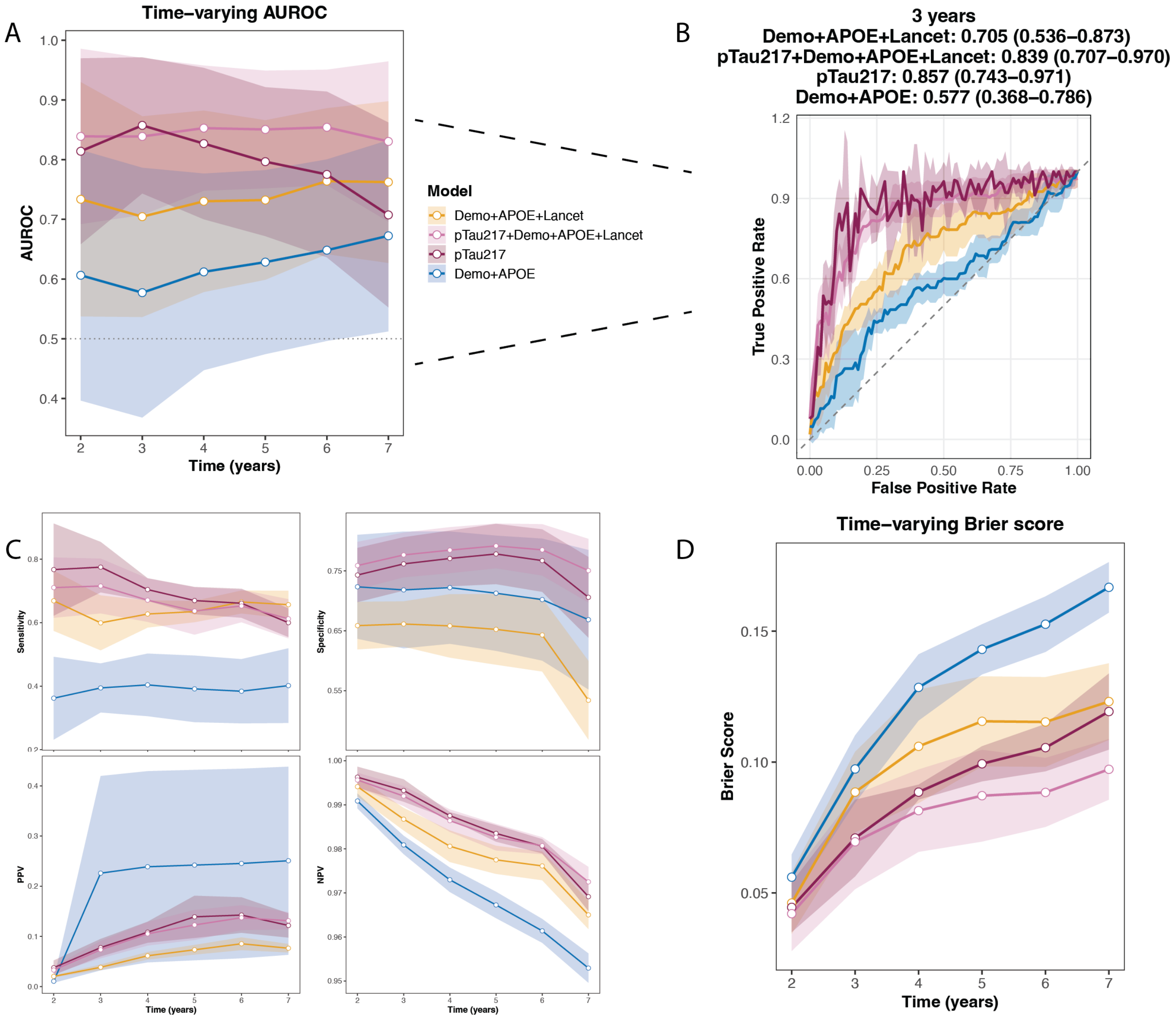


**Fig. S7**: Predicting all-cause dementia diagnosis in ADNI: 1) Demo + APOE + Lancet, 2) pTau-217, 3) pTau217 + Demographics + APOE + Lancet, 4) Demo + APOE. A) Time-varying AUROC for each year, with 95% confidence intervals. B) ROC curve for Year 2, which showed the largest difference in mean AUC between Models (1) and (3). C) Sensitivity, specificity, positive predictive value, and negative predictive value for all six models using optimal cutpoints by Youden's J statistic. D) Time-varying Brier score for each year, with standard deviations.

##### 6.2.3 Amyloid-PET in ADNI, AIBL, HABS, NACC, OASIS

| | Model | Coef | P-value |
|---|---|---|---|
| 1 | PET | 1.013 (0.001) | 7.96e-33 - 1.84e-16 |
| 2 | PET+Demo+APOE | 1.014 (0.001) | 1.05e-36 - 4.79e-14 |
| 3 | PET+Demo | 1.014 (0.002) | 1.19e-36 - 2.94e-14 |

**Table S10**: Coefficients and p-values for amyloid-PET in Cox models in five PET cohorts predicting clinical AD diagnosis. Coefficients are shown as mean (SD) across cross-validation folds, and p-values are shown as ranges across folds.

| **N=2354** | **Cases (244)** | **Controls (2110)** |
|---|---|---|
| Age (Mean ± SD) | 79.21 ± 6.59 | 75.16 ± 6.37 |
| Sex | | |
| Female | 114 | 1240 |
| Male | 130 | 870 |
| APOE Genotype | | |
| E3/E4 | 83 | 528 |
| E4/E4 | 14 | 69 |
| E2 carriers | 19 | 249 |
| E3/E3 | 117 | 1093 |
| E2/E4 | 3 | 57 |

**Table S11**: Five PET cohorts for predicting clinical AD diagnosis - Summary statistics for age, sex, and APOE genotype by group. "Cases" refers to patients who receive a diagnosis of Alzheimer's disease during follow-up and "Controls" refers to patients who do not.

| | Term | P-value | Coef | exp(Coef) | se(Coef) | z |
|---|---|---|---|---|---|---|
| 1 | PET | 1.42e-19 - 4.79e-14 | 0.011 - 0.013 | 1.011 - 1.013 | 0.001 - 0.002 | 7.538 - 9.051 |
| 2 | Age | 3.89e-05 - 0.000353 | 0.057 - 0.067 | 1.059 - 1.069 | 0.016 - 0.016 | 3.573 - 4.114 |
| 3 | Female | 9.67e-05 - 0.0121 | -0.554 - -0.355 | 0.574 - 0.701 | 0.141 - 0.142 | -3.899 - -2.508 |
| 4 | Education | 0.00346 - 0.104 | -0.234 - -0.126 | 0.791 - 0.882 | 0.076 - 0.080 | -2.923 - -1.624 |
| 5 | APOEe3/e4 | 0.0398 - 0.905 | 0.024 - 0.405 | 1.024 - 1.499 | 0.197 - 0.200 | 0.119 - 2.056 |
| 6 | APOEe4/e4 | 0.0894 - 0.426 | 0.265 - 0.564 | 1.303 - 1.758 | 0.326 - 0.337 | 0.795 - 1.698 |
| 7 | $Age^2$:APOEe4/e4 | 0.103 - 0.499 | 0.002 - 0.003 | 1.002 - 1.003 | 0.002 - 0.002 | 0.677 - 1.632 |
| 8 | APOEe2/e4 | 0.123 - 0.578 | -2.939 - -0.515 | 0.053 - 0.597 | 0.877 - 2.951 | -1.544 - -0.557 |
| 9 | Age:APOEe3/e4 | 0.138 - 0.906 | -0.030 - -0.003 | 0.970 - 0.997 | 0.020 - 0.022 | -1.482 - -0.118 |
| 10 | Age:APOEe4/e4 | 0.165 - 0.973 | -0.043 - -0.001 | 0.958 - 0.999 | 0.028 - 0.031 | -1.389 - -0.034 |
| 11 | $Age^2$:APOEe3/e4 | 0.188 - 0.748 | 0.000 - 0.002 | 1.000 - 1.002 | 0.001 - 0.002 | 0.322 - 1.318 |
| 12 | $Age^2$:APOEe2 carriers | 0.323 - 0.934 | -0.002 - 0.002 | 0.998 - 1.002 | 0.002 - 0.004 | -0.400 - 0.988 |
| 13 | $Age^2$ | 0.351 - 0.945 | -0.000 - 0.001 | 1.000 - 1.001 | 0.001 - 0.001 | -0.298 - 0.933 |
| 14 | APOEe2 carriers | 0.372 - 0.867 | -0.342 - 0.073 | 0.710 - 1.076 | 0.318 - 0.383 | -0.893 - 0.230 |
| 15 | Age:APOEe2 carriers | 0.388 - 0.842 | -0.024 - 0.054 | 0.976 - 1.056 | 0.032 - 0.063 | -0.729 - 0.863 |
| 16 | $Age^2$:APOEe2/e4 | 0.465 - 0.925 | -0.020 - 0.003 | 0.980 - 1.003 | 0.007 - 0.029 | -0.731 - 0.456 |
| 17 | Age:APOEe2/e4 | 0.634 - 0.924 | 0.011 - 0.286 | 1.011 - 1.331 | 0.111 - 0.604 | 0.096 - 0.476 |

**Table S12**: Statistics for the maximal Cox model (PET+Demo+APOE) in the five PET cohorts predicting clinical AD diagnosis. Statistics are shown as ranges across cross-validation folds.

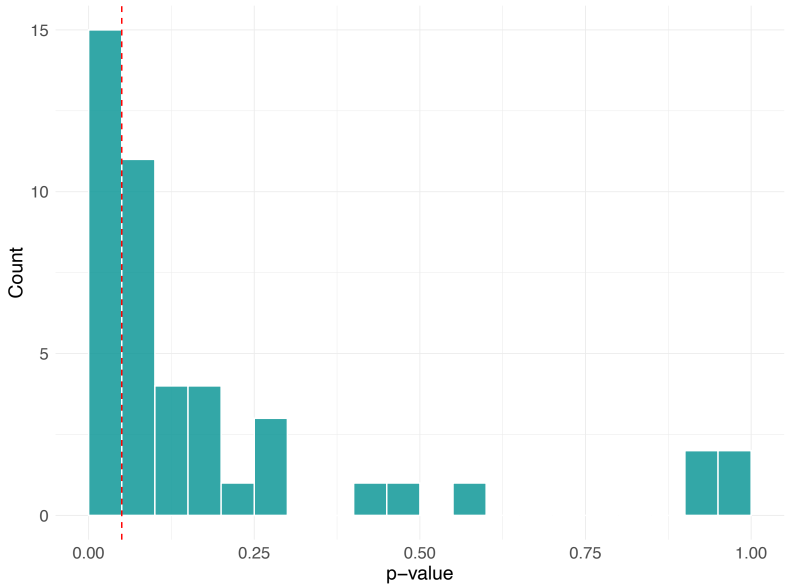


**Fig. S8**: Histogram of P-values comparing Demographics+APOE vs. PET+Demographics+APOE models across folds and time points for five PET cohorts (AIBL, ADNI, HABS, NACC, OASIS) predicting clinical AD diagnosis. 15/45 p-values were statistically significant.

| Fold | 2y | 3y | 4y | 5y | 6y | 7y | 8y | 9y | 10y |
|---|---|---|---|---|---|---|---|---|---|
| 1 | 0.1765 | 0.2568 | 0.0825 | **0.0459** | **0.0439** | 0.1367 | 0.0729 | 0.1145 | 0.0839 |
| 2 | 0.473 | 0.2914 | 0.1118 | **0.0261** | 0.1787 | 0.9918 | 0.9054 | 0.9124 | 1 |
| 3 | 0.5752 | 0.4036 | 0.1973 | 0.0818 | 0.0542 | 0.0655 | **0.0222** | **0.0028** | **9e-04** |
| 4 | 0.0933 | 0.0616 | **0.0176** | **0.0013** | **0.0011** | **0.0029** | **0.005** | **0.0076** | **0.0321** |
| 5 | 0.2591 | 0.0999 | 0.0701 | 0.1992 | 0.2287 | 0.1191 | 0.0869 | **0.0164** | **0.004** |

**Table S13**: P-values for time-varying AUROC with 95% confidence intervals for all folds and time points comparing PET+Demographics+APOE vs. Demographics+APOE across five PET cohorts (AIBL, ADNI, HABS, NACC, OASIS) predicting clinical AD diagnosis.

| Model | 2y | 3y | 4y | 5y | 6y | 7y | 8y | 9y | 10y |
|---|---|---|---|---|---|---|---|---|---|
| Demo | 0.55 (0.44-0.66) | 0.52 (0.42-0.61) | 0.51 (0.42-0.60) | 0.52 (0.43-0.61) | 0.55 (0.46-0.64) | 0.54 (0.45-0.63) | 0.57 (0.48-0.66) | 0.58 (0.49-0.67) | 0.58 (0.48-0.69) |
| Demo+APOE | 0.59 (0.48-0.70) | 0.55 (0.46-0.65) | 0.54 (0.45-0.64) | 0.56 (0.47-0.65) | 0.57 (0.48-0.66) | 0.56 (0.47-0.65) | 0.59 (0.50-0.69) | 0.59 (0.50-0.69) | 0.60 (0.49-0.70) |
| PET+Demo+APOE | 0.64 (0.52-0.76) | 0.61 (0.51-0.72) | 0.61 (0.51-0.71) | 0.63 (0.54-0.73) | 0.64 (0.54-0.73) | 0.62 (0.52-0.71) | 0.66 (0.57-0.75) | 0.67 (0.57-0.76) | 0.67 (0.57-0.77) |
| PET | 0.65 (0.51-0.78) | 0.64 (0.52-0.76) | 0.65 (0.54-0.75) | 0.66 (0.55-0.77) | 0.66 (0.55-0.77) | 0.62 (0.50-0.74) | 0.66 (0.55-0.78) | 0.68 (0.56-0.79) | 0.67 (0.56-0.79) |
| PET+Demo | 0.65 (0.54-0.76) | 0.62 (0.52-0.72) | 0.62 (0.52-0.72) | 0.64 (0.55-0.74) | 0.66 (0.57-0.75) | 0.63 (0.54-0.73) | 0.67 (0.59-0.76) | 0.68 (0.59-0.77) | 0.68 (0.58-0.77) |

**Table S14**: Time-varying AUROC with 95% confidence intervals for all models and time points for five PET cohorts predicting clinical AD diagnosis.

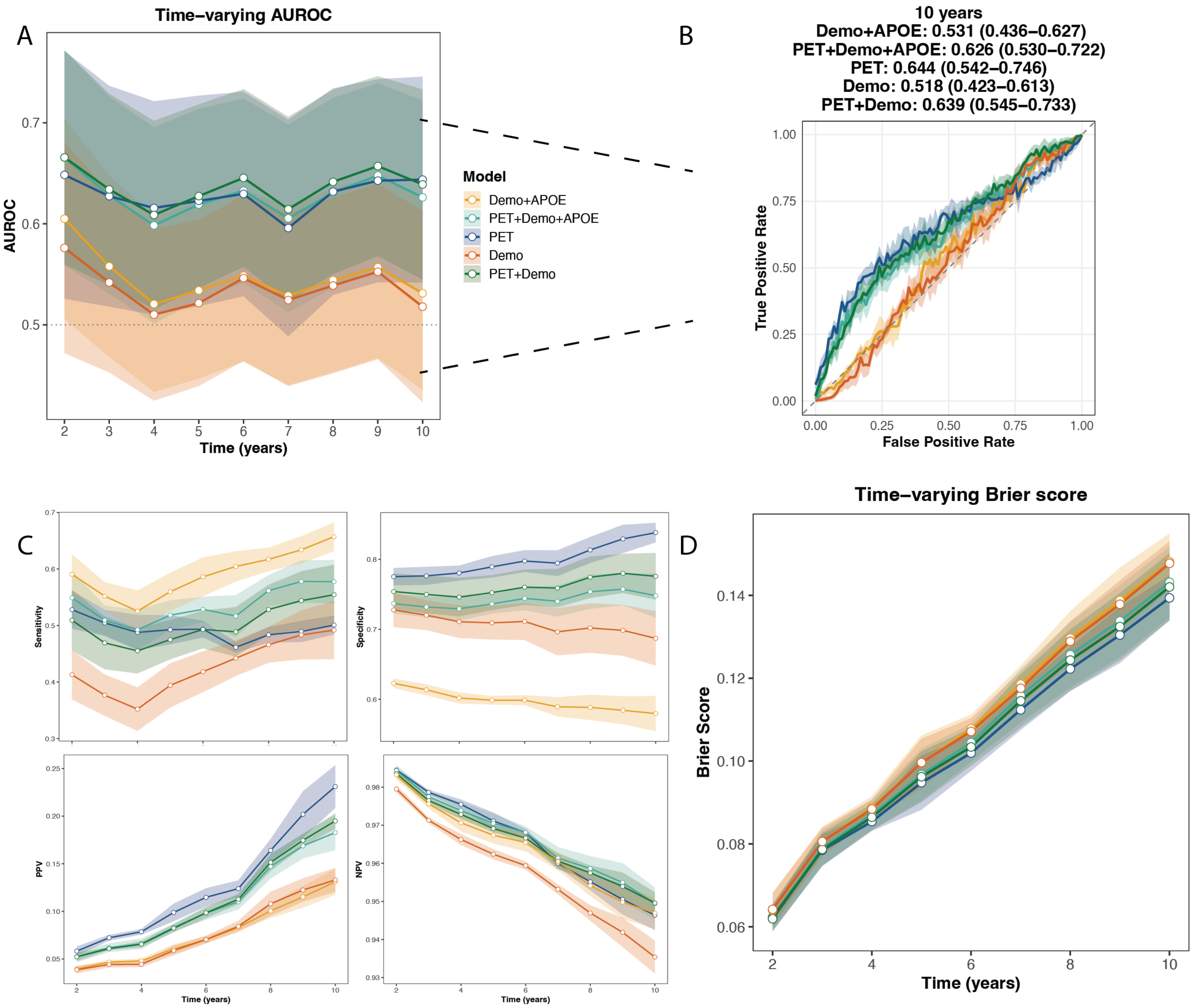


**Fig. S9**: Predicting all-cause dementia diagnosis across five PET cohorts using five models: 1) Demographics (age, sex, education) + APOE genotype, 2) Amyloid PET+demographics+APOE, 3) PET, 4) Demographics, 5) PET+Demographics. A) Time-varying AUROC for each year, with 95% confidence intervals. B) ROC curve for Year 10, which showed the largest difference in mean AUC between Demo and PET+Demo. C) Sensitivity, specificity, positive predictive value, and negative predictive value for all five models using optimal cutpoints by Youden's J statistic on the training set. D) Time-varying Brier score for each year, with standard deviations.

##### 6.2.4 Cerebrospinal fluid markers in NACC

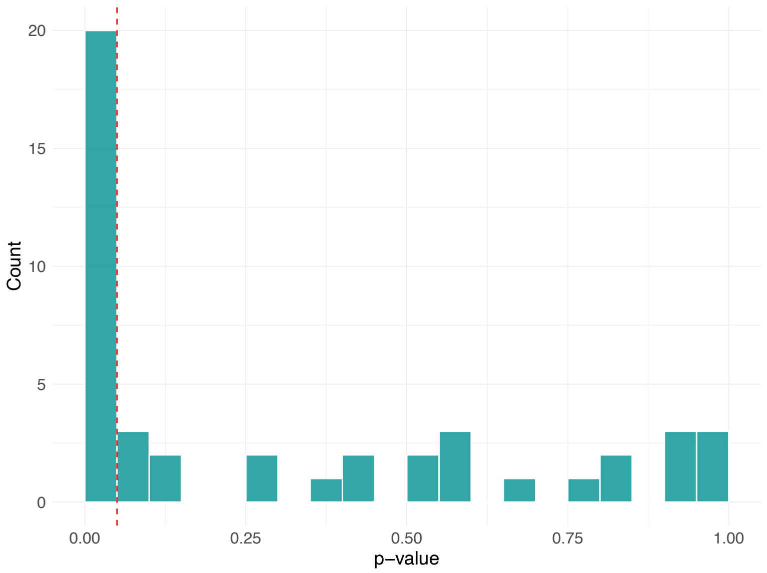


**Fig. S10**: Histogram of P-values comparing Demographics+APOE+Lancet vs. CSF+Demographics+APOE+Lancet models across folds and time points for CSF markers in NACC. 20/45 p-values were statistically significant.

| | term | P-value | Coef | exp(Coef) | SE(coef) | z |
|---|---|---|---|---|---|---|
| 1 | GDS score | 1.94e-10–4.03e-05 | 0.231–0.347 | 1.26–1.41 | 0.0543–0.0571 | 4.11–6.37 |
| 2 | Ratio pTau-181/Abeta | 2.42e-06–7.1e-05 | 0.232–0.497 | 1.26–1.64 | 0.0563–0.109 | 3.97–4.71 |
| 3 | Abeta | 1.3e-06–0.000149 | -0.521—0.415 | 0.594–0.66 | 0.104–0.109 | -4.84—3.79 |
| 4 | Age | 6.27e-06–0.000281 | 0.12–0.146 | 1.13–1.16 | 0.0298–0.037 | 3.63–4.52 |
| 5 | Age:APOEe4/e4 | 0.00179–0.107 | -0.136—0.0774 | 0.873–0.925 | 0.0414–0.052 | -3.12—1.61 |
| 6 | Total tau | 4.12e-05–0.151 | 0.153–0.513 | 1.17–1.67 | 0.0968–0.125 | 1.43–4.1 |
| 7 | Age$^2$ | 0.0148–0.175 | -0.00372—0.00188 | 0.996–0.998 | 0.00134–0.00156 | -2.44—1.36 |
| 8 | APOEe3/e4 | 0.00806–0.222 | 0.327–0.694 | 1.39–2 | 0.252–0.274 | 1.22–2.65 |
| 9 | Age$^2$:APOEe4/e4 | 0.0109–0.432 | 0.00252–0.00696 | 1–1.01 | 0.00229–0.00321 | 0.786–2.54 |
| 10 | Current depression treatment | 0.0131–0.307 | 0.208–0.497 | 1.23–1.64 | 0.2–0.213 | 1.02–2.48 |
| 11 | Age:APOEe3/e4 | 0.0624–0.556 | -0.0759—0.0247 | 0.927–0.976 | 0.0341–0.042 | -1.86—0.589 |
| 12 | Hypertension remote/inactive | 0.0224–0.702 | -1.04—0.175 | 0.354–0.84 | 0.406–0.47 | -2.28—0.383 |
| 13 | APOEe2/e4 | 0.0643–0.856 | -1.85—0.131 | 0.158–0.877 | 0.694–1.06 | -1.85—0.181 |
| 14 | Cigarettes last 30 days | 0.028–0.549 | 0.216–0.858 | 1.24–2.36 | 0.361–0.39 | 0.599–2.2 |
| 15 | Age$^2$:APOEe3/e4 | 0.00193–0.871 | -0.000381–0.0053 | 1–1.01 | 0.00171–0.00234 | -0.163–3.1 |
| 16 | BMI | 0.0711–0.398 | -0.151—0.0703 | 0.86–0.932 | 0.0808–0.0837 | -1.8—0.845 |
| 17 | Pack years | 0.158–0.527 | 0.08–0.174 | 1.08–1.19 | 0.122–0.133 | 0.632–1.41 |
| 18 | Education | 0.166–0.483 | -0.109—0.0553 | 0.897–0.946 | 0.0742–0.0788 | -1.39—0.701 |
| 19 | Cigarettes, 100+ in life | 0.0247–0.593 | -0.657—0.149 | 0.518–0.862 | 0.273–0.298 | -2.25—0.535 |
| 20 | Depression in last two years | 0.196–0.866 | -0.0359–0.286 | 0.965–1.33 | 0.204–0.225 | -0.168–1.29 |
| 21 | pTau-181 | 0.0232–0.97 | -0.041–0.238 | 0.96–1.27 | 0.102–0.129 | -0.349–2.27 |
| 22 | Alcohol once a week | 0.115–0.614 | 0.168–0.477 | 1.18–1.61 | 0.303–0.332 | 0.505–1.57 |
| 23 | APOEe4/e4 | 0.171–0.774 | 0.152–0.592 | 1.16–1.81 | 0.432–0.528 | 0.287–1.37 |
| 24 | Age$^2$:APOEe2/e4 | 0.299–0.5 | -0.012–0.00743 | 0.988–1.01 | 0.0058–0.0144 | -0.834–1.04 |
| 25 | Age$^2$:APOEe2 carriers | 0.118–0.685 | -0.00788—0.00171 | 0.992–0.998 | 0.00423–0.00504 | -1.56—0.406 |
| 26 | Age:APOEe2 carriers | 0.171–0.799 | 0.0227–0.154 | 1.02–1.17 | 0.0892–0.112 | 0.255–1.37 |
| 27 | Alcohol once a month | 0.304–0.607 | -1.07–0.385 | 0.345–1.47 | 0.742–1.04 | -1.03–0.515 |
| 28 | Abeta assay (Luminex) | 0.103–0.707 | -0.516—0.12 | 0.597–0.887 | 0.306–0.319 | -1.63—0.375 |
| 29 | TBI | 0.156–0.863 | -0.0378–0.306 | 0.963–1.36 | 0.21–0.223 | -0.173–1.42 |
| 30 | Depression | 0.271–0.87 | -0.0349–0.238 | 0.966–1.27 | 0.21–0.216 | -0.164–1.1 |
| 31 | Abeta assay (other) | 0.229–0.961 | 0.0133–0.31 | 1.01–1.36 | 0.257–0.272 | 0.0488–1.2 |
| 32 | Hypertension | 0.228–0.86 | 0.0398–0.263 | 1.04–1.3 | 0.218–0.226 | 0.176–1.21 |
| 33 | Diabetes | 0.0573–0.958 | 0.0181–0.66 | 1.02–1.94 | 0.34–0.379 | 0.0531–1.9 |
| 34 | Hypertension combo meds | 0.286–0.976 | -0.252–0.315 | 0.778–1.37 | 0.295–0.344 | -0.764–1.07 |
| 35 | Hearing loss | 0.176–0.939 | -0.288–0.0487 | 0.75–1.05 | 0.209–0.222 | -1.35–0.231 |
| 36 | Diabetes medication | 0.373–0.967 | -0.123–0.377 | 0.884–1.46 | 0.38–0.423 | -0.313–0.891 |
| 37 | Hypertension medication | 0.347–0.929 | -0.205–0.0229 | 0.814–1.02 | 0.213–0.222 | -0.941–0.103 |
| 38 | Age:APOEe2/e4 | 0.21–0.951 | -0.0384–0.312 | 0.962–1.37 | 0.0834–0.249 | -0.461–1.25 |
| 39 | Smoking years | 0.374–0.936 | -0.0406–0.0897 | 0.96–1.09 | 0.0924–0.104 | -0.425–0.889 |
| 40 | Depression over two years ago | 0.508–0.964 | -0.089–0.132 | 0.915–1.14 | 0.191–0.199 | -0.465–0.662 |
| 41 | APOEe2 carriers | 0.689–0.827 | -0.188–0.191 | 0.829–1.21 | 0.478–0.576 | -0.326–0.4 |
| 42 | Hearing aid | 0.341–0.938 | -0.0201–0.201 | 0.98–1.22 | 0.21–0.228 | -0.0884–0.953 |
| 43 | Female | 0.631–0.992 | -0.0807–0.00554 | 0.922–1.01 | 0.163–0.171 | -0.481–0.034 |
| 44 | Diabetes remote/inactive | 0.993–0.995 | -14.1—13.5 | 7.62e-07–1.39e-06 | 1650–2010 | -0.00854—0.00672 |

**Table S15**: Statistics for the maximal Cox model (CSF+Demo+APOE+Lancet) in the NACC cohort predicting clinical AD diagnosis. Statistics are shown as ranges across cross-validation folds.

| Fold | 2y | 3y | 4y | 5y | 6y | 7y | 8y | 9y | 10y |
|---|---|---|---|---|---|---|---|---|---|
| 1 | 0.0522 | **0.0092** | **0** | **1e-04** | **0.023** | 0.0513 | 0.5245 | 0.5842 | 0.5745 |
| 2 | 0.9997 | 0.9094 | 0.8078 | 1 | 0.9993 | 0.8245 | 0.9197 | 0.9061 | 0.6555 |
| 3 | 0.1385 | 0.7644 | 0.518 | 0.2662 | **0.028** | **0.0308** | **0.013** | **0.0198** | **0.0038** |
| 4 | **3e-04** | **0.0013** | **0.0169** | **0.0242** | **0.0118** | **0.016** | 0.1133 | 0.3955 | 0.4299 |
| 5 | **0.0085** | 0.4336 | 0.5675 | 0.2905 | 0.0825 | **0.0458** | **0.0143** | **0.0046** | **0.0017** |

**Table S16**: P-values for time-varying AUROC with 95% confidence intervals for all folds and time points comparing Demographics+APOE+Lancet vs. CSF+Demographics+APOE+Lancet predicting clinical AD diagnosis.

| Model | 2y | 3y | 4y | 5y | 6y | 7y | 8y | 9y | 10y |
|---|---|---|---|---|---|---|---|---|---|
| Demo | 0.51 (0.31-0.70) | 0.50 (0.35-0.66) | 0.51 (0.37-0.64) | 0.52 (0.40-0.65) | 0.51 (0.40-0.61) | 0.51 (0.40-0.61) | 0.51 (0.42-0.61) | 0.50 (0.40-0.60) | 0.50 (0.40-0.60) |
| Demo+APOE | 0.56 (0.39-0.74) | 0.55 (0.41-0.69) | 0.55 (0.43-0.67) | 0.57 (0.46-0.69) | 0.55 (0.45-0.65) | 0.54 (0.44-0.64) | 0.54 (0.45-0.64) | 0.53 (0.43-0.62) | 0.52 (0.42-0.61) |
| Demo+Lancet | 0.59 (0.39-0.80) | 0.65 (0.50-0.80) | 0.65 (0.53-0.77) | 0.64 (0.53-0.75) | 0.62 (0.51-0.72) | 0.62 (0.52-0.72) | 0.65 (0.55-0.74) | 0.63 (0.53-0.72) | 0.62 (0.52-0.71) |
| Demo+APOE+Lancet | 0.63 (0.44-0.82) | 0.67 (0.53-0.80) | 0.66 (0.55-0.78) | 0.67 (0.57-0.77) | 0.64 (0.54-0.74) | 0.63 (0.53-0.72) | 0.65 (0.55-0.74) | 0.63 (0.53-0.72) | 0.62 (0.52-0.71) |
| Lancet | 0.64 (0.49-0.80) | 0.69 (0.57-0.81) | 0.68 (0.57-0.80) | 0.67 (0.57-0.78) | 0.65 (0.55-0.75) | 0.64 (0.54-0.74) | 0.66 (0.56-0.75) | 0.64 (0.54-0.74) | 0.63 (0.54-0.73) |
| CSF+Demo | 0.71 (0.52-0.90) | 0.68 (0.54-0.83) | 0.69 (0.56-0.81) | 0.68 (0.57-0.80) | 0.67 (0.57-0.78) | 0.66 (0.56-0.77) | 0.67 (0.56-0.77) | 0.66 (0.56-0.76) | 0.66 (0.56-0.76) |
| CSF+Demo+APOE | 0.73 (0.56-0.91) | 0.69 (0.54-0.83) | 0.69 (0.56-0.82) | 0.70 (0.59-0.81) | 0.69 (0.58-0.79) | 0.67 (0.57-0.78) | 0.67 (0.57-0.77) | 0.65 (0.55-0.75) | 0.65 (0.55-0.75) |
| CSF+Demo+Lancet | 0.74 (0.56-0.92) | 0.76 (0.62-0.89) | 0.77 (0.65-0.88) | 0.75 (0.65-0.86) | 0.73 (0.63-0.83) | 0.73 (0.63-0.82) | 0.74 (0.65-0.83) | 0.72 (0.63-0.81) | 0.71 (0.62-0.80) |
| CSF+Demo+APOE+Lancet | 0.75 (0.58-0.93) | 0.75 (0.61-0.89) | 0.75 (0.63-0.87) | 0.75 (0.65-0.85) | 0.73 (0.63-0.83) | 0.72 (0.63-0.82) | 0.73 (0.63-0.82) | 0.70 (0.61-0.79) | 0.70 (0.60-0.79) |
| CSF | 0.78 (0.60-0.95) | 0.75 (0.62-0.89) | 0.75 (0.63-0.87) | 0.75 (0.65-0.85) | 0.75 (0.65-0.84) | 0.73 (0.63-0.83) | 0.73 (0.64-0.83) | 0.71 (0.61-0.81) | 0.72 (0.62-0.82) |

**Table S17**: Time-varying AUROC with 95% confidence intervals for all models and time points for CSF in NACC with AD as the outcome.

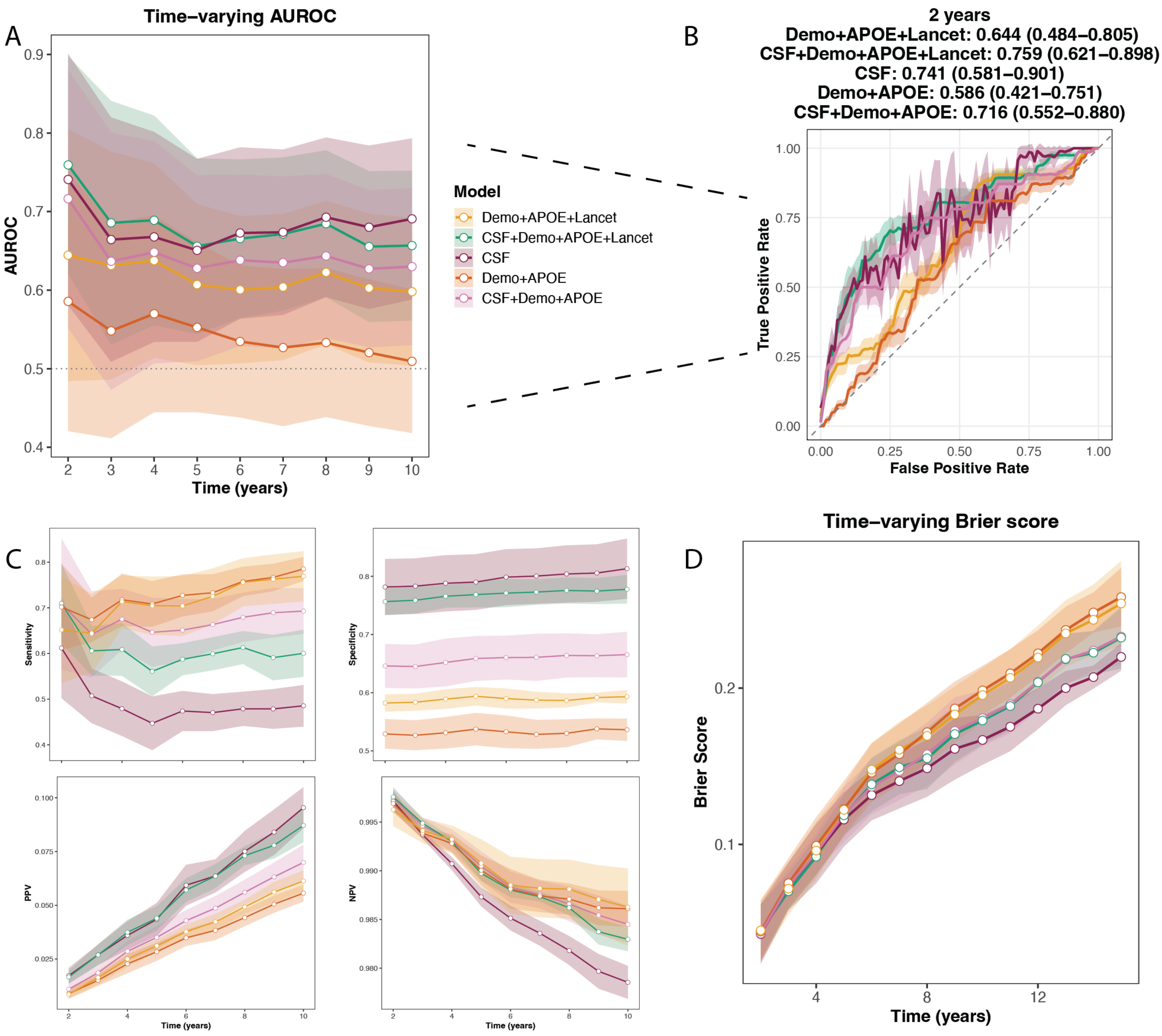


**Fig. S11**: Predicting all-cause dementia diagnosis in NACC using CSF Abeta42, pTau-181, and total tau, showing the following models: 1) Demographics+Lancet, 2) CSF+Demo+Lancet, 3) CSF, 4) Demographics, 5) CSF+Demographics. A) Time-varying AUROC for each year, with 95% confidence intervals. B) ROC curve for Year 2, which showed the largest difference in mean AUC between Demo+Lancet and CSF+Demo+Lancet. C) Sensitivity, specificity, positive predictive value, and negative predictive value for all five models using optimal cutpoints by Youden's J statistic on the training set. D) Time-varying Brier score for each year, with standard deviations.

##### 6.2.5 Plasma proteomics in the UK Biobank

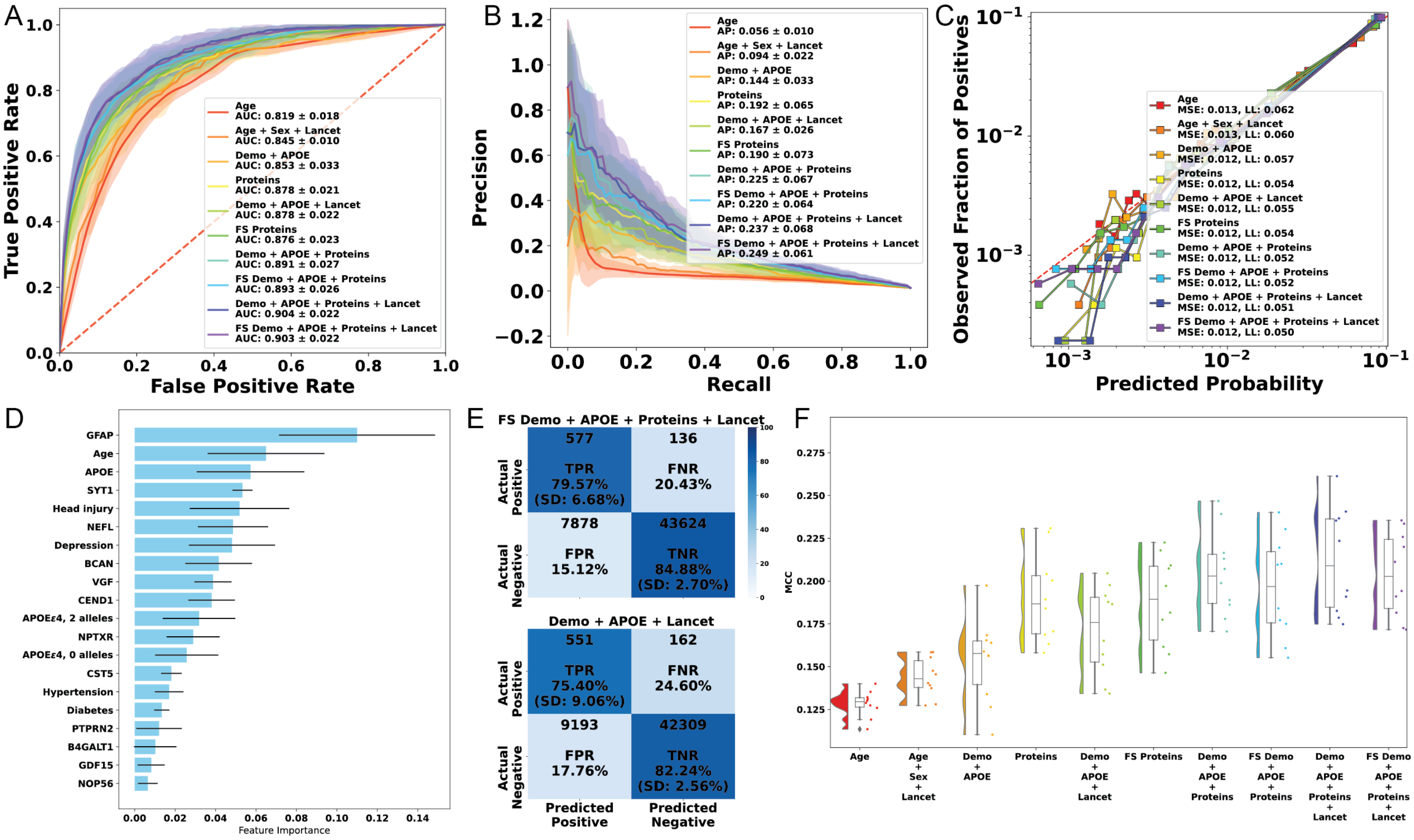


**Fig. S12**: Predicting Alzheimer's dementia with blood plasma proteomics with no age cutoff. A) Mean ROC curves. B) Mean precision-recall curves. C) Calibration curves (all folds combined). D) Mean feature importance plots. E) Confusion matrices for Demo+Lancet and Feature Selected Demo+Lancet+Proteins experiments. F) Raincloud plots of Matthews correlation coefficient.

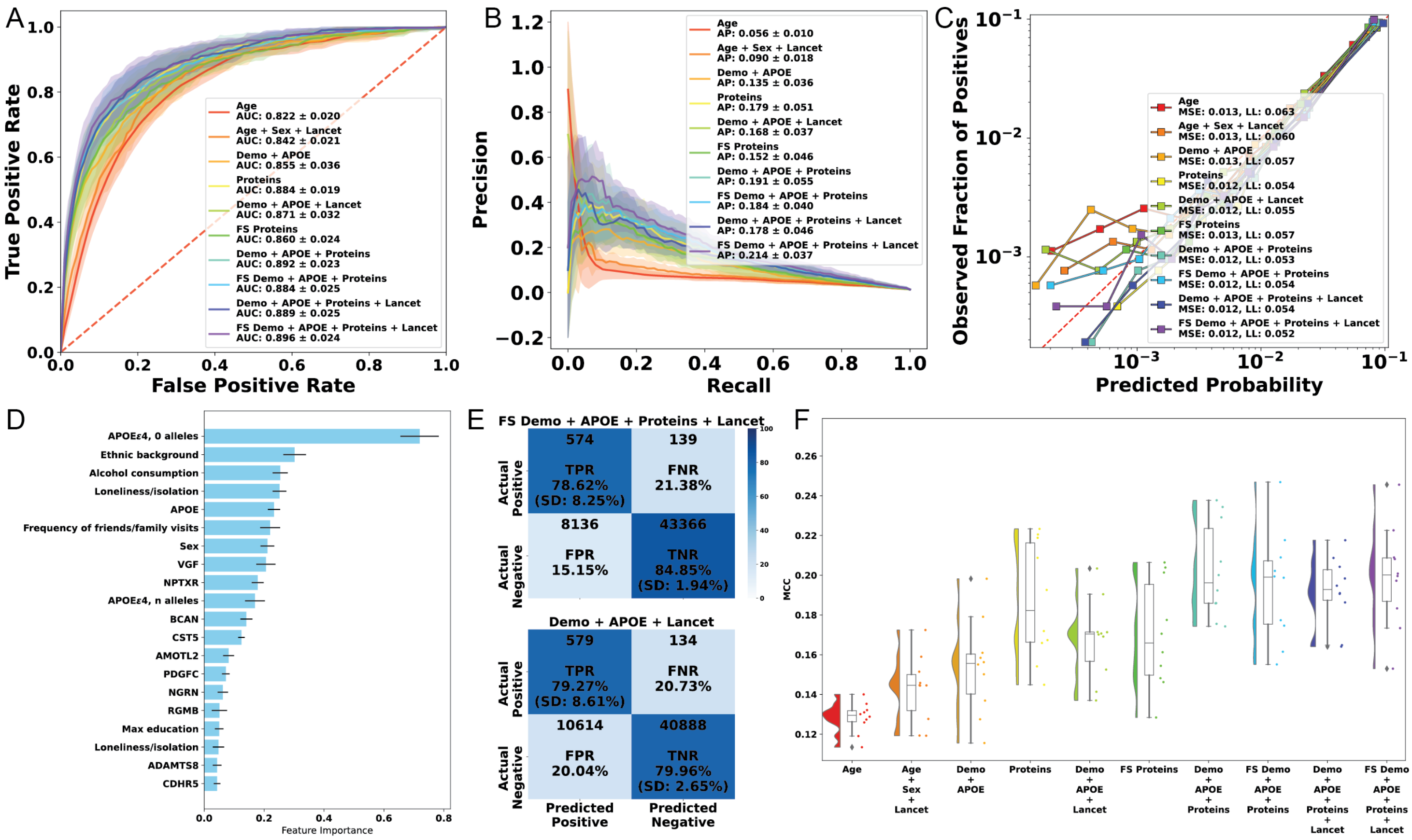


**Fig. S13**: Predicting Alzheimer's dementia with plasma proteomics, using logistic regression instead of LightGBM. A) Mean ROC curves. B) Mean precision-recall curves. C) Calibration curves (all folds combined). D) Mean feature importance plots. E) Confusion matrices for Demo+Lancet and Feature Selected Demo+Lancet+Proteins experiments. F) Raincloud plots of Matthews correlation coefficient.

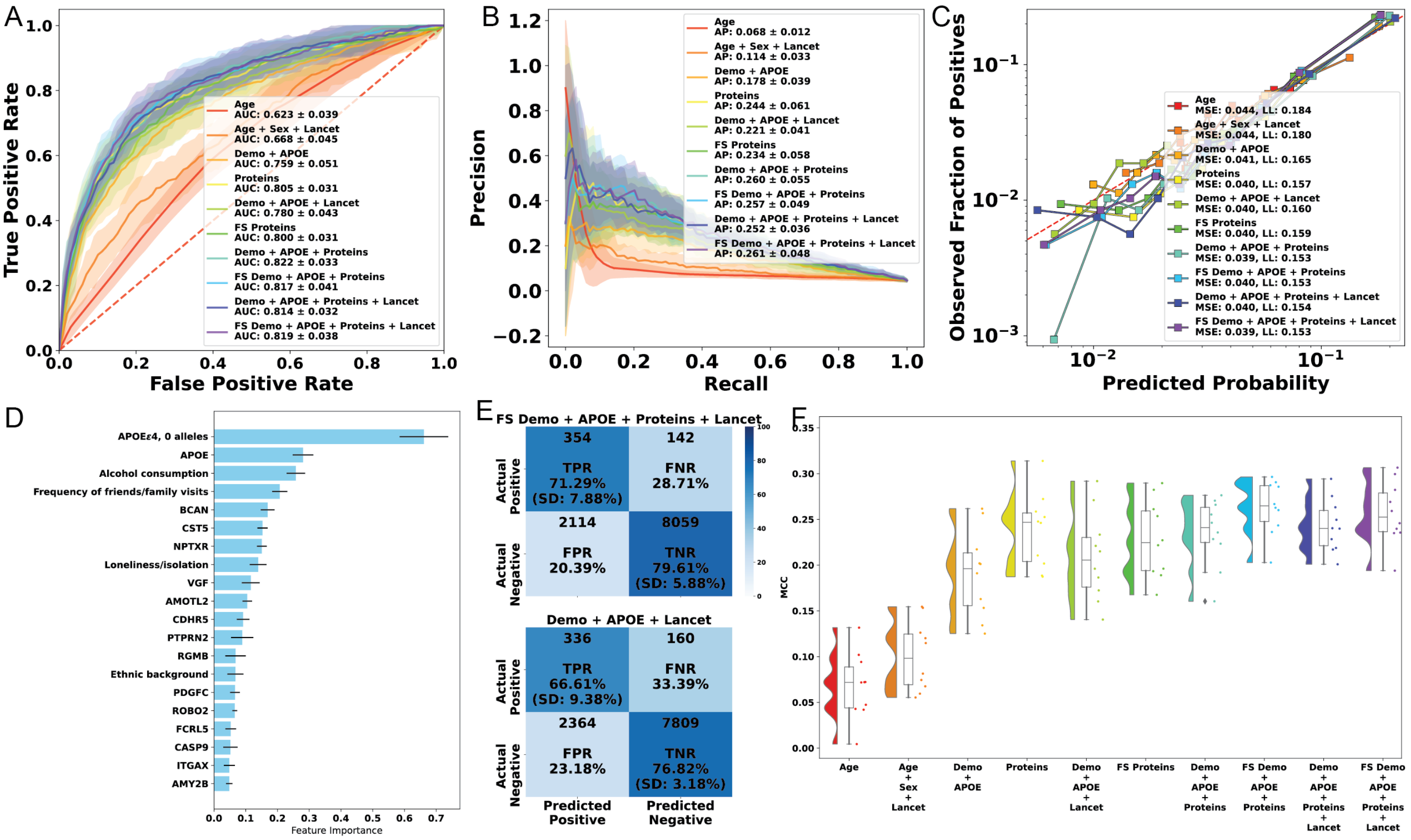


**Fig. S14**: Predicting Alzheimer's dementia with plasma proteomics, only including participants aged 65 and older, with logistic regression instead of LightGBM. A) Mean ROC curves. B) Mean precision-recall curves. C) Calibration curves (all folds combined). D) Mean feature importance plots. E) Confusion matrices for Demo+Lancet and Feature Selected Demo+Lancet+Proteins experiments. F) Raincloud plots of Matthews correlation coefficient.

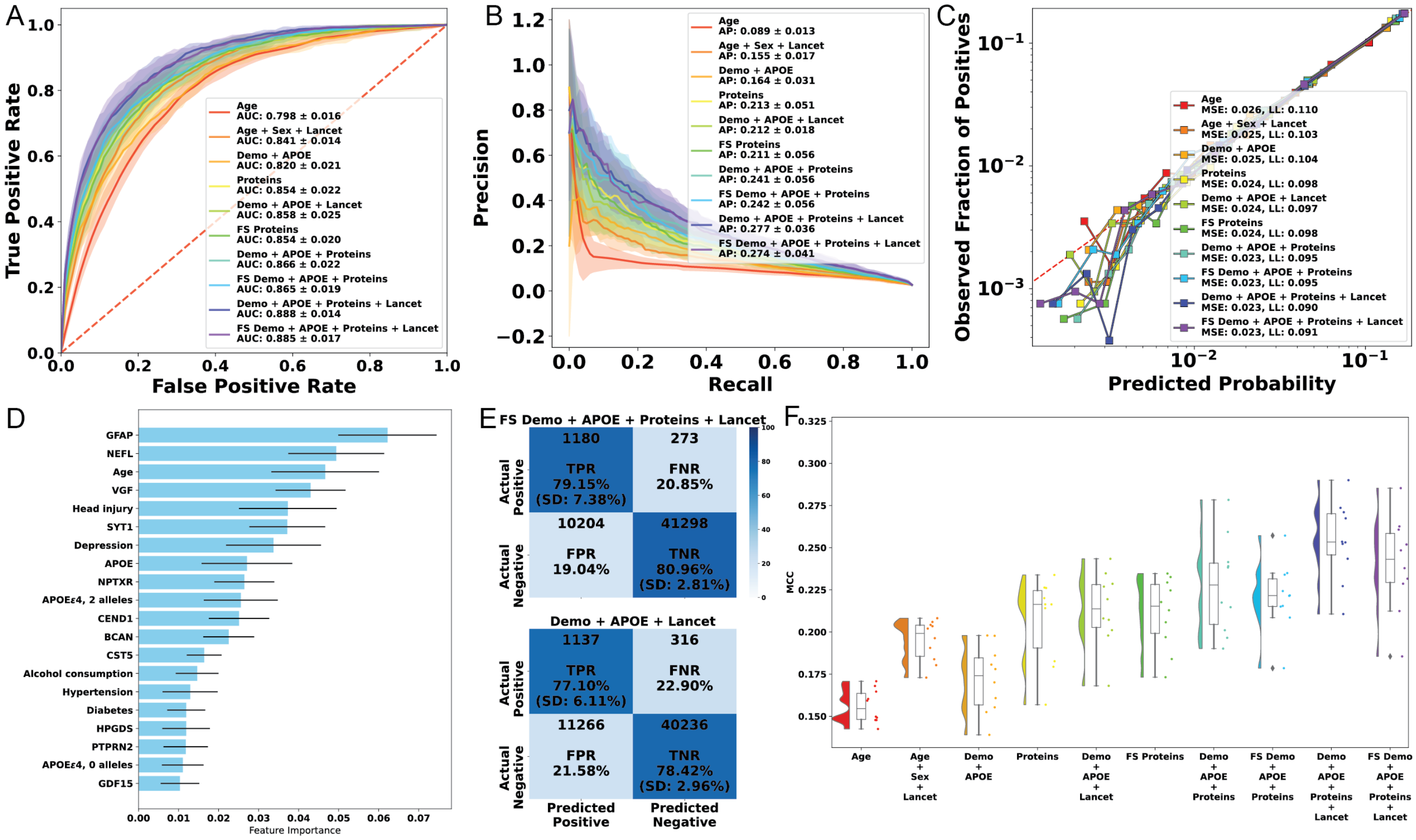

**Fig. S15**: Predicting all-cause dementia with plasma proteomics. A) Mean ROC curves. B) Mean precision-recall curves. C) Calibration curves (all folds combined). D) Mean feature importance plots. E) Confusion matrices for Demo+Lancet and Feature Selected Demo+Lancet+Proteins experiments. F) Raincloud plots of Matthews correlation coefficient.

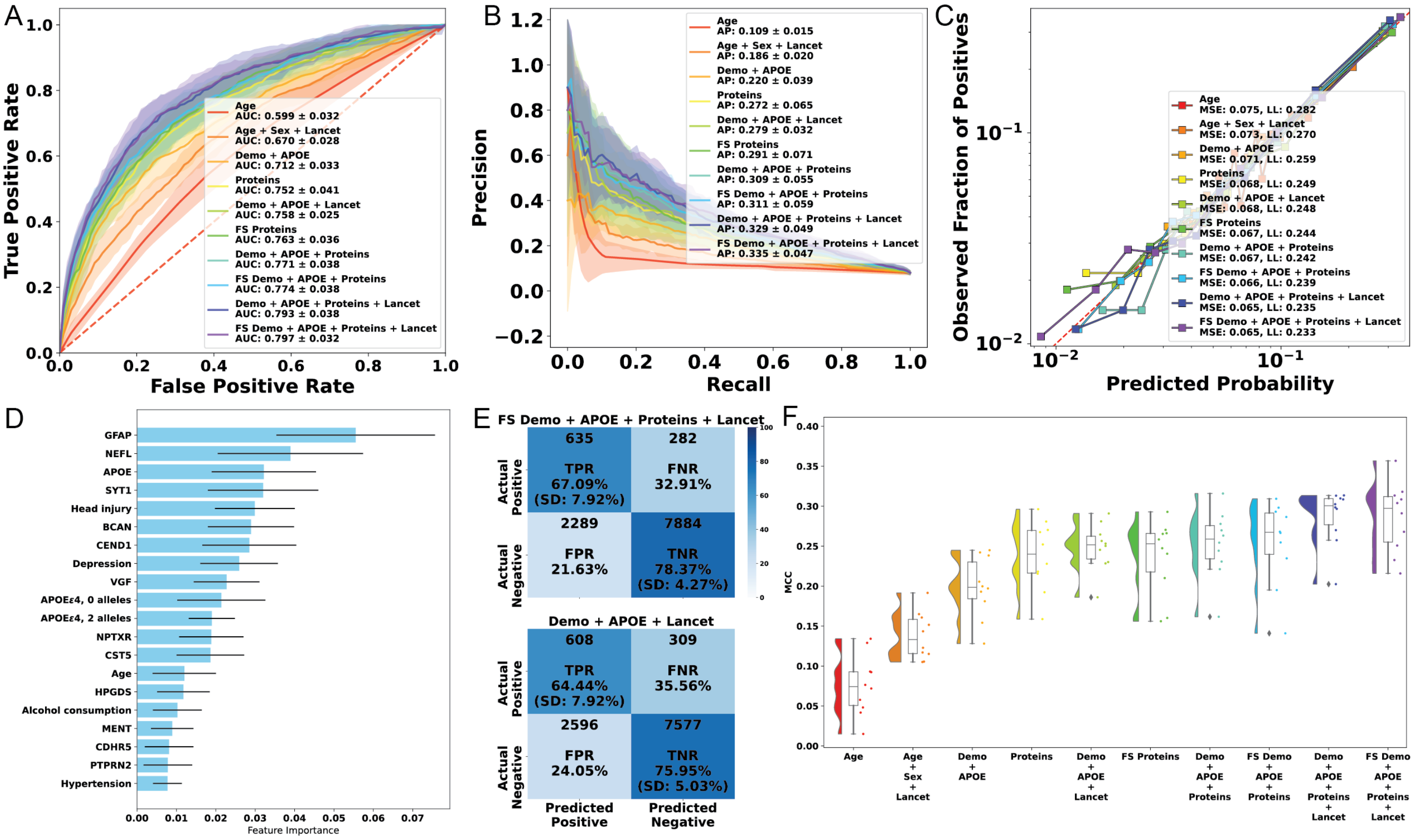


**Fig. S16**: Predicting all-cause dementia with plasma proteomics, only including participants aged 65 and older. A) Mean ROC curves. B) Mean precision-recall curves. C) Calibration curves (all folds combined). D) Mean feature importance plots. E) Confusion matrices for Demo+Lancet and Feature Selected Demo+Lancet+Proteins experiments. F) Raincloud plots of Matthews correlation coefficient.

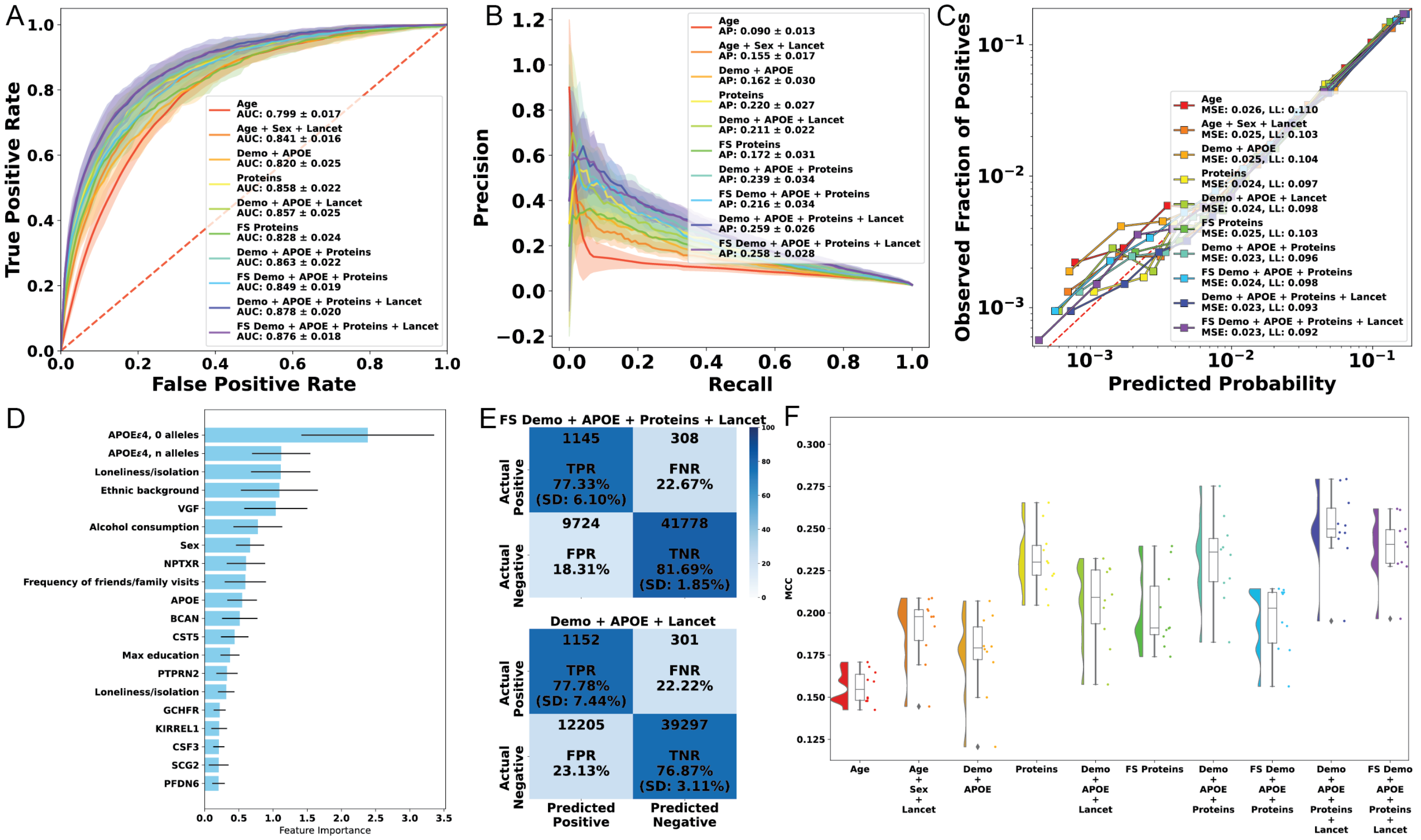


**Fig. S17**: Predicting all-cause dementia with plasma proteomics, using logit regression instead of LightGBM. A) Mean ROC curves. B) Mean precision-recall curves. C) Calibration curves (all folds combined). D) Mean feature importance plots. E) Confusion matrices for Demo+Lancet and Feature Selected Demo+Lancet+Proteins experiments. F) Raincloud plots of Matthews correlation coefficient.

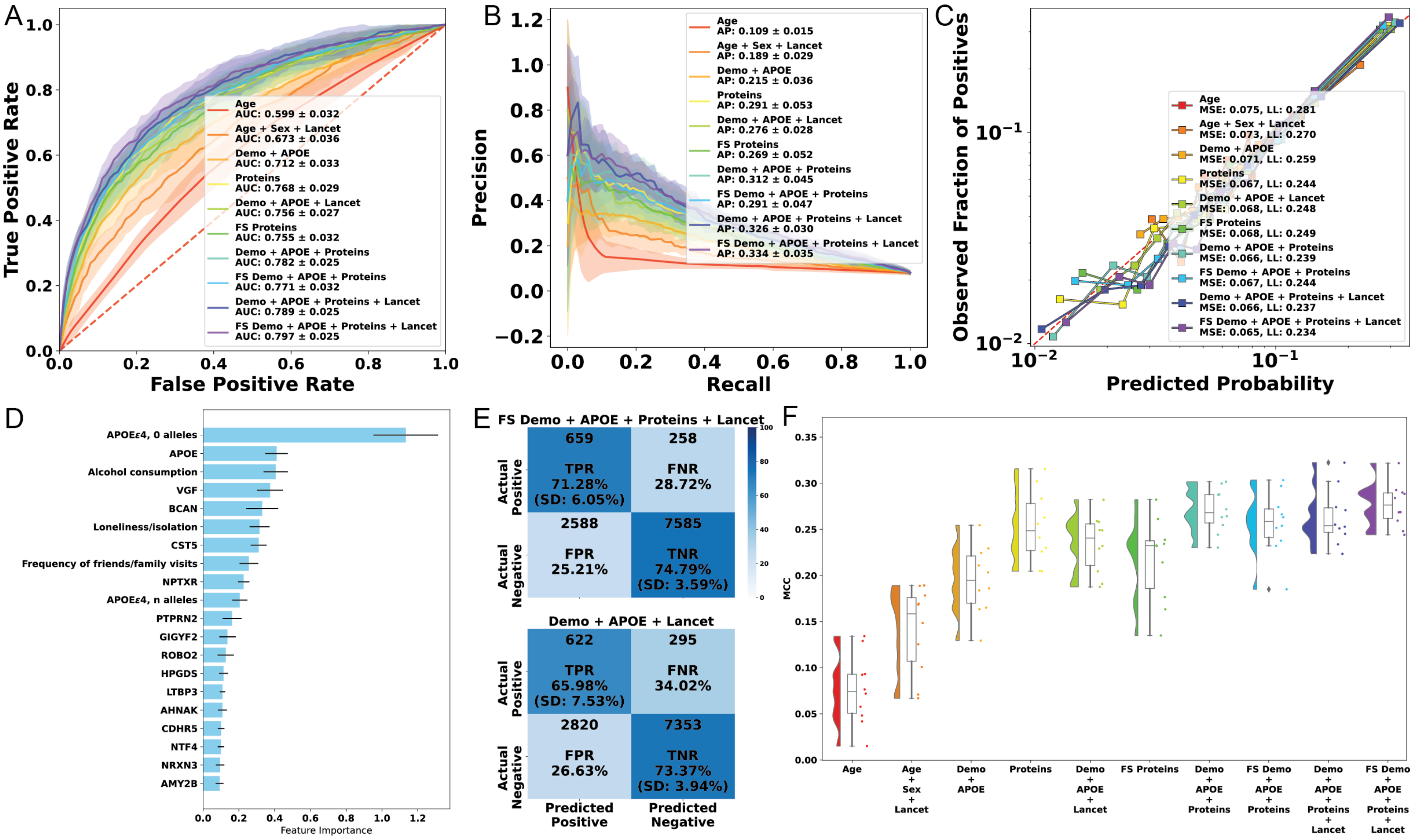

**Fig. S18**: Predicting all-cause dementia with plasma proteomics, only including participants aged 65 and older, with logit regression. A) Mean ROC curves. B) Mean precision-recall curves. C) Calibration curves (all folds combined). D) Mean feature importance plots. E) Confusion matrices for Demo+Lancet and Feature Selected Demo+Lancet+Proteins experiments. F) Raincloud plots of Matthews correlation coefficient.

##### 6.2.6 Brain imaging-derived phenotypes in the UK Biobank

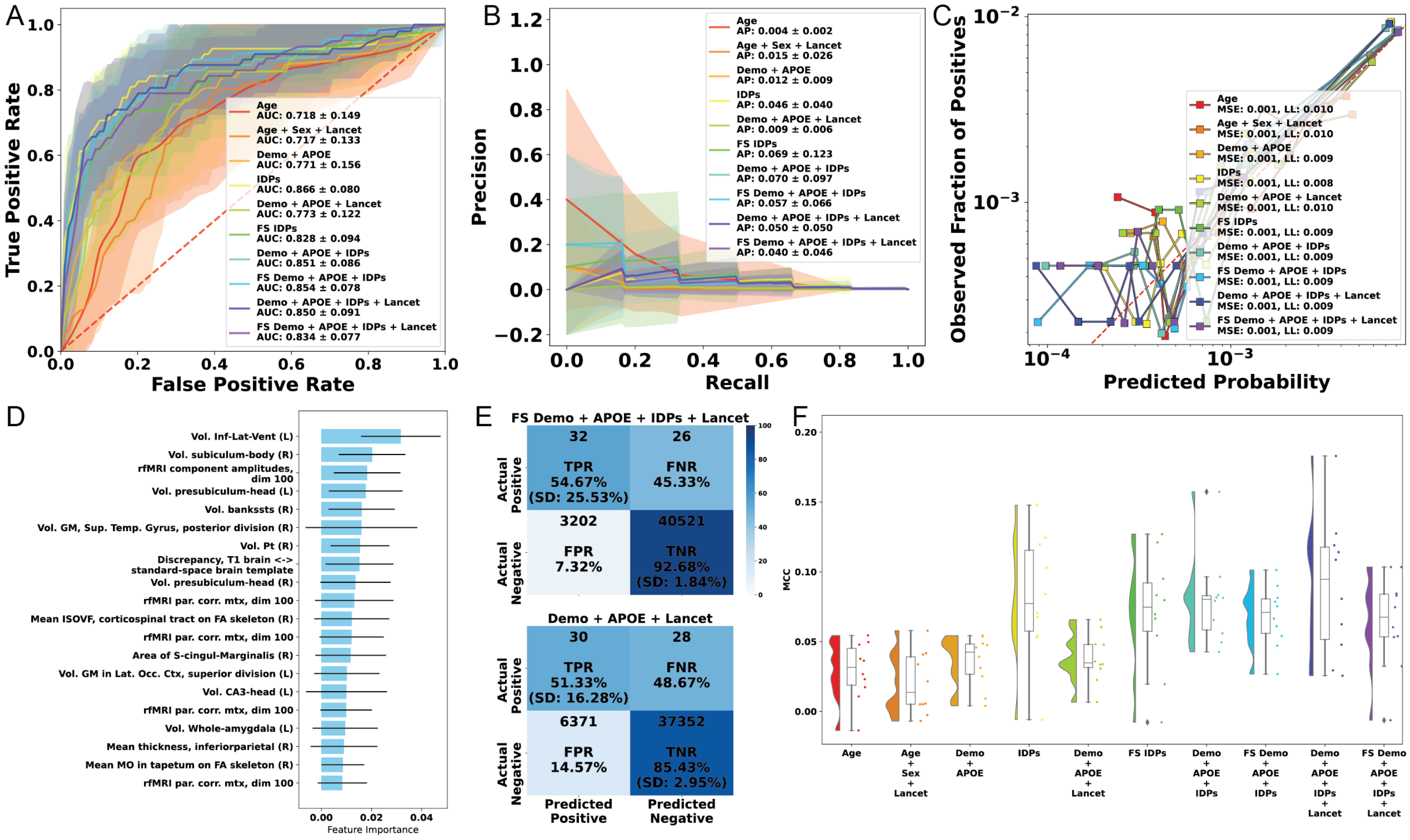


**Fig. S19**: Predicting Alzheimer's dementia with imaging-derived phenotypes with no age cutoff. A) Mean ROC curves. B) Mean precision-recall curves. C) Calibration curves (all folds combined). D) Mean feature importance plots. E) Confusion matrices for Demo+Lancet and Feature Selected Demo+Lancet+IDPs experiments. F) Raincloud plots of Matthews correlation coefficient.

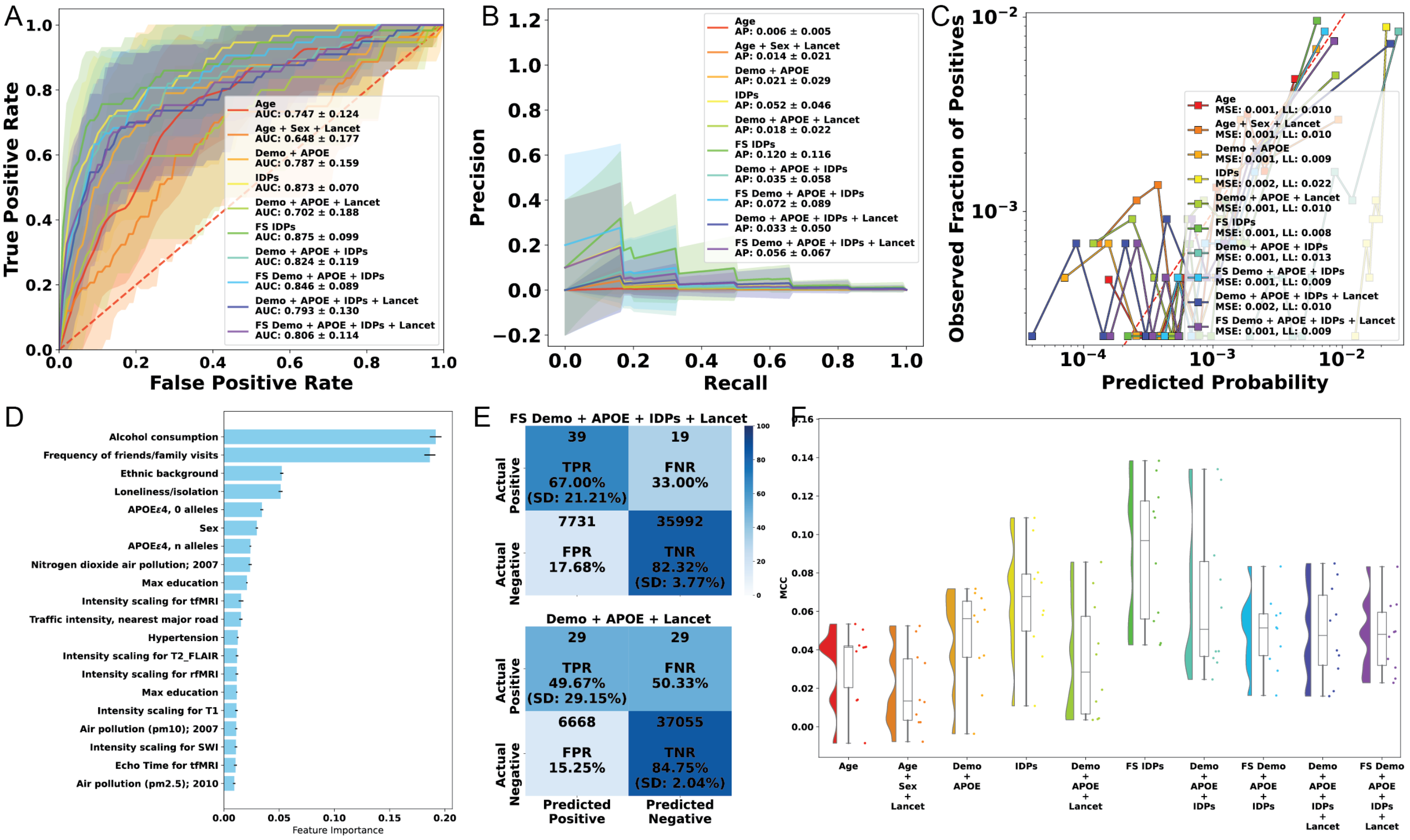


**Fig. S20**: Predicting Alzheimer's dementia with brain imaging-derived phenotypes, using logistic regression instead of LightGBM. A) Mean ROC curves. B) Mean precision-recall curves. C) Calibration curves (all folds combined). D) Mean feature importance plots. E) Confusion matrices for Demo+Lancet and Feature Selected Demo+Lancet+IDPs experiments. F) Raincloud plots of Matthews correlation coefficient.

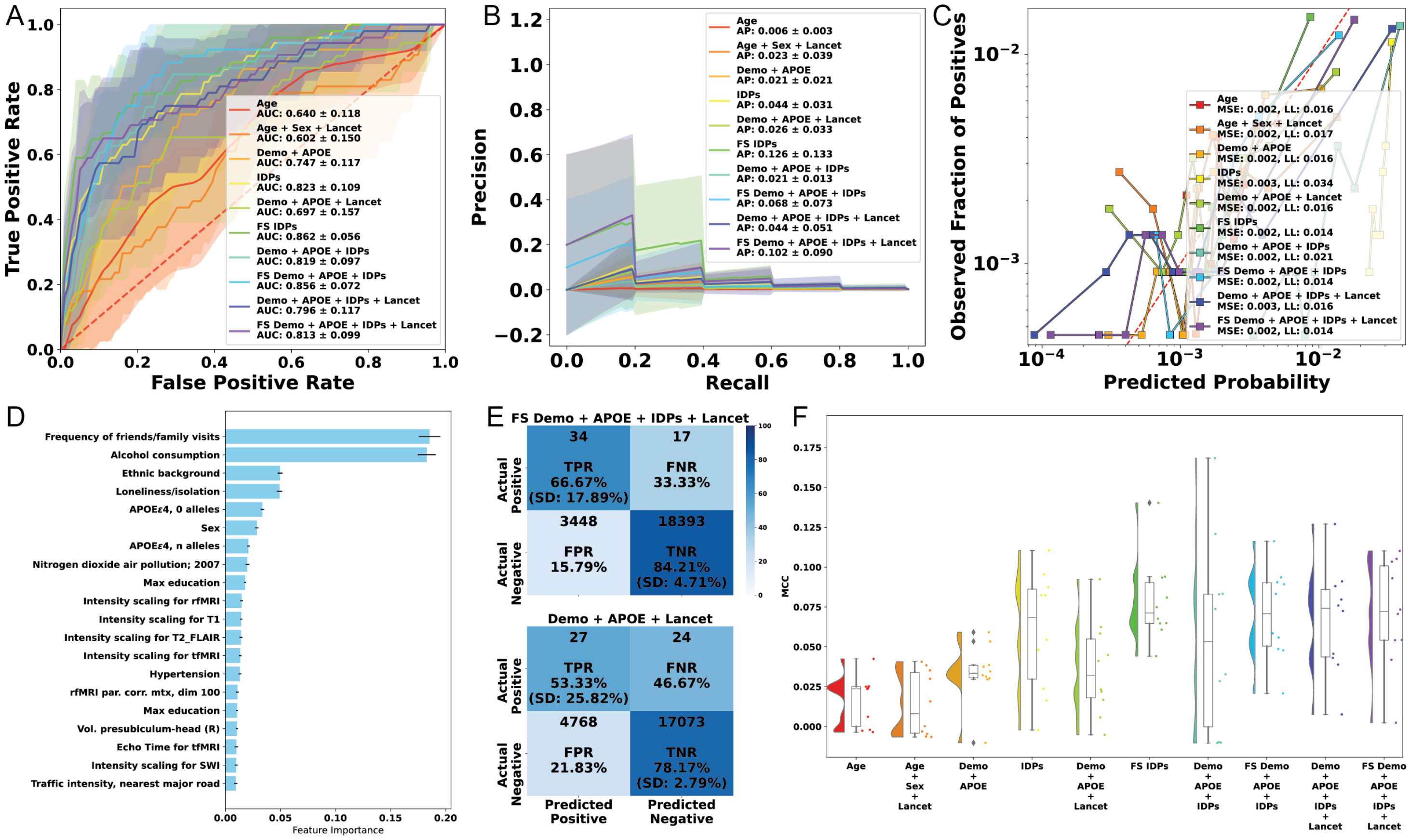


**Fig. S21**: Predicting Alzheimer's dementia with brain imaging-derived phenotypes, only including participants aged 65 and older, with logistic regression instead of LightGBM. A) Mean ROC curves. B) Mean precision-recall curves. C) Calibration curves (all folds combined). D) Mean feature importance plots. E) Confusion matrices for Demo+Lancet and Feature Selected Demo+Lancet+IDPs experiments. F) Raincloud plots of Matthews correlation coefficient.

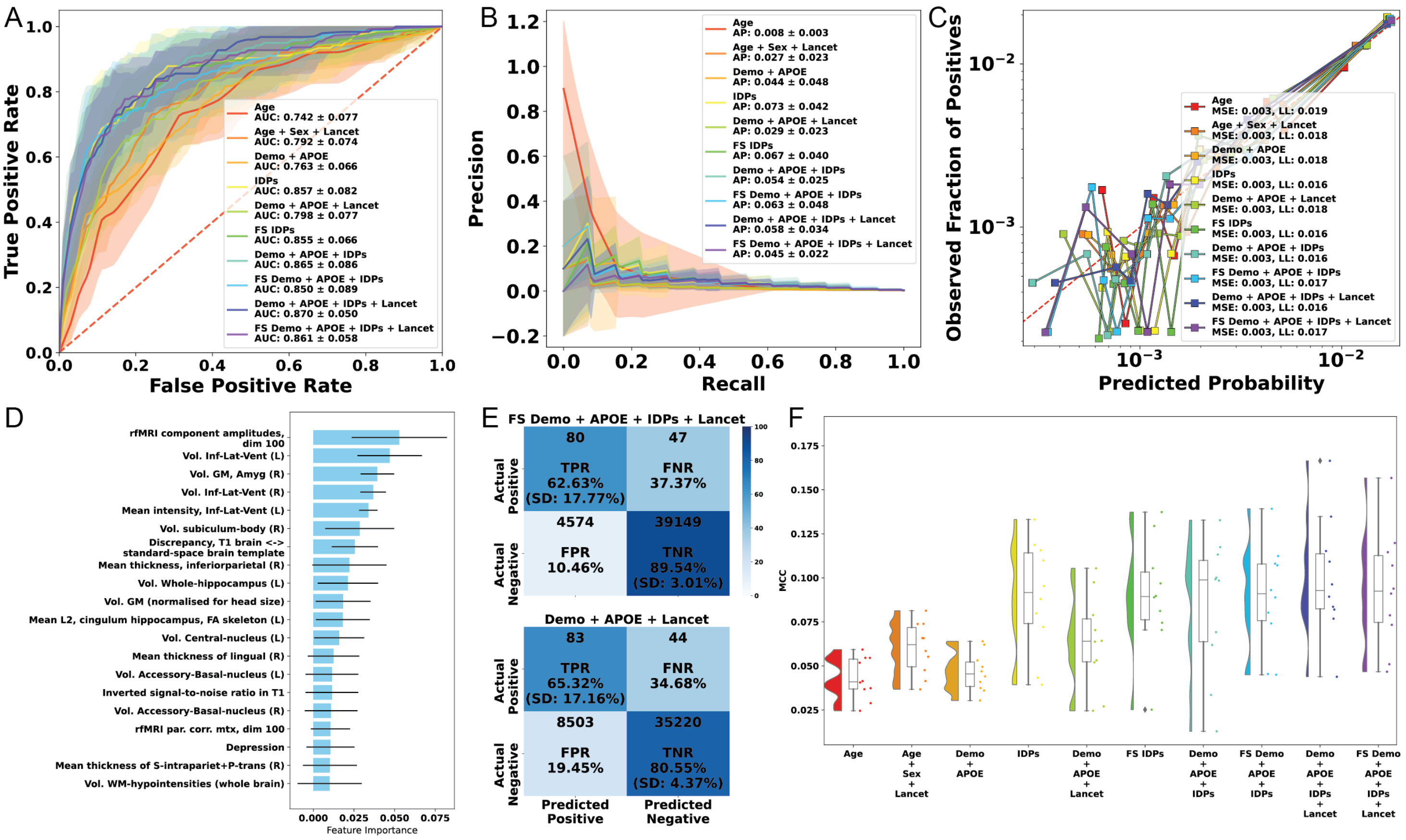


**Fig. S22**: Predicting all-cause dementia with brain imaging-derived phenotypes. A) Mean ROC curves. B) Mean precision-recall curves. C) Calibration curves (all folds combined). D) Mean feature importance plots. E) Confusion matrices for Demo+Lancet and Feature Selected Demo+Lancet+IDPs experiments. F) Raincloud plots of Matthews correlation coefficient.

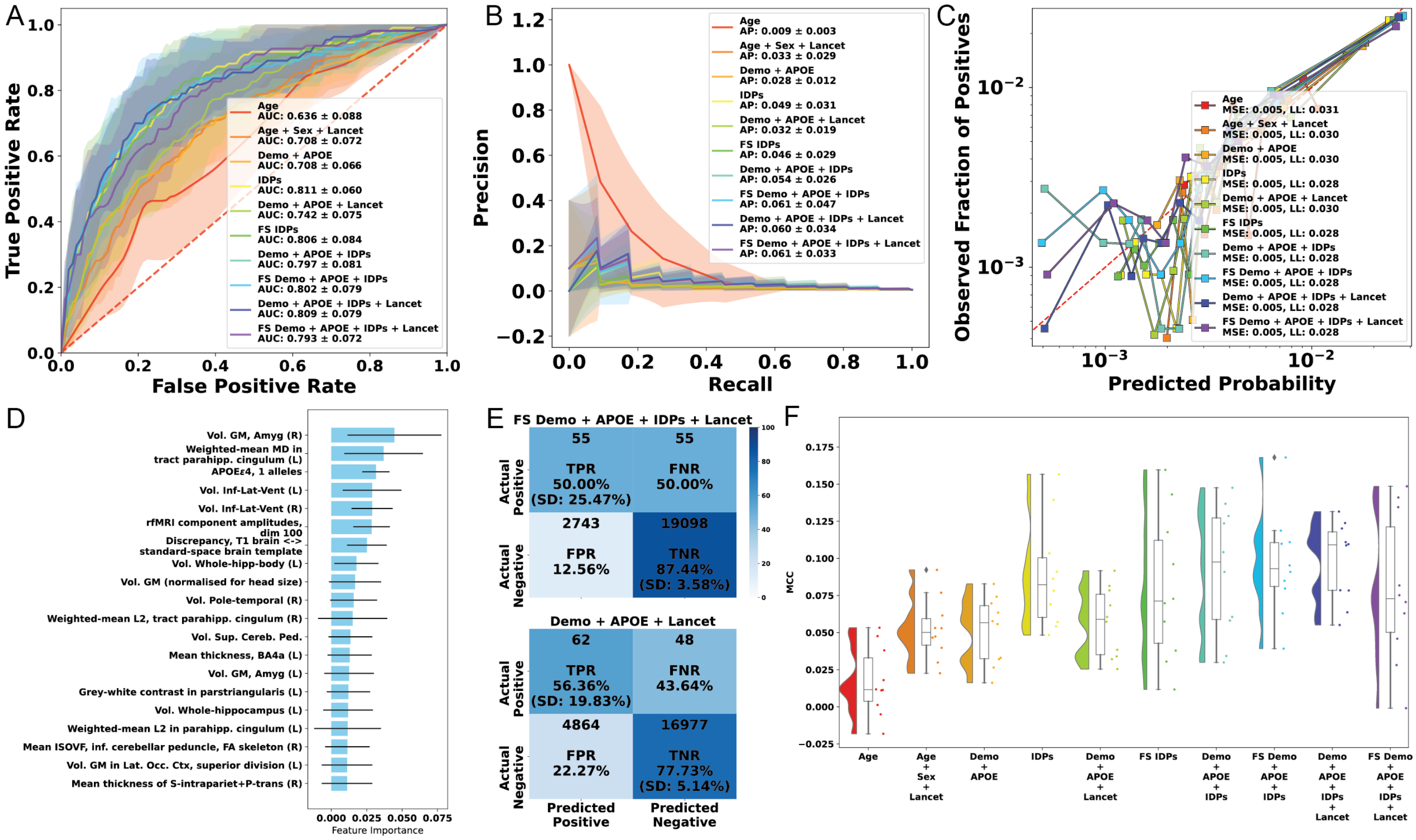


**Fig. S23**: Predicting all-cause dementia with brain imaging-derived phenotypes, only including participants aged 65 and older. A) Mean ROC curves. B) Mean precision-recall curves. C) Calibration curves (all folds combined). D) Mean feature importance plots. E) Confusion matrices for Demo+Lancet and Feature Selected Demo+Lancet+IDPs experiments. F) Raincloud plots of Matthews correlation coefficient.

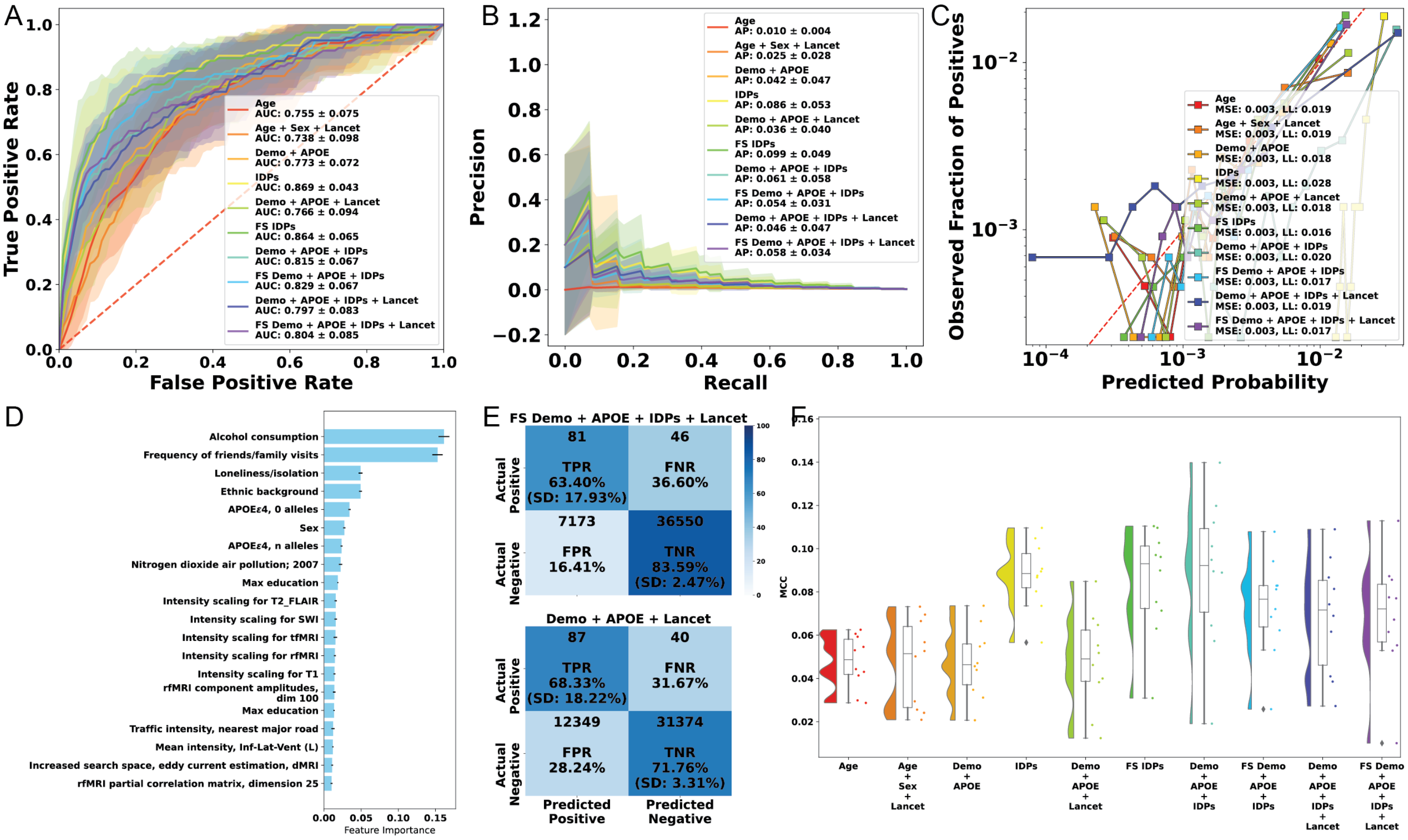


**Fig. S24**: Predicting all-cause dementia with brain imaging-derived phenotypes, using logit regression. A) Mean ROC curves. B) Mean precision-recall curves. C) Calibration curves (all folds combined). D) Mean feature importance plots. E) Confusion matrices for Demo+Lancet and Feature Selected Demo+Lancet+IDPs experiments. F) Raincloud plots of Matthews correlation coefficient.

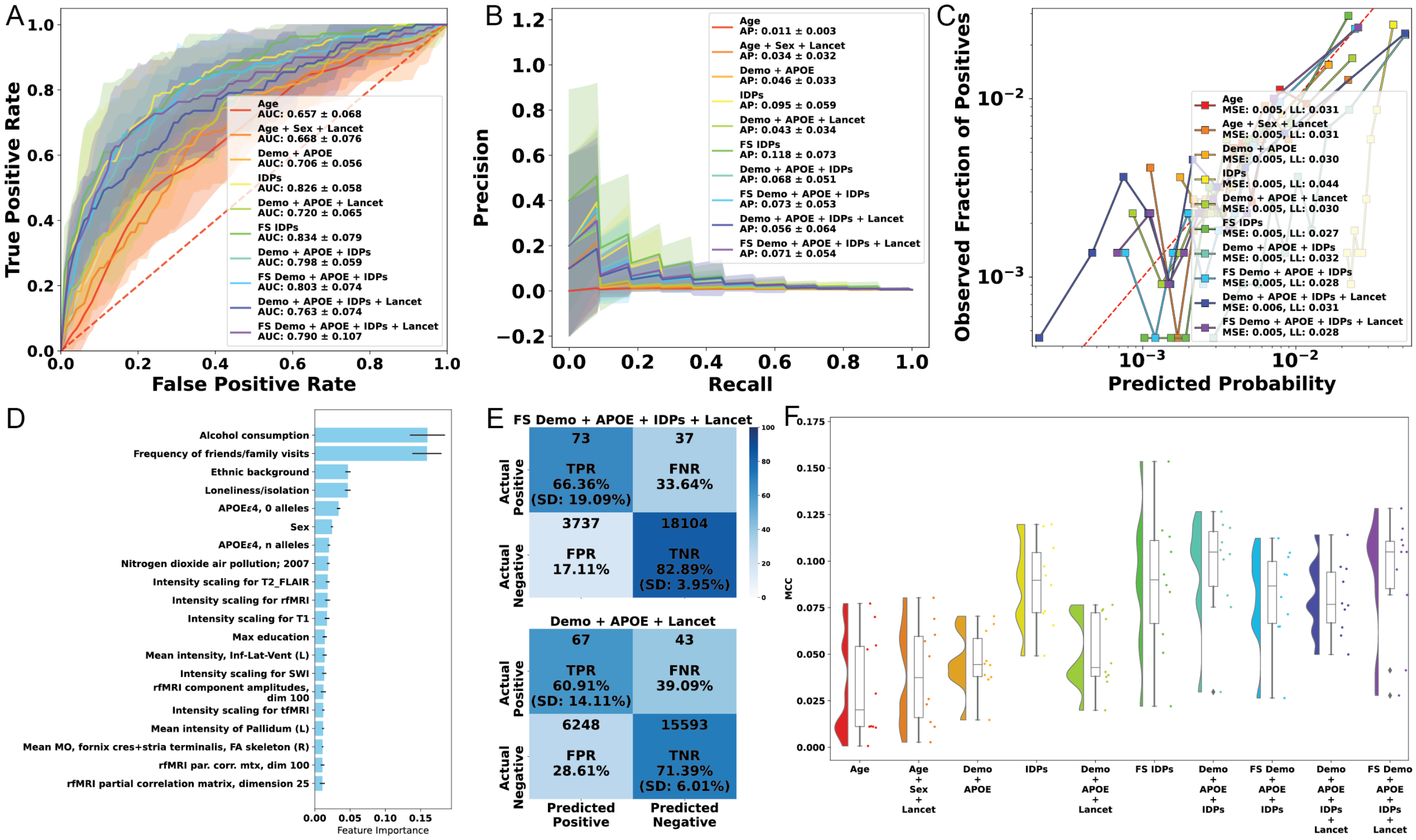


**Fig. S25**: Predicting all-cause dementia with brain imaging-derived phenotypes, only including participants aged 65 and older, using logit regression. A) Mean ROC curves. B) Mean precision-recall curves. C) Calibration curves (all folds combined). D) Mean feature importance plots. E) Confusion matrices for Demo+Lancet and Feature Selected Demo+Lancet+IDPs experiments. F) Raincloud plots of Matthews correlation coefficient.

##### 6.2.7 Cognitive tests in the UK Biobank

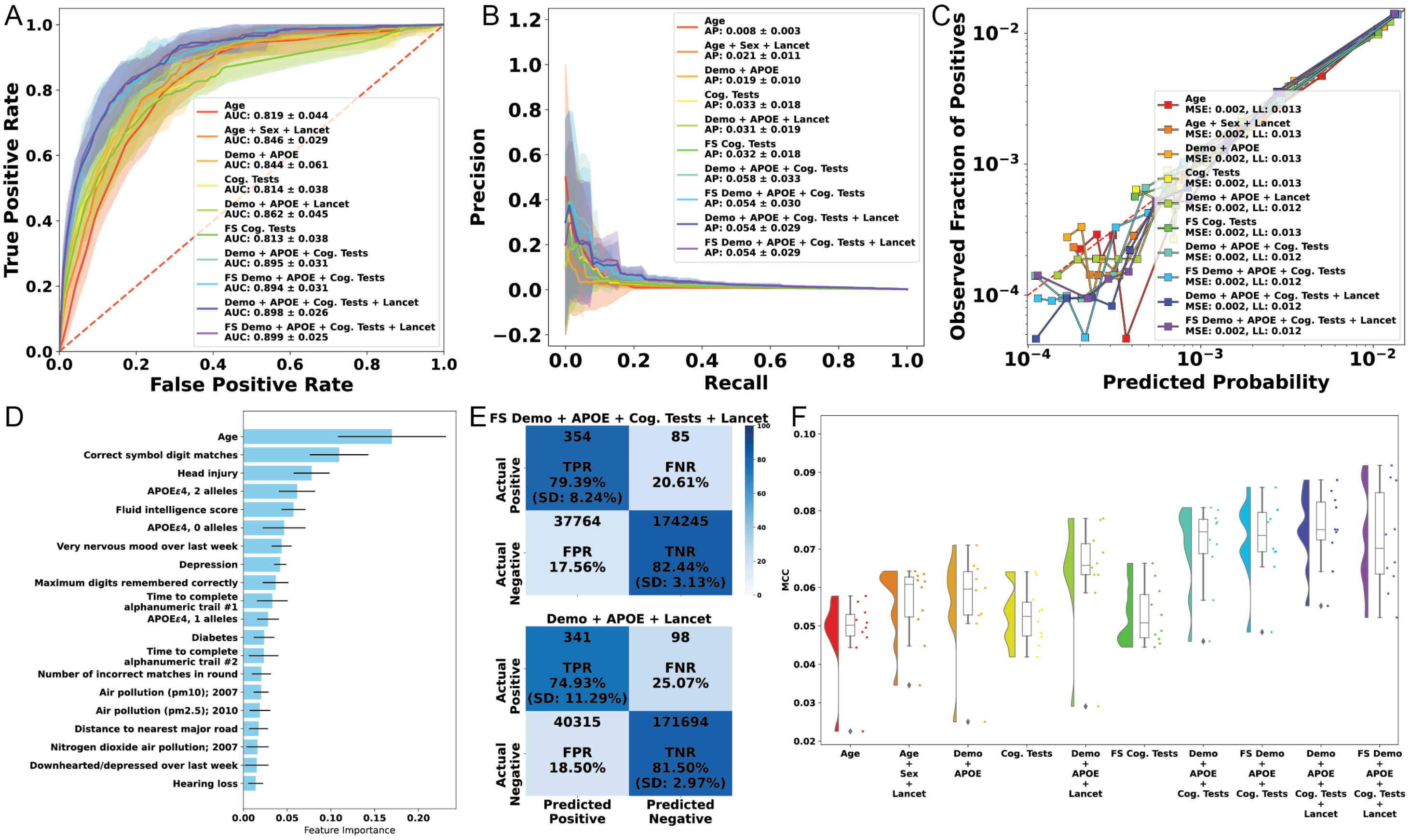


**Fig. S26**: Predicting Alzheimer's dementia with cognitive tests. A) Mean ROC curves. B) Mean precision-recall curves. C) Calibration curves (all folds combined). D) Mean feature importance plots. E) Confusion matrices for Demo+Lancet and Feature Selected Demo+Lancet+Cognitive tests experiments. F) Raincloud plots of Matthews correlation coefficient.

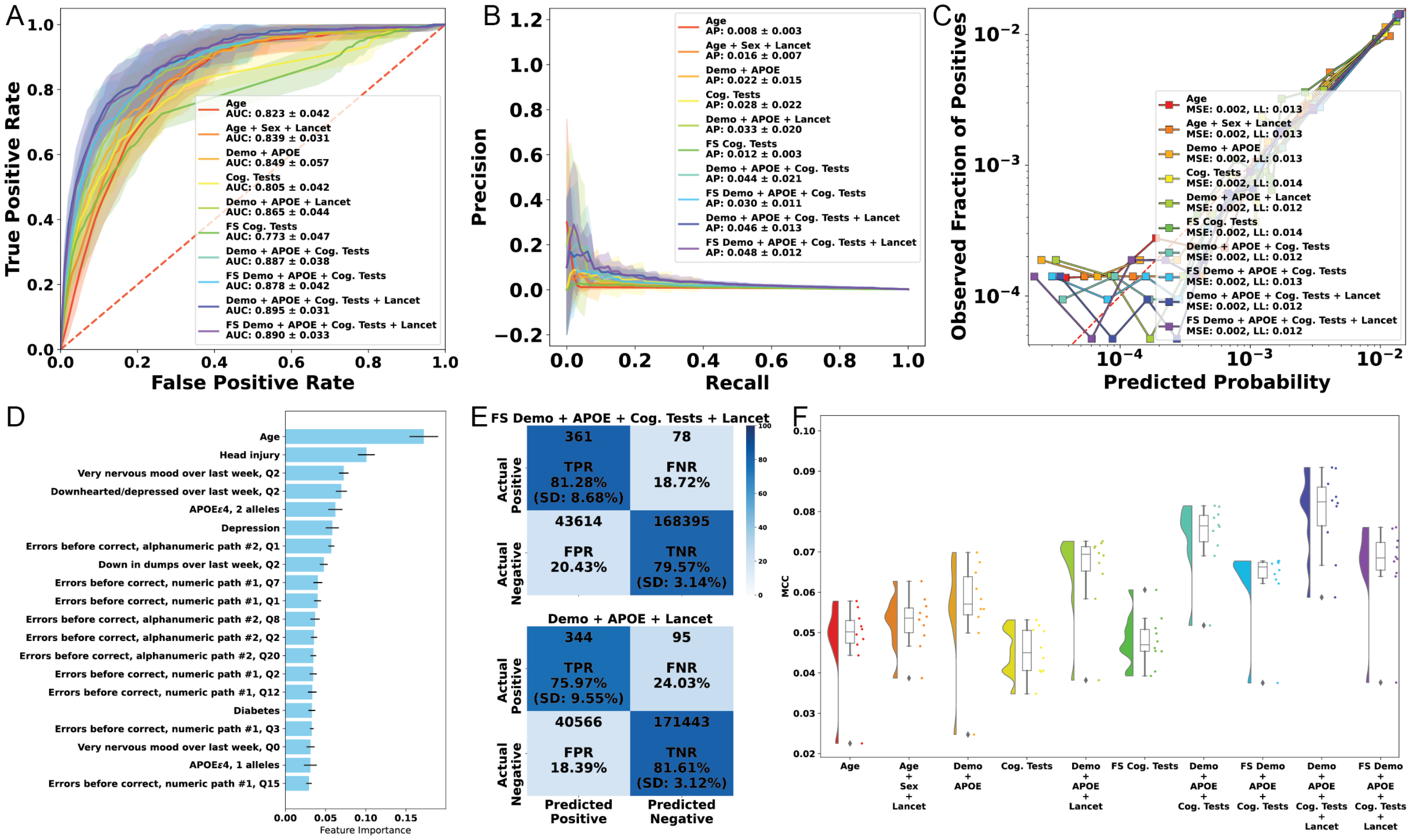


**Fig. S27**: Predicting Alzheimer's dementia with cognitive tests, using logistic regression instead of LightGBM. A) Mean ROC curves. B) Mean precision-recall curves. C) Calibration curves (all folds combined). D) Mean feature importance plots. E) Confusion matrices for Demo+Lancet and Feature Selected Demo+Lancet+Cognitive Tests experiments. F) Raincloud plots of Matthews correlation coefficient.

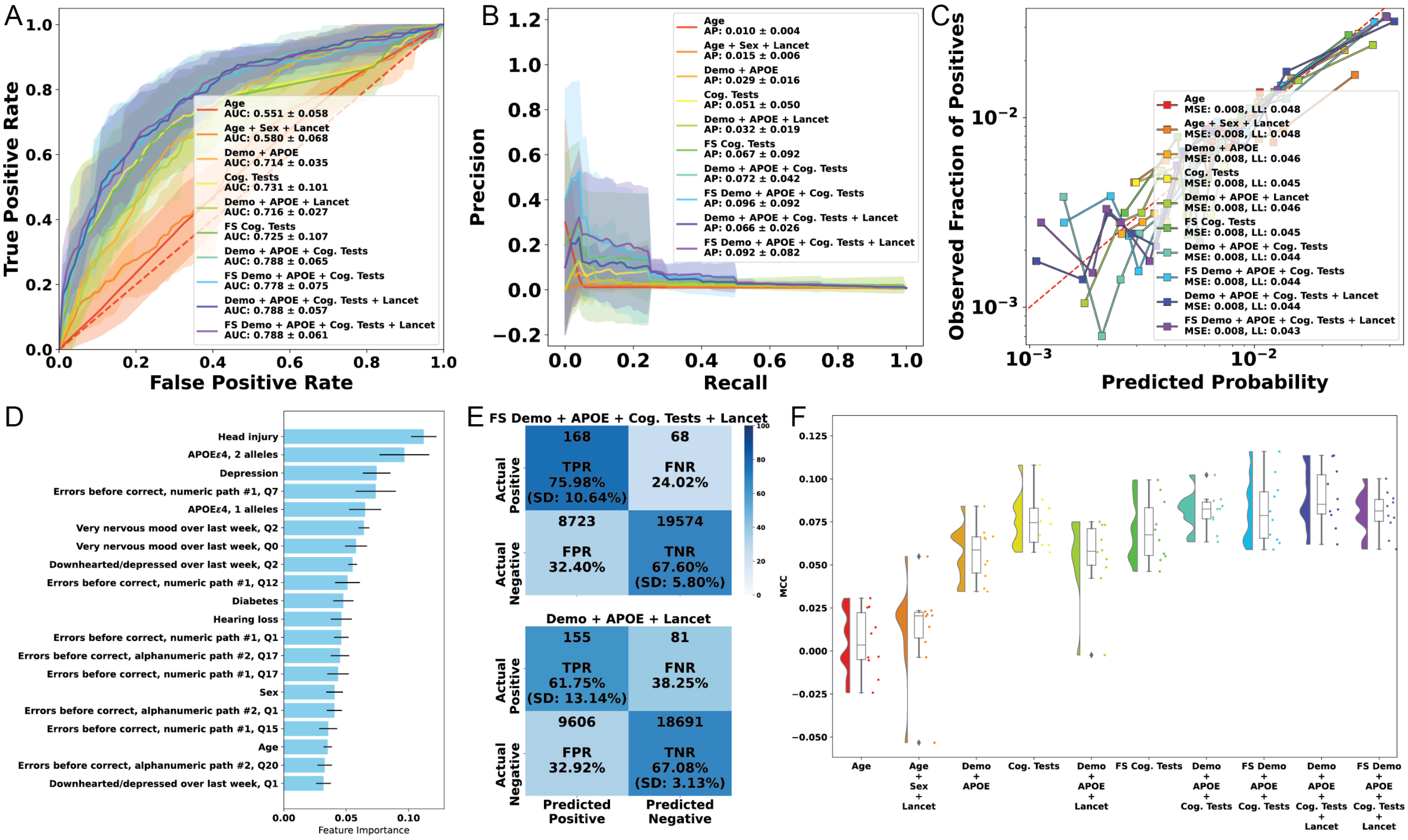

**Fig. S28**: Predicting Alzheimer's dementia with cognitive tests, only including participants aged 65 and older, with logistic regression instead of LightGBM. A) Mean ROC curves. B) Mean precision-recall curves. C) Calibration curves (all folds combined). D) Mean feature importance plots. E) Confusion matrices for Demo+Lancet and Feature Selected Demo+Lancet+Cognitive Tests experiments. F) Raincloud plots of Matthews correlation coefficient.

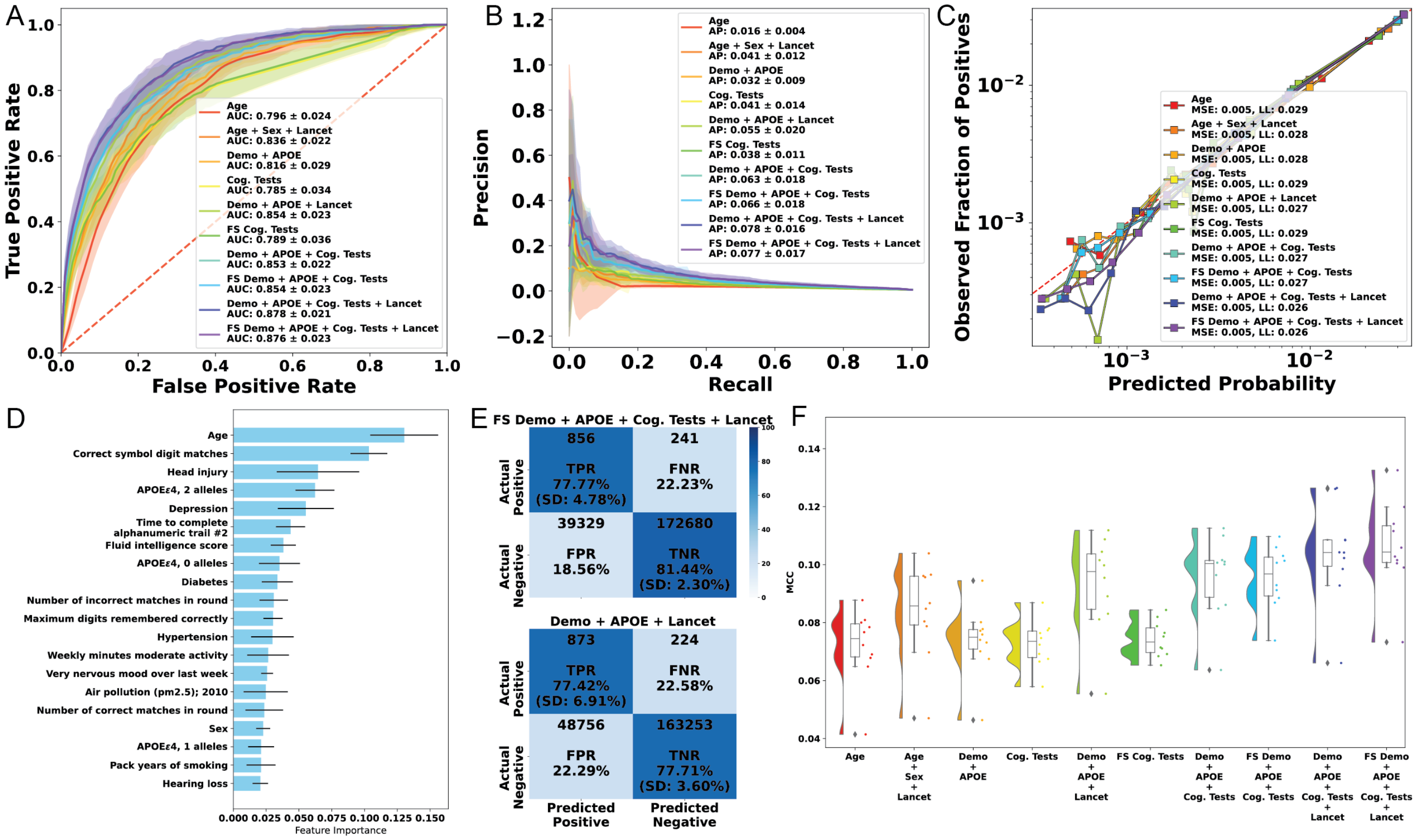


**Fig. S29**: Predicting all-cause dementia with cognitive tests. A) Mean ROC curves. B) Mean precision-recall curves. C) Calibration curves (all folds combined). D) Mean feature importance plots. E) Confusion matrices for Demo+Lancet and Feature Selected Demo+Lancet+Cognitive Tests experiments. F) Raincloud plots of Matthews correlation coefficient.

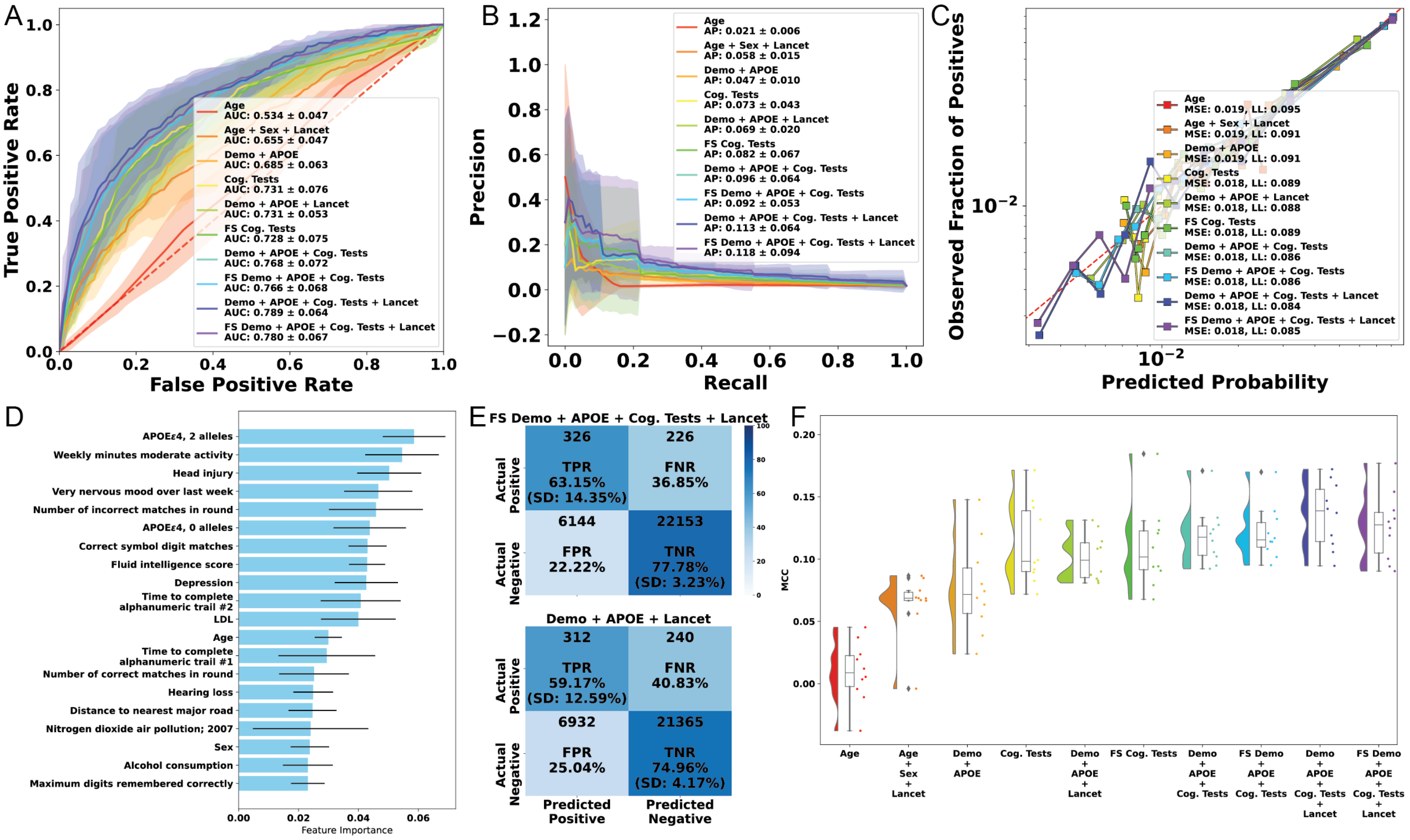


**Fig. S30**: Predicting all-cause dementia with cognitive tests, only including participants aged 65 and older. A) Mean ROC curves. B) Mean precision-recall curves. C) Calibration curves (all folds combined). D) Mean feature importance plots. E) Confusion matrices for Demo+Lancet and Feature Selected Demo+Lancet+Cognitive Tests experiments. F) Raincloud plots of Matthews correlation coefficient.

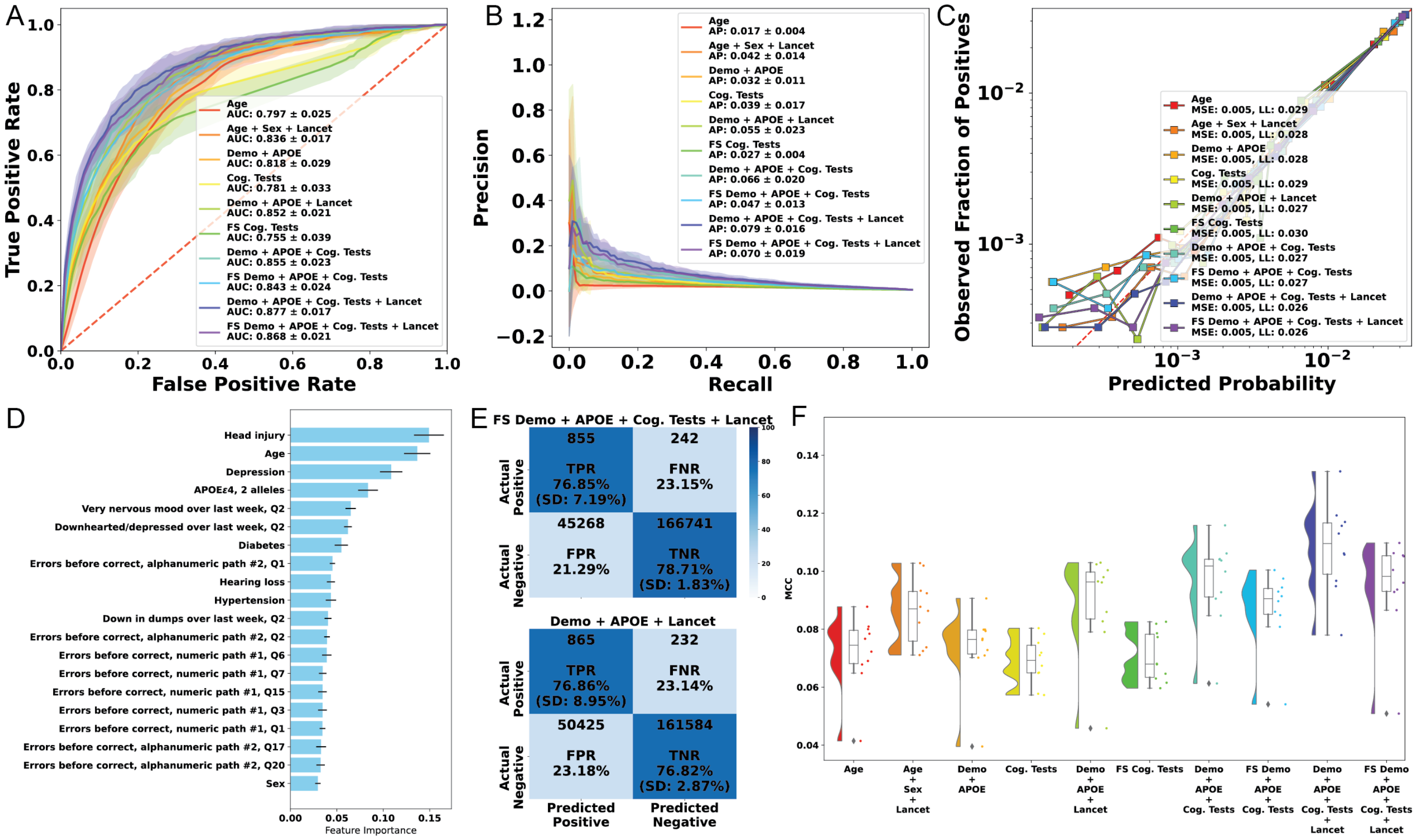


**Fig. S31**: Predicting all-cause dementia with cognitive tests, using logit regression. A) Mean ROC curves, using logit regression. B) Mean precision-recall curves. C) Calibration curves (all folds combined). D) Mean feature importance plots. E) Confusion matrices for Demo+Lancet and Feature Selected Demo+Lancet+Cognitive Tests experiments. F) Raincloud plots of Matthews correlation coefficient.

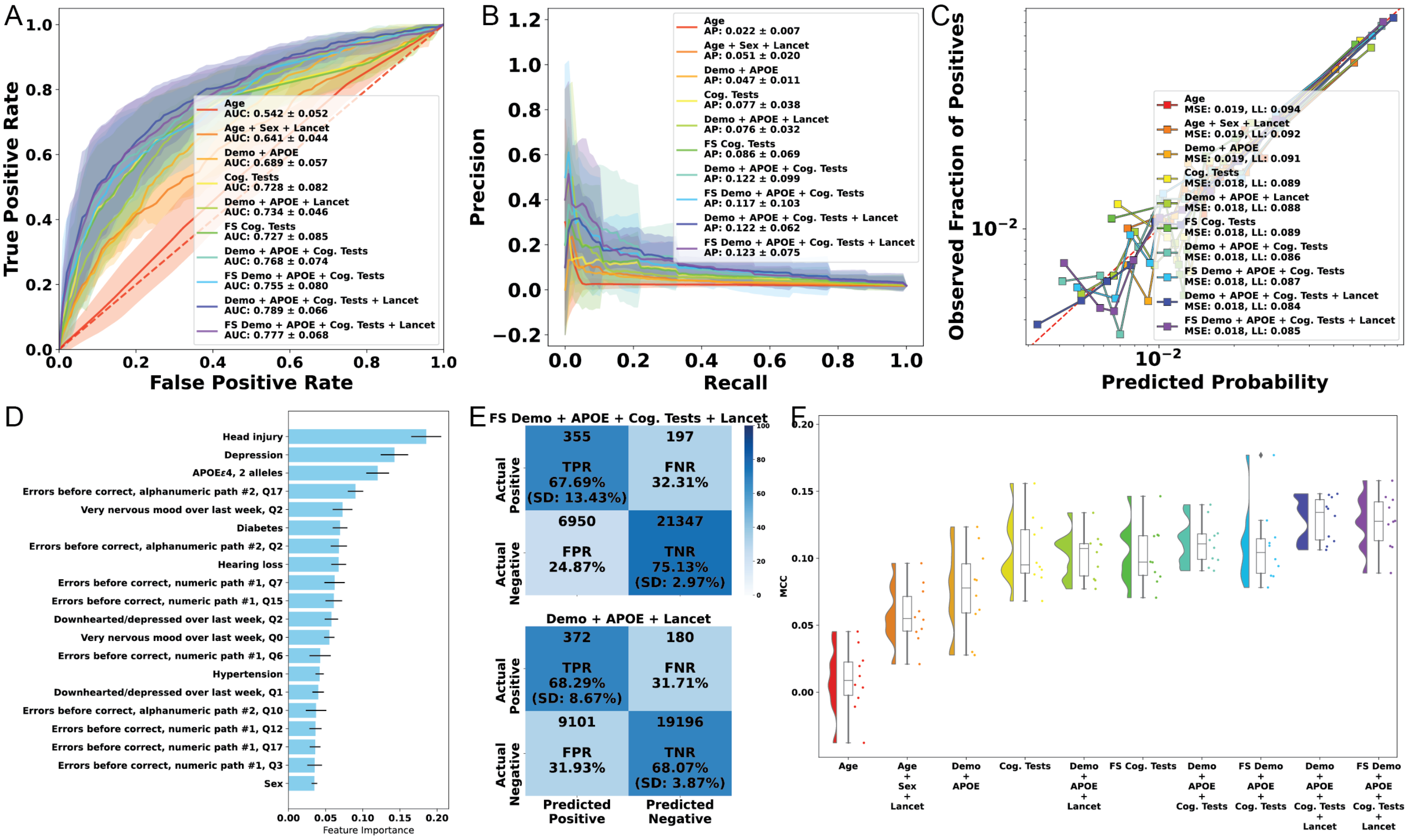


**Fig. S32**: Predicting all-cause dementia with cognitive tests, only including participants aged 65 and older, using logit regression. A) Mean ROC curves. B) Mean precision-recall curves. C) Calibration curves (all folds combined). D) Mean feature importance plots. E) Confusion matrices for Demo+Lancet and Feature Selected Demo+Lancet+Cognitive Tests experiments. F) Raincloud plots of Matthews correlation coefficient.

##### 6.2.8 NPV-Prevalence curves

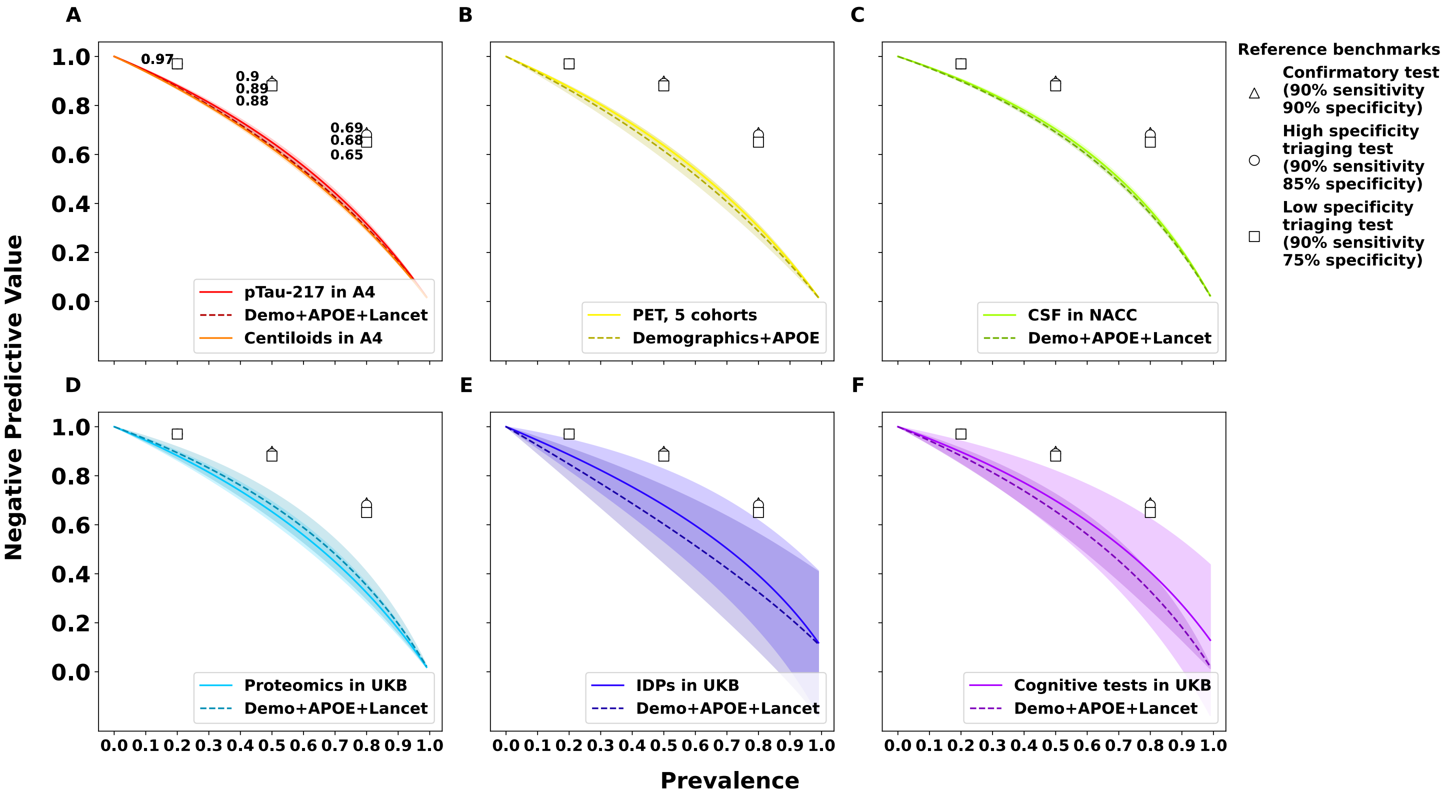


**Fig. S33**: Negative predictive value-prevalence curves for all six experiments, with reference benchmarks from Table 2 in[25] for confirmatory, high-specificity triaging, and low-specificity triaging tests at prevalences of 20%, 50%, and 80%. Sensitivities and specificities were fixed, with prevalence allowed to vary from 0 to 1 in step sizes of 0.01 to calculate positive predictive value. A) pTau217 predicting clinical impairment in A4/LEARN, B) PET predicting AD Dx in five cohorts, C) CSF predicting AD Dx in NACC, D) Plasma proteomics predicting AD ICD codes in the UK Biobank, E) Brain imaging-derived phenotypes predicting AD ICD codes in the UK Biobank, F) Cognitive tests predicting AD ICD codes in the UK Biobank.
